# Immunogenicity and seroefficacy of pneumococcal conjugate vaccines – a systematic review and network meta-analysis

**DOI:** 10.1101/2023.01.13.23284388

**Authors:** Shuo Feng, Julie McLellan, Nicola Pidduck, Nia Roberts, Julian PT Higgins, Yoon Choi, Alane Izu, Mark Jit, Shabir A Madhi, Kim Mulholland, Andrew J Pollard, Beth Temple, Merryn Voysey

## Abstract

**Background:** Vaccination of infants with pneumococcal conjugate vaccines (PCV) is recommended by the World Health Organisation. Evidence is mixed regarding the differences in immunogenicity and efficacy of the different pneumococcal vaccines.

**Methods:** In this systematic-review and network meta-analysis, we searched the Cochrane Library, Embase, Global Health, Medline, clinicaltrials.gov and trialsearch.who.int up to July 2022 (Protocol PROSPERO ID CRD42019124580). Studies were eligible if they presented data comparing the immunogenicity of either PCV7, PCV10 or PCV13 in head-to-head randomised trials for young children, and provided at least one time point after the primary vaccination series and/or one-month after a booster dose. Individual participant level data were requested from publication authors and/or the relevant vaccine manufacturer; aggregate data were extracted if individual data were unavailable. Outcomes included the geometric mean ratio (GMR) of serotype-specific IgG and relative risk (RR) of seroinfection. Seroinfection is defined as a rise in antibody between the primary vaccination series and the booster dose, as evidence of subclinical infection. We also estimated the relationship between the GMR one month after priming and the RR of seroinfection by the time of the booster dose.

**Findings:** In total 45 studies were eligible from 38 countries across six continents. 27 and 12 studies with data available were included in immunogenicity and seroefficacy analyses respectively. GMRs comparing PCV13 vs PCV10 favoured PCV13 for serotypes 4, 9V, and 23F at 1 month after primary vaccination series, with 1.14- to 1.54-fold significantly higher IgG responses with PCV13. Risk of seroinfection prior to the time of booster dose was lower for PCV13 for serotype 4, 6B, 9V, 18C and 23F than for PCV10. Two-fold higher antibody after primary vaccination was associated with 54% decrease in risk of seroinfection (RR 0.46, 95%CI 0.23-0.96).

**Conclusion:** Serotype-specific differences were found in immunogenicity and seroefficacy between PCV10 and PCV13. Higher immunogenicity of PCVs are associated with lower risk of subsequent infection. These findings could be further used to compare PCVs and optimise vaccination strategy.

**Funding:** This study is funded by the NIHR Health Technology Assessment programme (17/148/03).

## Introduction

*Streptococcus pneumoniae* (pneumococcus) causes severe disease including bacterial pneumonia, meningitis, and sepsis, leading to substantial morbidity and mortality worldwide, with the highest disease burden being in young children and older adults.^1,2^ There have been more than 100 serotypes of pneumococcus documented as of 2020, not all of which cause severe disease, and the distribution of these serotypes varies substantially between countries.^1,2^ Three pneumococcal conjugate vaccines (PCV)s, have been widely deployed worldwide in the past two decades: PCV7 (Prevnar, Pfizer), PCV10 (Synflorix, GSK) and PCV13 (Prevenar 13, Pfizer), resulting in substantial reduction in disease.^1,3^ New PCVs such as PCV15, PCV20 and PCV10-SII have been licensed in some countries but have yet to be widely deployed.

Between 2009 and 2011, PCV7 was gradually replaced by PCV13 and PCV10 and is no longer available. Currently, three PCVs are recommended by the World Health Organisation (WHO) for infants worldwide: PCV13, PCV10, and a new 10-valent PCV manufactured by Serum Institute of India (PCV10-SII, PNEUMOSIL) which was prequalified by WHO in December 2019.^4-6^ PCV13 provides three serotypes (3, 6A and 19A) to the 10 serotypes included in PCV10 (serotype 1, 4, 5, 6B, 7F, 9V, 14, 18C, 19F, and 23F). PCV10-SII covers serotypes 1, 4, 5, 6B, 7F, 9V, 14, 18C, 19F and 23F. The licensure of PCVs is benchmarked against anti-capsular IgG antibody responses above a threshold of 0.35 mcg/mL for all vaccine serotypes, which was established using data from three randomised controlled efficacy trials.^7^ Real world evidence suggests that correlates of protection and effectiveness against invasive pneumococcal disease vary across serotypes.^8^

The WHO does not preferentially endorse one PCV over another. Both PCV13 and PCV10, have been shown to provide both direct and indirect protection against pneumococcal pneumonia, invasive pneumococcal disease and nasopharyngeal carriage.^3,6,9^ Although there are 10 common serotypes in these two vaccines the content of the vaccines differ, with different carrier proteins used in the conjugation process, as well as different amounts of polysaccharide, and these differences may contribute to differences in protection. In 2017 a systematic review of head-to-head studies comparing PCV10 vs PCV13 showed differences in anti-pneumococcal IgG responses between vaccines. However, no meta-analysis has included in this review and there remains uncertainty over whether one vaccine is consistently more immunogenic, and whether differences in immunogenicity result in clinically important differences in protection. Large head-to-head randomised controlled trials of PCVs with invasive pneumococcal disease as the primary outcome are not feasible. Studies that assessed the impact of different PCVs on nasopharyngeal carriage have reported very few or no differences.^10,11^ Episodes of nasopharyngeal carriage often last only a few days or weeks therefore cross-sectional swabbing studies may misclassify participants when swabs are not taken at the time of infection, resulting in underpowered comparisons. We previously used “seroefficacy” as an outcome for estimating correlates of protection for PCVs against pneumococcal carriage,^12^ where seroinfection is defined as an increase in antibody levels between the primary vaccination series (typically complete at 5-7 months of age) and the booster dose (typically administered at 9-18 months of age). Seroinfection can be regarded as evidence of exposure to the pathogen and a resultant sub-clinical infection, given antibody responses wane rapidly during this period otherwise.^12^

In this study, we meta-analysed individual participant data from head-to-head studies of PCVs to compare the immunogenicity and seroefficacy of PCV10 with PCV13 for each serotype. We aimed to determine if serotype specific immune responses were higher for either vaccine, and whether this resulted in greater protection again carriage (seroefficacy) for the same serotypes. In addition, we explored the overall relationship between the higher immune response and protection against carriage in infants.

## Methods

Our systematic review is reported in line with the recommendations from the Preferred Reporting Items for Systematic Reviews and Meta-Analyses statement plus the extension statements for network and individual patient data systematic reviews.^13-15^

### Primary and secondary objectives

The primary objective was to compare the immunogenicity of PCV10 vs PCV13 for each serotype contained in the vaccines. The secondary objectives were: 1) to compare the seroefficacy of PCV10 vs PCV13 for each serotype contained in the vaccines, 2) for PCV10 and PCV13 separately, to estimate immunogenicity and seroefficacy in comparison to the older PCV7 vaccine, and 3) to determine how the comparisons of immunogenicity and efficacy of PCV10 to PCV13 are affected by the co-administration of different routine vaccines.

### Systematic review

We conducted a systematic review identifying studies that compared the immunogenicity of licensed PCVs for infants or children in head-to-head randomised trials. The PCVs included in the review were PCV13, PCV10 and PCV7. The last was included so that we could compare PCV13 and PCV10 indirectly through them each being compared with PCV7 for the same serotypes. Methods for search strategy, study selection, data retrieval and assessment of risk of bias are described in the supplementary material.

### Outcomes

The primary outcome was serotype-specific anti-capsular pneumococcal immunoglobulin G. Antibodies measured one-month after the primary series of 1-3 doses in infancy, prior to a booster dose, and one-month post-booster dose were included. The outcome for seroefficacy analyses was the difference between log_10_- transformed serotype-specific anti-pneumococcal IgG measured one-month after the primary series of doses and prior to administration of the booster dose.

### Statistical Analysis

#### Immunogenicity

Each trial that had individual participant level data available was analysed to obtain the log of the ratio of geometric means (log-GMR) and its standard error, for each serotype and time point of interest. If individual participant data were unavailable, published GMR estimates and confidence intervals were used. The estimates combined from individual participant data and aggregate data formed the input data for data synthesis. Sensitivity analysis for immunogenicity results were conducted by restricting analyses to only those studies providing data for all three time points of interest.

#### Seroefficacy

We defined seroinfection as a rise in anti-serotype-specific IgG between the post-primary vaccination timepoint and the booster dose. As a binary variable, seroinfection was equivalent to 1 if antibody levels increased by any amount, or 0 otherwise. The relative risk (RR) of seroinfection was then estimated by comparing the proportion of participants with seroinfection between vaccine groups. When no seroinfection occurred in any group (numerator of absolute risk was 0), a small nonzero value (0.5) was added to both numerator and denominator to allow estimation of the RR. The log-RRs and their standard errors were then the input data for evidence synthesis. Only trials supplying individual participant data were included in seroefficacy analyses.

#### Data synthesis by network meta-analysis and meta-analysis

Serotypes 4, 6B, 9V, 14, 18C, 19F, and 23F were contained in all three vaccines, therefore evidence could be synthesized using a network meta-analysis of all comparisons between PCVs, including PCV7 (see Supplementary Figure 1). Serotypes 1, 5, 7F, 3, 6A and 19A are only included in PCV10 and PCV13 vaccines therefore for these serotypes evidence was synthesized by meta-analysing studies that directly compared PCV13 vs PCV10.

For the analysis of immunogenicity, we synthesized evidence for all PCV13 serotypes, However, seroefficacy could only be assessed in situations where the serotypes of interest were included in both vaccines and therefore seroefficacy of serotypes 3, 6A, and 19A could not be assessed as these are only included in one vaccine (PCV13). The assessment of heterogeneity and inconsistency of NMA is described in the supplementary material.

#### Association between ratios of immunogenicity and seroefficacy

To estimate separate serotype-specific relationships between the GMRs and RRs, study level data were combined regressing the RR of seroinfection on the GMR using linear regression models weighted by the sample size of the study. Weighted Pearson correlation coefficients were calculated.

To estimate the overall association between antibody GMR and RR across all serotypes, we fitted a mixed-effect model regressing study-level RRs of seroinfection on GMRs across serotypes, weighted by the sample size of each study. Fixed-effects included GMR, serotype, and interactions between GMR and serotype (allowing serotype-specific association), while study was included as a random effect. As a sensitivity analysis, we reversed both RRs and GMRs estimated (i.e. PCV13 vs PCV7 was changed to PCV7 vs PCV13). By shifting comparators, we aimed to evaluate of the stability of the association estimates.

PCV10 and PCV13 are manufactured slightly differently, with different carrier proteins, conjugation process, polysaccharide concentrations and sources. To evaluate if these differences between two products change the relationship between antibody levels and protection against seroinfection, we assessed the association between immunogenicity and seroefficacy restricting to studies that compared PCV13 vs PCV10 and PCV7 vs PCV10 only (comparisons between PCV13 and PCV7 were removed from analysis as these vaccines are from the same manufacturer). We examined whether PCVs of different manufacturers that produce equivalent levels of antibody (GMR=1) also provide comparable seroefficacy (RR=1).

All analyses were performed in R 4.2.2. NMA and meta-analysis were conducted using the netmeta and metafor packages.^16,17^

## Results

### Search results

Database registry and hand searches identified 4466 publication records (Figure 1), of which 45 studies (745 publication records) satisfied our eligibility criteria. ^10,11,18-89^18 studies (20 publication records) were excluded from the analysis: 6 studies did not provide individual patient or aggregate data,^70-73^ and 12 studies (14 publication records) were head-to-head studies with the vaccines of interest, but it was not possible to form a loop within the network meta-analysis to provide indirect evidence (See Supplementary Figure 1 and Supplementary Table 2).^74-89^ Of these 12 studies, 8 reported results from different PCVs including a new Cuban PCV7, PCV10-SII, PCV11, PCV12, a Chinese PCV13, PCV14, PCV15, PCV20, PCV24 and PCV SP0202-VI. The remaining 27 studies (53 publication records) from 2009 to 2022 were included in the network meta-analyses.^10,11,18-69^ 22 studies provided individual patient data with a further 5 studies reporting aggregate data (Table 1).

**Figure 1.**
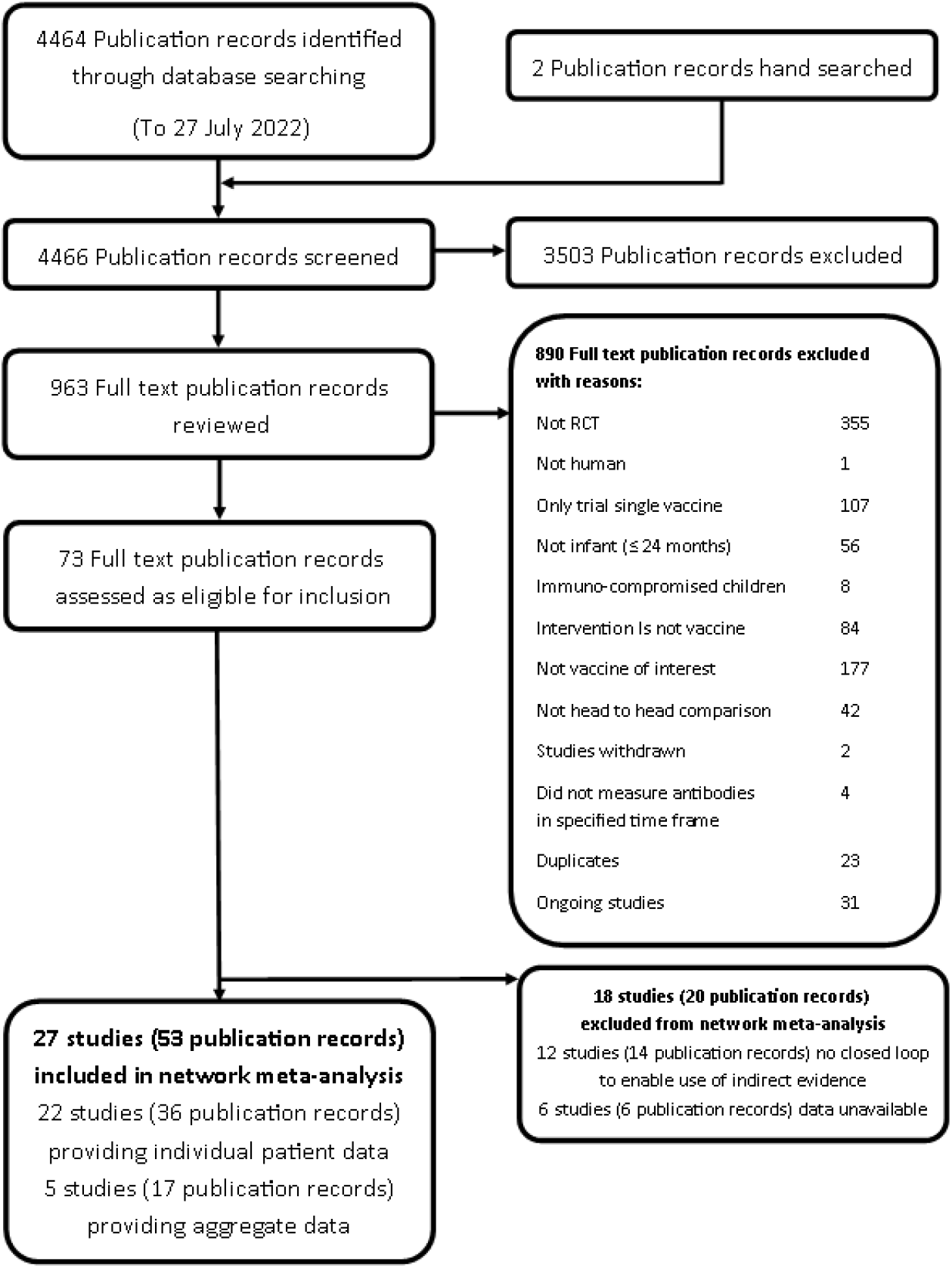
PRISMA flow diagram to show study selection process.

**Table 1.**
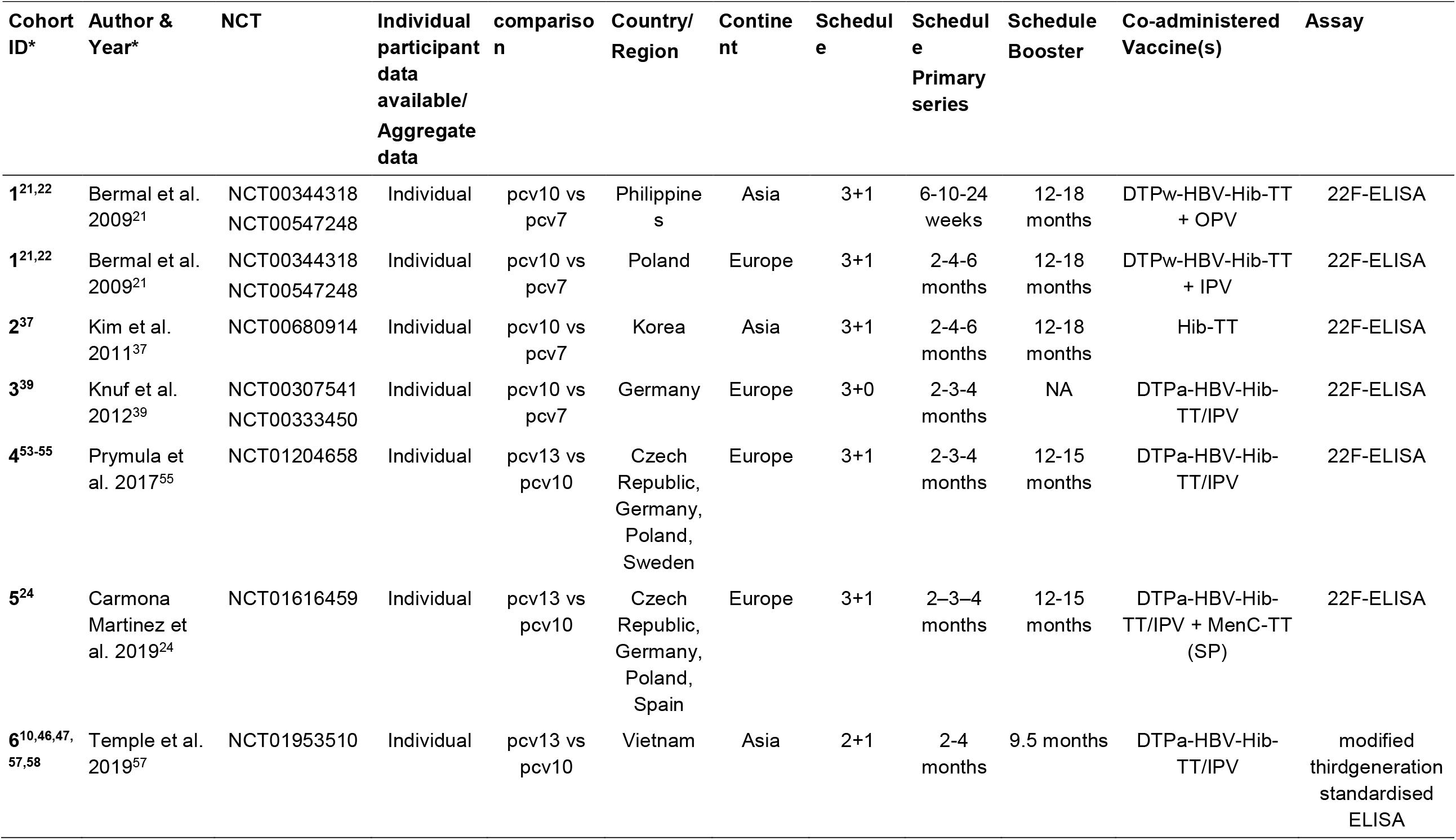
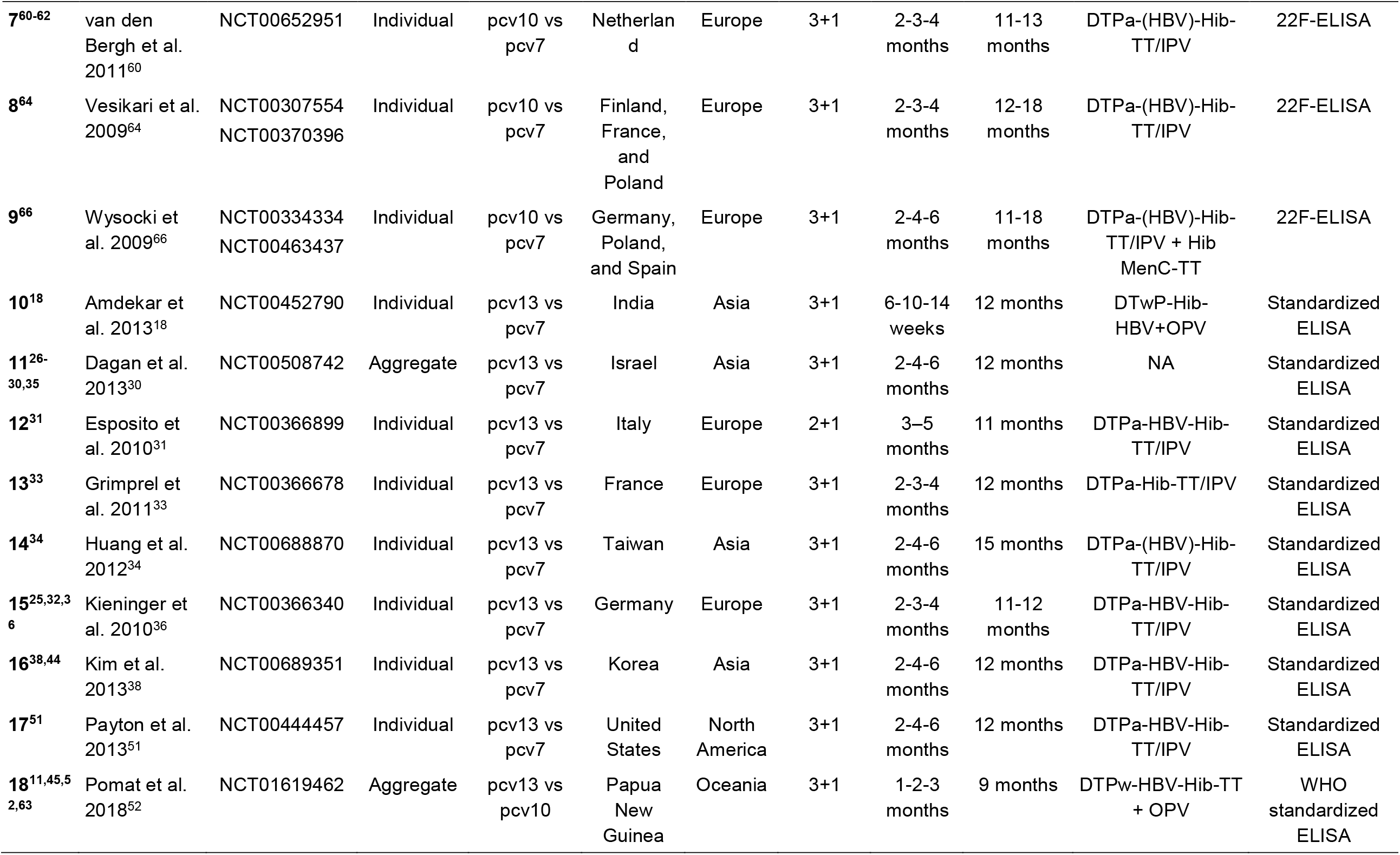
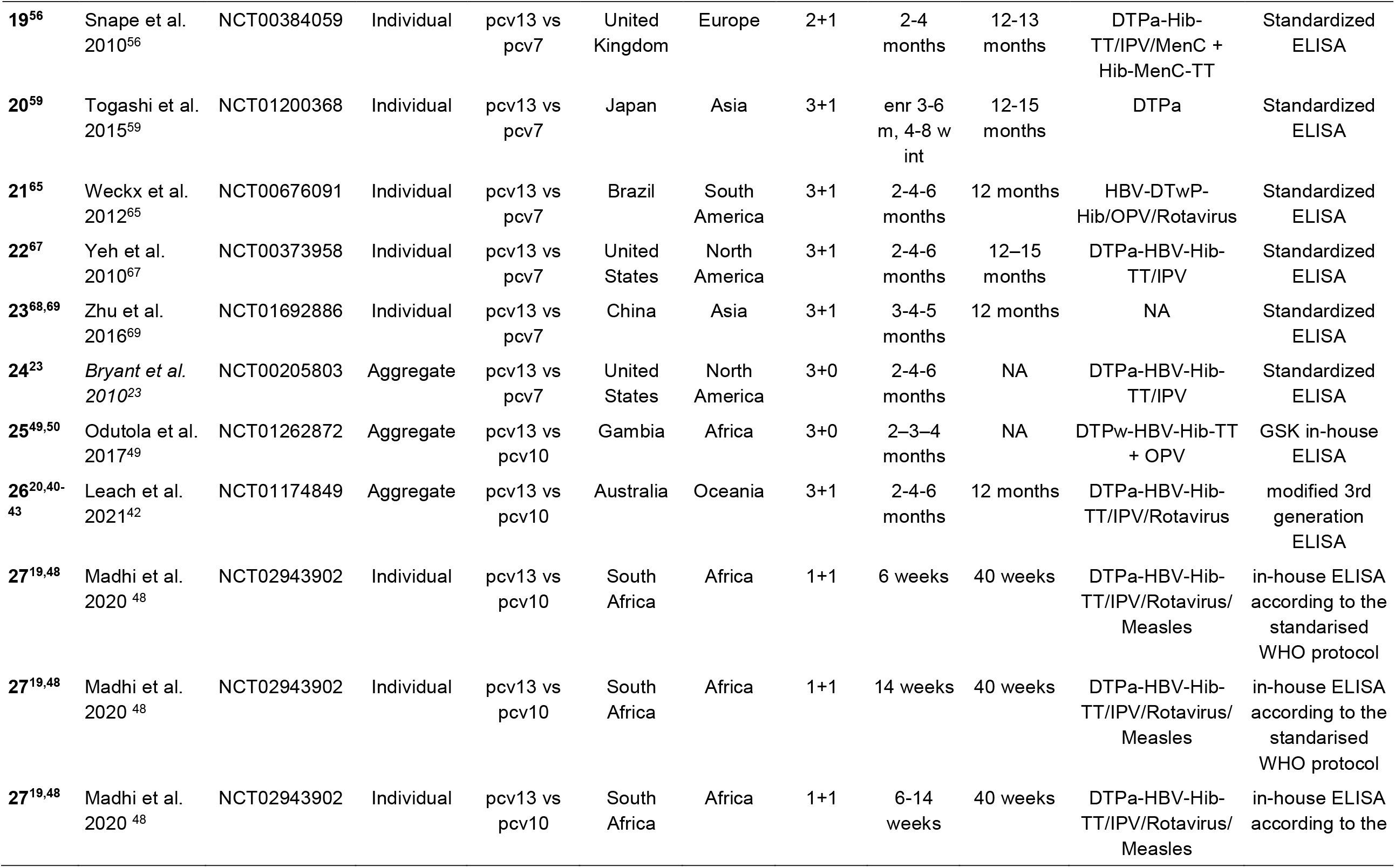
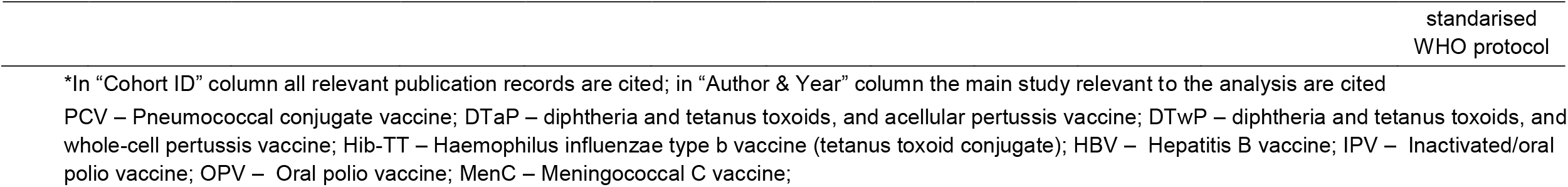
Summary of studies included in immunogenicity and seroefficacy analyses.

The 27 included studies comprised 30 cohorts of children as one study conducted in two countries reported results separately,^21,22^ and one study included head-to-head comparisons of 3 vaccination schedules^19,48^ (Table 1). Studies with multiple NCT numbers or publications but the same population were counted as one cohort. These 30 cohorts were representative of 38 countries in six continents – Europe (n=11 cohorts), Asia (n=9 cohorts), North America (n=3 cohorts), Africa (n=2 cohort), Oceania (n=4 cohort) and South America (n=1 cohort). Four cohorts were from studies conducted in multiple countries in Europe and analysis were combined across sites. ^24,53-55,64,66^

There were 7 studies comparing PCV10 vs PCV7, 14 studies comparing PCV13 vs PCV7, and 7 studies comparing PCV13 vs PCV10 (Supplementary Figure 1b). Two cohorts used a 1+1 schedule with the first dose administered at either 6- or 14-weeks of age to South African infants and compared PCV13 with PCV10.^48^ Four cohorts used a 2+1 prime-boost schedule: one study in Vietnam comparing PCV13 vs PCV10,^57^ one in South Africa comparing PCV13 vs PCV10 with additional comparisons with 1+1 schedules^48^, and two from studies conducted in Europe comparing PCV13 vs PCV7.^31,56^ Three cohorts used a 3+0 schedule: one in the Gambia comparing PCV13 vs PCV10,^50^ one in the United States comparing PCV13 vs PCV7^67^ and one in Germany comparing PCV10 vs PCV7.^39^ The remaining 20 cohorts tested a 3+1 schedule, with most cohorts receiving a primary series at 2-4-6 months (n=9) and a booster at around 12 months (n=18). Infants’ age at receipt of the first dose ranged from 1 month to 3.5 months, and the age of the booster dose ranged from 9 to 18 months, resulting in an interval between primary and booster dose (used for the calculation of seroefficacy) of between 6 to 12 months. Most cohorts reported or cited types of co-administered vaccines (n=24), and PCVs were commonly co-administered with routine childhood vaccine including diphtheria, tetanus, and acellular/whole-cell pertussis vaccine (DTaP/DTwP, n=23), haemophilus influenzae type b vaccine with tetanus toxoid conjugate (Hib-TT, n=23), hepatitis B vaccine (HepB, n=20), inactivated/oral polio vaccine (IPV/OPV, n=22), and meningococcal C vaccine (MenC, n=3)(Table 1). Serotype-specific IgG antibody responses were defined as primary outcomes in all studies. Studies comparing PCV10 vs PCV7 (n=7) assessed serotypes included in PCV10, while all other studies assessed all serotypes included in PCV13. Geometric mean concentrations (GMC) were reported at 28 days post-primary series (n=29 cohorts), prior to a booster (n=17 cohorts) and 28 days post-booster (n=25 cohorts). Fourteen cohorts (46.7%) reported GMC at all three time points. Individual participant data were available from 25 of 30 (83.3%) cohorts.

### Assessment of risk of bias

Risk of bias assessments for the 27 included studies are summarised in Supplementary Figure 2. Results of ten studies^31,33,36,38,51,55,56,65,67,69^ were assessed to be at ‘low risk of bias’ across all domains and overall. Two studies^23,66^ had results judged to be at ‘high risk of bias’ due to problems identified in one domain each: Wysocki 2009^66^ only analysed immunogenicity for a subset of participants and Bryant 2010^23^ did not report whether participants or staff delivering the intervention were blinded to the vaccine received. Lack of information was reported in Bryant 2010^23^ for the analysis, raising concerns on appropriateness of the analysis for the aggregate data obtained from this study. The remaining 15 studies^18,21,24,30,34,37,39,42,48,49,52,57,59,60,64^ were judged to have ‘some concerns’ over risk of bias. These concerns predominantly arose because the randomisation process was not described, and/or the study did not report if the participants or staff delivering the vaccines were blinded to which vaccines were given.

### Immunogenicity

Figure 2 shows the number of study cohorts included in each analysis and the estimated GMR for each serotype and time point from the network meta-analysis/meta-analysis, and Supplementary Table 3 summarises the heterogeneity statistics and inconsistency of the network. Substantial heterogeneity and network inconsistency were present for most serotypes at all three time points.

Direct (comparisons between PCV10 and PCV13) and indirect (comparisons of PCV13 vs PCV7 and PCV10 vs PCV7) evidence from 28 cohorts were available for immunogenicity analysis at 28 days post-primary vaccination (Supplementary Figure 3a). GMRs comparing PCV13 vs PCV10 for any primary series schedule were higher in PCV13 for serotypes 4, 7F, 9V, and 23F at 1 month after primary vaccination series, with 1.14- to 1.54-fold significantly higher IgG responses in PCV13. Additional serotypes contained only in the PCV13 vaccine (3, 6A and 19A) also favoured PCV13 as expected. GMRs were similar for the remaining serotypes (1, 5, 6B, 14, 18C, 19F, Figure 2a).

**Figure 2.**
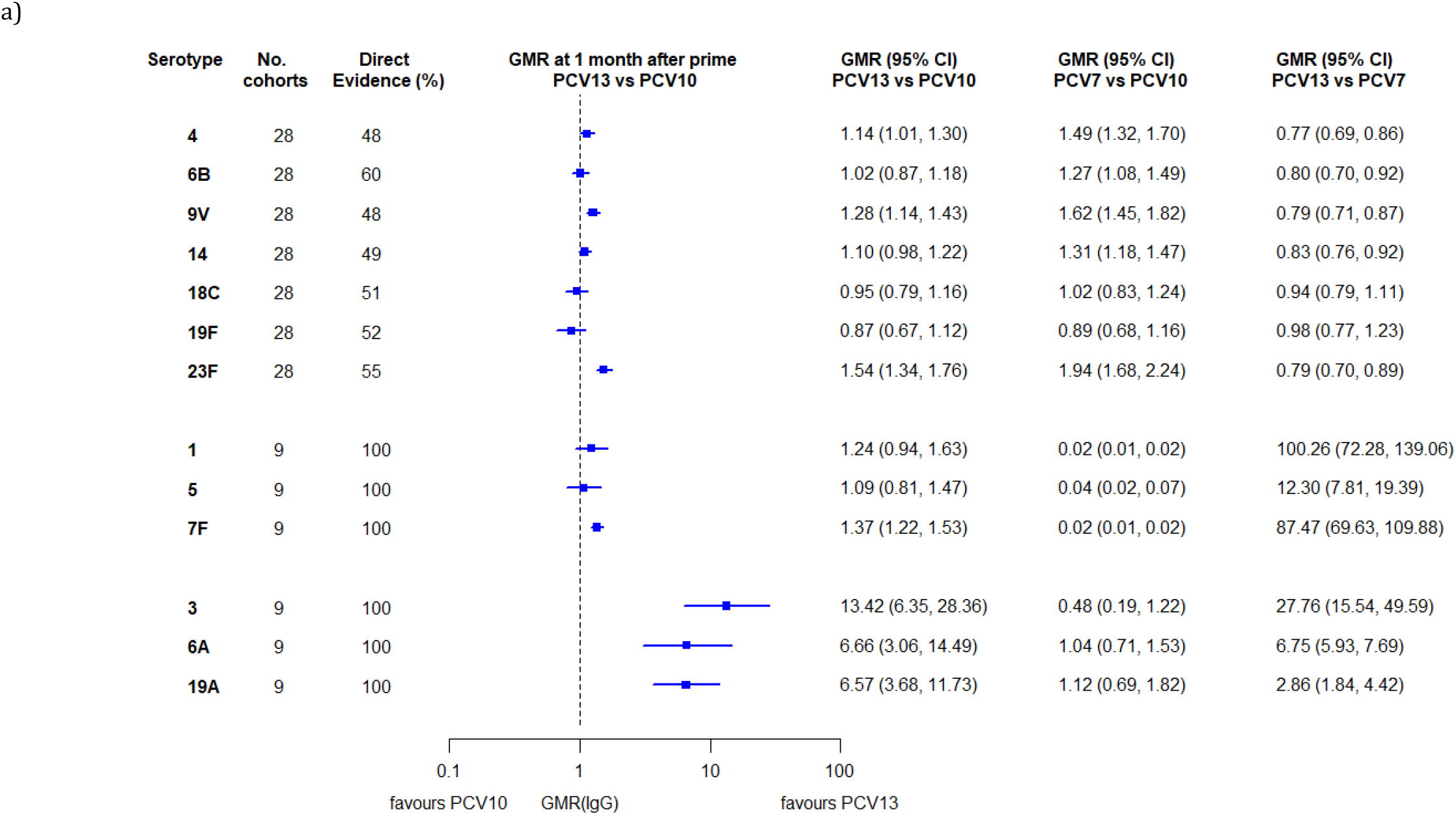
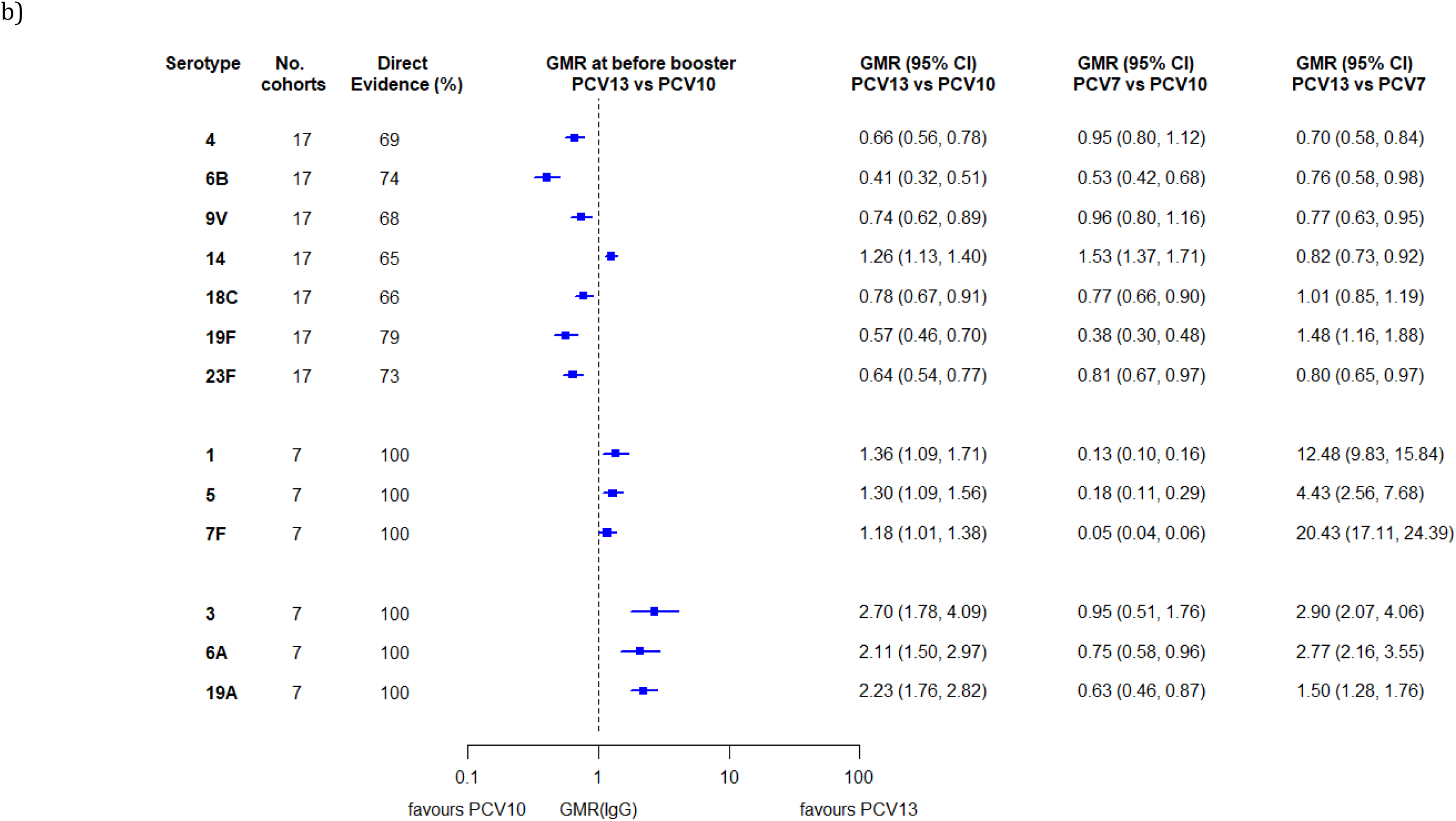
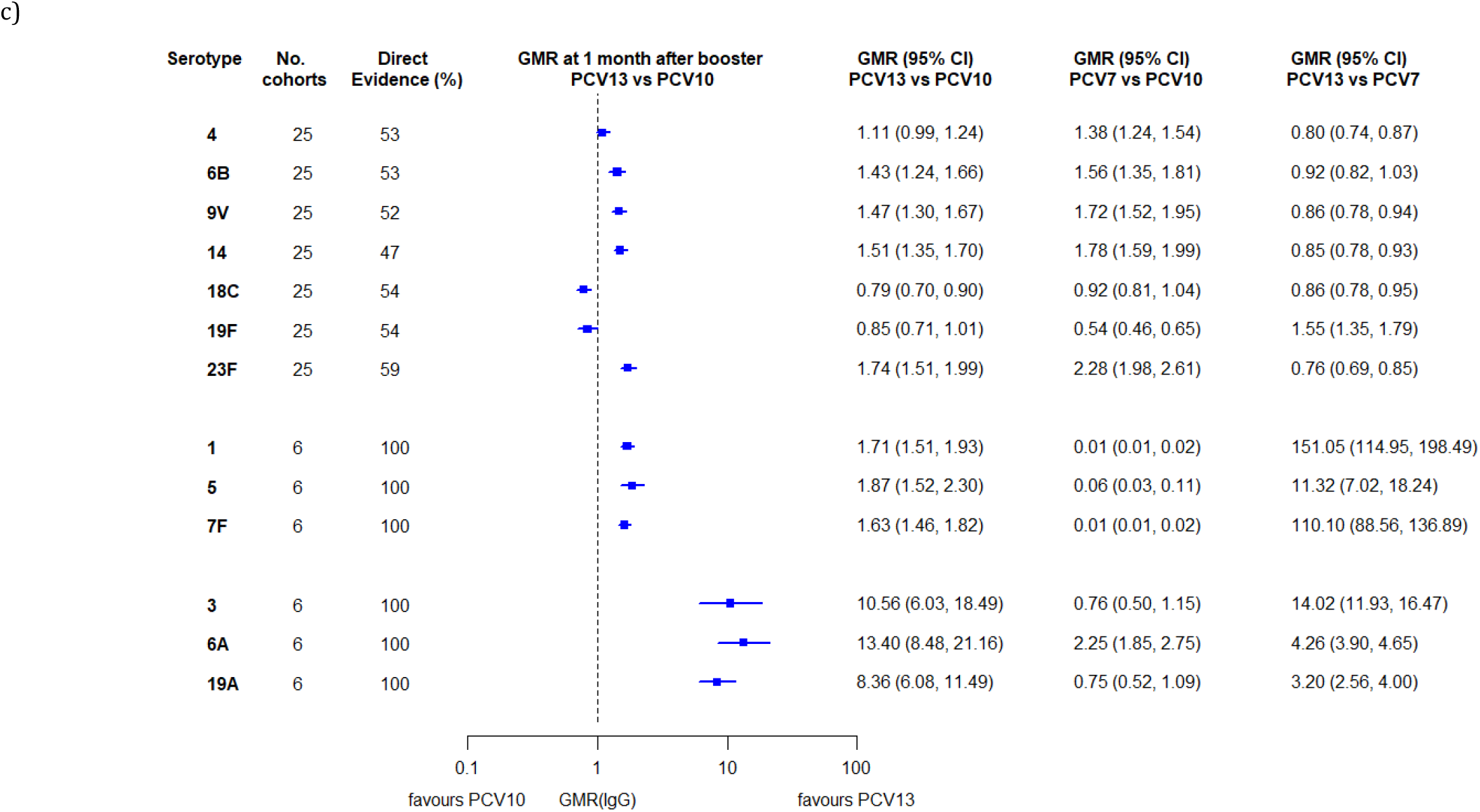
Geometric mean ratios from meta-analyses of head-to-head studies at a) 28 days post-primary vaccination series, b) pre-booster, and c) 28 days post-booster. GMR: Geometric mean ratio; PCV: Pneumococcal conjugate vaccine. Each line in the figure shows the output from a network meta-analyses (PCV7 serotypes) or direct meta-analyses (PCV13 but non-PCV7 serotypes). Blue boxes and blue lines show the point estimates and confidence intervals for geometric mean ratios comparing PCV13 vs PCV10. Points to the right of the vertical line are those with higher antibody responses in the PCV13 arm of the study, and points to the left are those with higher antibody responses in the PCV10 arm. The direct evidence column shows the percentage of evidence from studies directly comparing PCV13 vs PCV10 that contributes to the estimates presented in the figure in blue (PCV13 vs PCV10). GMR of PCV13 vs PCV10 for PCV10 and PCV13 serotypes are from a meta-analysis of only head-to-head studies of PCV13 vs PCV10.

Within the network meta-analyses comparisons with PCV7, GMRs favoured PCV7 over either PCV13 or PCV10 for serotypes 4, 6B, 9V, 14, and 23F. There was no difference in GMRs for Serotypes 18C and 19F across three vaccines. (Figure 2a).

There were inconsistencies between direct and indirect evidence from the network meta-analysis (p-value for inconsistency <0.05) for serotype 6B, 14, 18C and 19F (Supplementary Table 3).

At the pre-booster time point data were available from 17 cohorts. IgG responses were higher with PCV10 compared with PCV13 for all PCV7 serotypes except for serotype 14, with the point estimates of GMRs comparing PCV13 vs PCV10 ranging from 0.41 to 0.78. IgG responses were higher for PCV13 for serotypes 1, 5 and 7F. GMRs comparing PCV13 vs PCV7 showed higher IgG with PCV7 for serotypes 4, 6B, 9V, 14 and 23F, and higher IgG with PCV13 for serotype 19F (Figure 2b).

At 28 days post booster, data were available from 25 cohorts. GMRs favoured PCV13 over PCV10 for serotype 6B, 9V, 14 and 23F, and favoured PCV10 over PCV13 for serotype 18C (Figure 2c). For serotype 1, 5 and 7F, antibody responses were higher in PCV13 compared with PCV10. PCV7 recipients had higher GMCs compared with PCV13 for all PCV7 serotypes except 6B for which there was no difference, and19F, which favoured PCV13. For PCV13-only serotypes (3, 6A and 19A), GMRs favour PCV13 at all three time points. Inconsistencies were found for serotype 4 and 6B between direct and indirect evidence (Supplementary Table 3, Supplementary Figure 3).

To explore potential reasons for the observed heterogeneity, we summarised cohort-level GMRs for each vaccine comparison and present these with concomitant vaccines and vaccine schedules at all three time points in Supplementary Figure 4-42. These descriptive analyses revealed a lack of consistency in the direction of study-level estimates within each vaccine comparison, resulting in the significant heterogenicity. There was also no observable pattern in any trial level variable (region, co-administered vaccines, vaccine schedule), from which one might propose a mechanism that would adequately explain this variation in GMRs, although studies which compared vaccines with the same carrier protein seemed to have more consistent estimates. In sensitivity analysis, we restricted to 11 cohorts providing IgG results for all the three time points, and observed similar results (Supplementary Figure 43). Sensitivity analyses excluding the two studies having overall ‘high risk of bias’ did not provide different results. Excluding the study of 1+1 schedule comparing PCV10 and PCV13 did not affect the results.

### Seroefficacy

There were 12 studies (15 cohorts) with available individual participant antibody data at both post-primary and prior to the booster dose, allowing serotype-specific estimation of seroefficacy from a total of 5152 participants. Of these 15 cohorts, 6 compared PCV10 vs PCV7, 3 compared PCV13 vs PCV7 and 6 compared PCV13 vs PCV10 (Supplementary Figure 2b).

The relative risk of seroinfection from the network meta-analysis for each serotype is summarised in Figure 3 and a summary of direct and indirect evidence is given in Supplementary Figure 44. The I^2^ and p value indicate some heterogeneity for all PCV7 serotypes except for serotype 4 and 19F (Supplementary Table 4).

**Figure 3.**
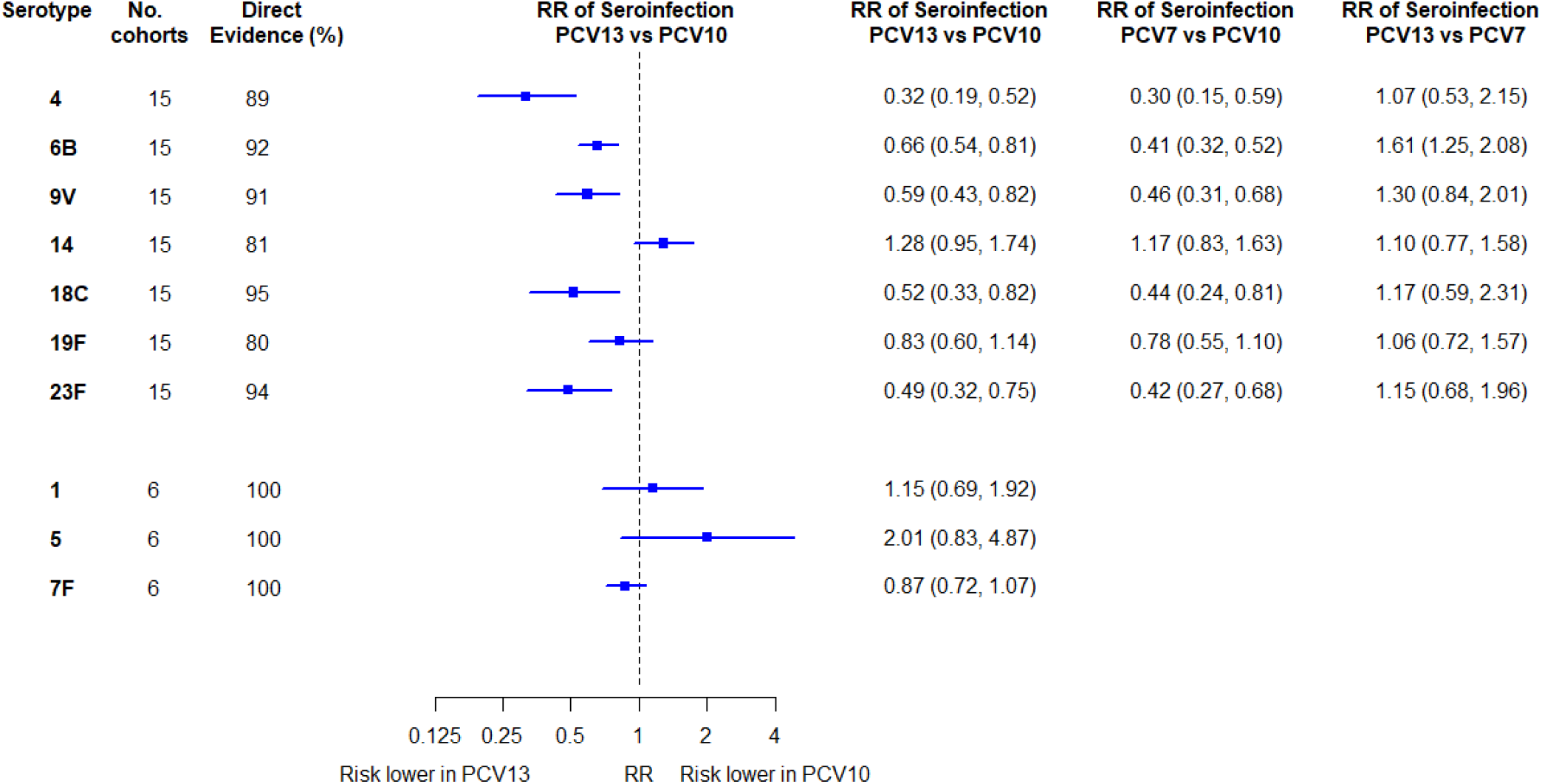
Relative risk of seroinfection from meta-analyses of head-to-head studies. RR: relative risk; PCV: Pneumococcal conjugate vaccine. Each line in the figure shows the output from a network meta-analyses (PCV7 serotypes) or direct meta-analyses (PCV10 serotypes). Blue boxes and blue lines show the point estimates and confidence intervals of relative risk of seroinfection comparing PCV13 vs PCV10. The direct evidence column shows the percentage of evidence from studies directly comparing PCV13 vs PCV10. Results for PCV10 serotypes are from a meta-analysis of only head-to-head studies of PCV13 vs PCV10, therefore estimates of PCV7 vs PCV10 and PCV13 vs PCV7 were not available.

Among PCV7 serotypes, the risk of seroinfection was lower with PCV13 than PCV10 for serotypes 4, 6B, 9V, 18C and 23F, while no difference was seen for serotype 14 and 19F (Figure 2). The RRs of seroinfection (PCV13 vs PCV10) for PCV7 serotypes ranged from 0.32 (95% CI: 0.19, 0.52) for serotype 4 to 1.28 (95% CI: 0.95, 1.74) for serotype 14. The direct evidence contributed to around 80%-95% of total evidence, and we found no inconsistency between direct and indirect evidence for all but serotype 19F (p values > 0.05, Supplementary Table 4, Supplementary Figure 45-54).

For serotypes 1, 5, and 7F, evidence was summarised from 6 studies directly comparing PCV13 with PCV10. Heterogeneity was observed for serotype 5 and all confidence intervals overlapped 1.0. Comparisons between PCV13 and PCV7 favoured neither vaccine over the other, whereas comparisons between PCV7 and PCV10 favoured PCV7 for serotypes 5, 6B, 9V, 18C, and 23F. The seroefficacy analysis results remained consistent after removing one “high risk of bias” study from the analysis.

### Association between ratios of immunogenicity and seroefficacy

Supplementary Figure 55 shows the serotype-specific relationships between immunogenicity (GMRs) and seroefficacy (RRs). Log-GMRs and log-RRs were highly or moderately correlated for all PCV7 serotypes (with weighted Pearson correlation coefficients (*ρ*) ranging from -0.76 to -0.60, all p<0.05) except for serotype 14 (*ρ* =-0.30, p=0.26).

In the combined analysis across all serotypes vaccines that produced the same amount of antibody (GMR=1) had very similar protection (adjusted RR: 0.80, 95%: CI 0.41-1.58, Figure 4). The model estimate indicates that for each two-fold increase in antibody response, the risk of seroinfection was halved (GMR of 2.0; RR 0.46, 95% CI 0.23-0.96, Figure 4a, 4b). The estimates were stable when estimates of PCV13 vs PCV7 were analysed in reverse as PCV7 vs PCV13 (GMR of 2.0; RR: 0.51, 95% CI: 0.23-1.15, Figure 4c).

**Figure 4.**
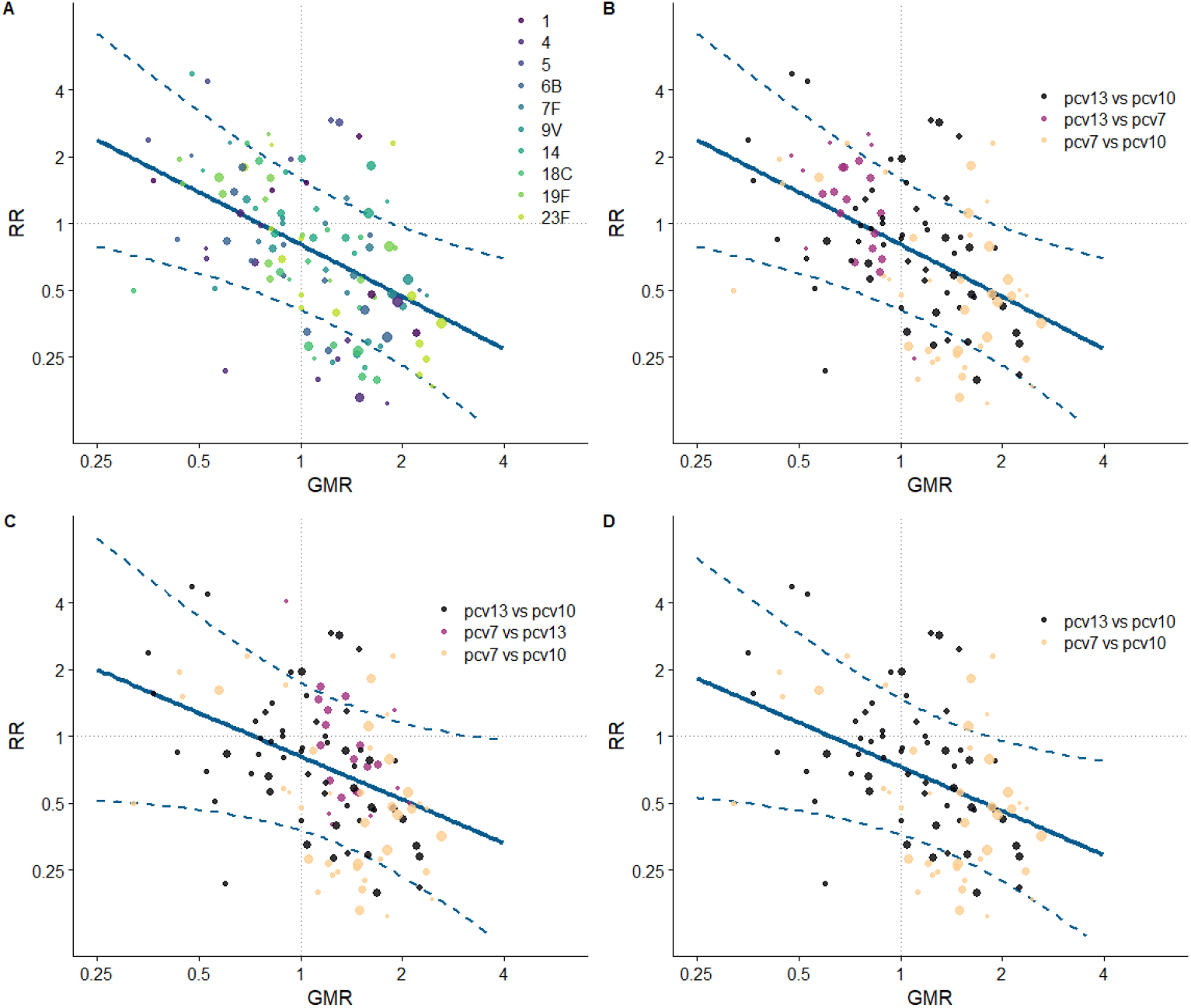
Overall association between geometric mean ratio and relative risk across all serotypes in PCV10. RR: relative risk; GMR: Geometric mean ratio; PCV: Pneumococcal conjugate vaccine. Each point shows results of a serotype specific head-to-head comparison between two vaccines from one study. Solid line shows the relationship between relative risk predicted from the model and geometric mean ratio. Dashed line shows the confidence intervals of predicted relative risk. Reference lines show geometric mean ratio equivalent to one (vertical) and relative risk equivalent to one (horizontal) which represent values associated with no difference between vaccines. Points sizes represent sample size of the trial. Panel A) shows the relationship by 13 serotypes covered by PCV13, B) shows the same data as panel A classified by vaccine comparison groups, C) shows the same data as panel B, however, studies comparing PCV13 vs PCV7 are analysed and displayed as PCV7 vs PCV13 as a sensitivity analysis D) shows a further sensitivity analysis that excludes studies of PCV13 vs PCV7 and only shows studies that compared vaccines from two different manufacturers.

When analyses were restricted to comparison between products from different manufacturers the relationship between immunogenicity and seroprotection remained similar to the main analysis with a confidence interval that incorporates 1.0 (GMR 1.0; RR: 0.73, 95% CI: 0.36-1.47) (Figure 4d).

## Discussion

In our study we used a novel methodology to define seroinfection from immunogenicity data to compare the relative efficacy of pneumococcal conjugate vaccines in preventing infection. Our analysis using individual level data from a global meta-analysis, provides the first estimates of the comparative protection afforded by different vaccines, and shows that for many serotypes, carriage events are less common after PCV13 than PCV10, likely due to a higher antibody response. In addition, we quantify the relationship between the immune response to vaccination and protection against infection, measured serologically, and show that greater protection from infection is associated with higher antibody responses in infants.

The observed heterogenicity in immunogenicity analyses was unexpected. A priori hypothesis was that if one vaccine is able to induce more antibody than another, then it would do so with some degree of consistency across all trials. However, this was not what we observed. Comparisons of the same vaccines in different studies gave widely varying estimates and although we have reported the summary GMR estimates in our immunogenicity meta-analyses, the large degree of between-study heterogeneity in these models means these overall estimates are difficult to interpret. In some settings PCV13 performed better yet in others PCV10 was the more immunogenic vaccine. Although there was no single predictor that could be identified that might explain the variation in estimates, only three candidate factors could be considered (location, schedule, and co-administered vaccines) and data reporting on co-administered vaccines were not always comprehensive. The assays used have been WHO standardised and unlikely to cause this variation, and only head-to-head studies were included.

Of note, comparisons between vaccines from the same manufacturer (PCV13 vs PCV7) were more consistent than comparisons between vaccines with different manufacturers. Immune interference (“bystander effects”) have been noted when vaccines with similar components are co-administered,^90^ and this may affect one vaccine in a comparison over another. It is interesting that 18C and 19F were serotypes that showed a very large degree of between-study heterogeneity. These two serotypes in PCV10, have different carrier proteins (18C is conjugated to tetanus toxoid and 19F is conjugated to diphtheria toxoid) and may be more susceptible than other serotypes to the influence of the type and dose of co-administered diphtheria-tetanus vaccines, and additionally, co-administration of other conjugated vaccines containing tetanus or diphtheria carrier proteins such as meningococcal vaccines. An additional potential confounder that is unmeasured in these studies, is the exposure to circulating serotypes of pneumococcus in each setting, which also has the potential to influence the immune response to vaccines. Additionally, there is growing evidence showing that antibody responses were higher in PCV10 for serotypes compared with PCV13 after the primary vaccinations of 1+1 schedule. Further studies could provide insights into the impact of vaccine schedules on protection.

These diverse immunogenicity findings from studies of the same vaccines raise the question of whether such differences in immunogenicity lead to meaningful differences in protection. If so, it may be important to know which vaccine performs better in which setting and further investigation into the predictors of the immune response to vaccines may be warranted. We addressed this question by modelling the relationship between seroefficacy estimates and immunogenicity comparisons (GMRs), analysed at the trial level across all serotypes and studies. This regression analysis did not require a meta-analysis to be performed to obtain an overall estimate of the difference between vaccines, but instead used the individual trial estimates themselves. This method capitalises on the observed between-study heterogenicity rather than being hindered by it. In our model, vaccines with higher antibody levels were also those with greater protection against subclinical infections in general. A vaccine with twice the antibody production was predicted to halve the rate at which carriage occurred. Licensure of new vaccines is based on non-inferiority comparisons with current vaccines and the proportion of antibody responses above the agreed threshold as a minimum requirement. Once a vaccine meets this “at-least-as-good-as” immunogenicity criteria, it has previously not been clear whether exceeding it is of benefit, and the WHO position paper states “*It is unknown whether a lower serotype-specific GMC of antibody indicates less efficacy*”.^6^ Our results show that lower protection against subclinical infection does indeed follow from lower antibody production, and that two vaccines that produce a similar level of antibody will provide similar levels of protection, even if they are from different manufacturers.

The implications of these findings are of greatest importance when a new vaccine rollout is being considered. Lower antibody production or lower seroefficacy for one vaccine product does not necessarily imply limited effectiveness against invasive pneumococcal diseases when considering vaccines such as PCV10 and PCV13 which have been shown to be highly effective vaccines in many settings. Instead, lower antibody responses that resulted in lower protection against carriage would lead to less rapidly observed indirect protection after implementation into a national programme as a smaller proportion of transmission events would be blocked by the vaccine. For serotypes where protective impact has not been observed (serotype 3), new vaccines with substantially higher antibody responses may be needed. A phase II clinical trial of PCV15 compared with PCV13 reported almost twice the antibody level for serotype 3 at 28 days post-primary series for PCV15 (GMR 1.93, 95% CI: 1.71, 2.18).^91^ Based on our modelled association between GMR and RR, the relative risk of seroinfection with PCV15 versus PCV13 was estimated to be 0.48. Previously reported vaccine effectiveness estimates for serotype 3 included -27% (95% CI: -180, 44) and 1% (95% CI: -106, 52) against nasopharyngeal carriage ^30,92^, and these translate to point estimates of 39% (95% CI: -16%, 66%) and 52% (95% CI: 9%, 79%) vaccine effectiveness against carriage of this serotype with PCV15 based on the relationship: (VE_(pcv15)_=(1-RR_(pcv15 vs pcv13)_*(1-VE_(pcv13)_/100%))*100%).

This evidence of differences in serotype-specific protection can be incorporated into cost-effectiveness models used to compare vaccine products.^93^ Cost-effectiveness studies have highlighted the lack of head-to-head evidence of efficacy for different PCVs, resulting in cost-effectiveness models that ignore serotype-specific differences and assume equivalent efficacy for different PCVs. ^94,95,96^ Our study fills this evidence gap and allows researchers and policy-makers to use more accurate vaccine specific models in decision-making.

Seroinfection data can be feasibly collected in a randomised controlled trial and used as a surrogate outcome for estimating vaccine efficacy against carriage. Antibody levels in children are highest when measured approximately one-month after their primary series of vaccines and decline sharply in the subsequent months. Instances of increase in antibody between primary and booster doses can therefore be assumed to be indicative of nasopharyngeal colonisation, an assumption supported by data from challenge studies. There is substantial evidence from pneumococcal challenge studies that participants exposed (“challenged”) with pneumococcus who go on to develop an established carriage infection experience significant increases in antibody post-exposure, whereas those who remain carriage negative do not.^97-99^ We previously observed in studies in Nepal that the serotypes commonly detected in seroinfection data from a clinical trial were closely aligned with serotypes that were circulating in the community at the same time, as measured through a separate cross-sectional nasopharyngeal carriage study, further validating the robustness of this approach.^12^ Cross-sectional carriage studies using nasopharyngeal swabs are susceptible to misclassification bias when the time of sampling is not at the exact time of peak infection resulting in a negative swab. Using seroinfection as an outcome reduces this type of bias as the antibody response to carriage persists for a longer period of time than the carriage event. Nevertheless, misclassification bias can exist if antibody wanes quickly following infection, which may bias the RR estimates to the null.

Seroefficacy analyses need to be restricted to serotypes contained in both vaccines. Comparing a vaccinated cohort to a cohort that is unvaccinated, or receives a vaccine that does not contain the serotype of interest, will result in biased estimates as the immune response after exposure to a pathogen will differ in children whose immune system is primed for that pathogen, when compared with a naïve population. For this reason, we restricted our seroefficacy analysis to shared serotypes between vaccines. Whilst seroinfection is most likely an indicator of nasophryngeal carriage, it may also represent cases of asymptomatic bacteremia.

In conclusion, we estimated serotype-specific difference in both seroefficacy and immunogenicity between PCV10 and PCV13. Higher IgG antibody levels confer better protection against seroinfection. This methodology can be further used to compare novel high-valent PCVs and to inform cost-effectiveness models.

## Data Availability

This publication is based on research using data from data contributor Pfizer that has been made available through Vivli, Inc and data contributor GSK that has been made available through clinicalstudydatarequest.com. Vivli and other third parties have not contributed to or approved, and are not in any way responsible for, the contents of this publication. Data were made available for a limited period of time to conduct these analyses. The authors do not have continuing access to the datasets.

## Contributors

MV conceived and designed the study and obtained funding. SF, JM, NR, JH and MV contributed to the methods of the study. NR devised and conducted the search strategy. JM and NP conducted the study selection and assessment of bias. SF and MV obtained the individual participant data and extracted the aggregated data. SF and MV had full access to all study data and analysed the data. SF wrote the first draft of the manuscript. All authors interpreted the data and contributed to the writing of the final version of the manuscript.

