## Supplementary Material for "Immunogenicity and seroefficacy of pneumococcal conjugate vaccines – a systematic review and network meta-analysis"

**Supplementary file**

Supplementary Figure 14. Trial level geometric mean ratios for serotypes 3 at post-primary vaccination series. ... 37
Supplementary Figure 15. Trial level geometric mean ratios for serotypes 6A at post-primary vaccination series. ... 38
Supplementary Figure 16. Trial level geometric mean ratios for serotypes 19A at post-primary vaccination series. ... 39
Supplementary Figure 17. Trial level geometric mean ratios for serotypes 4 at pre-booster. ... 40
Supplementary Figure 18. Trial level geometric mean ratios for serotypes 6B at pre-booster. ... 41
Supplementary Figure 19. Trial level geometric mean ratios for serotypes 9V at pre-booster. ... 42
Supplementary Figure 20. Trial level geometric mean ratios for serotypes 14 at pre-booster. ... 43
Supplementary Figure 21. Trial level geometric mean ratios for serotypes 18C at pre-booster. ... 44
Supplementary Figure 22. Trial level geometric mean ratios for serotypes 19F at pre-booster. ... 45
Supplementary Figure 23. Trial level geometric mean ratios for serotypes 23F at pre-booster. ... 46
Supplementary Figure 24. Trial level geometric mean ratios for serotypes 1 at pre-booster. ... 47
Supplementary Figure 25. Trial level geometric mean ratios for serotypes 5 at pre-booster. ... 48
Supplementary Figure 26. Trial level geometric mean ratios for serotypes 7F at pre-booster. ... 49
Supplementary Figure 27. Trial level geometric mean ratios for serotypes 3 at pre-booster. ... 50
Supplementary Figure 28. Trial level geometric mean ratios for serotypes 6A at pre-booster. ... 51
Supplementary Figure 29. Trial level geometric mean ratios for serotypes 19A at pre-booster. ... 52
Supplementary Figure 30. Trial level geometric mean ratios for serotypes 4 at post-booster. ... 53
Supplementary Figure 31. Trial level geometric mean ratios for serotypes 6B at post-booster. ... 54
Supplementary Figure 32. Trial level geometric mean ratios for serotypes 9V at post-booster. ... 55
Supplementary Figure 33. Trial level geometric mean ratios for serotypes 14 at post-booster. ... 56
Supplementary Figure 34. Trial level geometric mean ratios for serotypes 18C at post-booster. ... 57
Supplementary Figure 35. Trial level geometric mean ratios for serotypes 19F at post-booster. ... 58
Supplementary Figure 36. Trial level geometric mean ratios for serotypes 23F at post-booster. ... 59
Supplementary Figure 37. Trial level geometric mean ratios for serotypes 1 at post-booster. ... 60
Supplementary Figure 38. Trial level geometric mean ratios for serotypes 5 at post-booster. ... 61
Supplementary Figure 39. Trial level geometric mean ratios for serotypes 7F at post-booster. ... 62
Supplementary Figure 40. Trial level geometric mean ratios for serotypes 3 at post-booster. ... 63
Supplementary Figure 41. Trial level geometric mean ratios for serotypes 6A at post-booster. ... 64
Supplementary Figure 42. Trial level geometric mean ratios for serotypes 19A at post-booster. ... 65
Supplementary Figure 43. Sensitivity analysis on geometric mean ratios comparing PCV13 vs PCV10, PCV7 vs PCV10, and PCV13 vs PCV7 at a) post-primary vaccination series, b) pre-boost, and c) post-boost by including studies providing data for all three time points. ... 66
Supplementary Figure 44. Direct and indirect evidence on relative risk comparing PCV13 vs PCV10 for serotypes in PCV7 (4, 6B, 9V, 14, 18C, 19F and 23F) ... 69
Supplementary Figure 45. Trial level relative risk for serotype 4. ... 70
Supplementary Figure 46. Trial level relative risk for serotype 6B. ... 71

### Methods

Our systematic review protocol was registered with PROSPERO (International Prospective Register of Systematic Reviews). Registration number: CRD42019124580.

#### Search strategy

The search strategy was devised and conducted by an information specialist (NR). Five databases and two trial registers were searched from database inception to 27th July 2022. The original search was run in June 2019, with an update search run in July 2022. The databases searched were Cochrane Database of Systematic Reviews & Cochrane Central Register of Controlled Trials (Cochrane Library, Wiley)[Issue 7 of 12, July 2022], Embase (OvidSP)[1974-present], Global Health (OvidSP)[1973 to 2022 Week 29] and Medline (OvidSP)[1946-present]. The trial registers searched were ClinicalTrials.gov (https://clinicaltrials.gov/) and WHO International Clinical Trials Registry Platform (https://trialsearch.who.int/). The search comprised of title/abstract keywords and subject headings for pneumococcal vaccines and children. A methodological search filter for randomised controlled trials taken from the Cochrane Handbook was used to limit to RCTs.[1,2] Pharmaceutical company websites (GSK and Pfizer) were also hand searched for relevant studies. A full list of search terms for each database is summarised in Supplementary Table 1. No date or language limits were applied. References were exported to Endnote 20 for de-duplication.

#### Study selection

Two reviewers (JM, NP) independently reviewed the title and abstract of each reference and identified potentially relevant references. Two reviewers (JM, NP) independently selected studies to be included in the review from retrieved full-text papers using predetermined inclusion criteria. Disagreements about study inclusion were resolved by a third reviewer (MV).

Randomised controlled trials were included if they provided head-to-head comparisons of either PCV7, PCV10, or PCV13 among infants and children less than 2 years of age, and if they provided estimates on antibody responses (serotype-specific anti-pneumococcal IgG) to PCVs for at least one time point of 1) between 4 and 6 weeks after the primary vaccination series, and/or one-month after a booster vaccination. Trials were eligible only if they included at least one of the three currently licensed (PCV10 and 13) or previously licensed (PCV7) vaccines.

Trials were excluded if they did not contain a head-to-head randomised comparison of eligible vaccines, contained only a single vaccine, or enrolled immuno-compromised (e.g. HIV) children.

#### PCVs in systematic review

1. 7-valent pneumococcal conjugate vaccine (PCV7: Prevnar, Pfizer), containing serotypes 4, 6B, 9V, 14, 18C, 19F, and 23F, each conjugated to diphtheria cross-reacting material (CRM).
2. 13-valent pneumococcal conjugate vaccine (PCV13: Prevenar 13, Pfizer), containing serotypes 1, 3, 4, 5, 6A, 6B, 7F, 9V, 14, 18C, 19A, 19F, and 23F, each conjugated to diphtheria cross-reacting material (CRM).
3. 10-valent pneumococcal conjugate vaccine (PCV10: Synflorix, GlaxoSmithKline), containing serotypes 1, 4, 5, 6B, 7F, 9V, 14, 18C, 19F, and 23F, conjugated to non-typeable Haemophilus influenzae protein D, for 8 serotypes, or tetanus or diphtheria protein (serotypes 18C and 19F respectively).

#### Data retrieval

For all eligible trials, the publication authors/data owners were approached for trial and individual participant level data. Baseline characteristics and potential effect modifiers were extracted on participants' age, sex, country, immunogenicity assays, co-administered study vaccines and vaccine schedules. The following study-level data were extracted from trial registries/published studies:

- trial registration number/study identifier
- study country
- PCV vaccination schedule (e.g. 2+1, 3+1, 3+0)

Individual participant level data were retrieved if available for variables below.

- vaccines administered (both study vaccines and vaccines administered concomitantly as part of the routine immunisation schedule)

- vaccination dates
- details of laboratory assays conducted, including where assays were run, units of measurement, and the lower limit of quantification
- participants' age at enrolment
- participants' sex
- serotype-specific anti-pneumococcal IgG measured by ELISA at all time-points

Aggregate data from publications were extracted if individual participant data were not available. Data extraction of published results and individual participant level data were independently completed by SF and MV.

#### Assessment of risk of bias in included studies

Risk of bias in results of the included studies was assessed independently by two reviewers (JM, NP) using the Cochrane Risk of Bias Tool (RoB 2).[3] This considers the risk of bias in five domains (randomisation process, deviations from the intended interventions, missing outcome data, measurement of the outcome, selection of the reported result) and generates an overall risk of bias. Assessments were undertaken for immunogenicity of PCV7, PCV10 and PCV13 for each serotype contained in the vaccines. Risks of bias for sero-efficacy outcomes are assumed to be identical because the data came from the same blood samples and were analysed in similar ways. The possible risk of bias judgments for each domain, and overall, are 'low risk of bias', 'some concerns' and 'high risk of bias'. Disagreements between reviewers were resolved by consensus. Results for the risk of bias assessment were presented using robvis (visualisation tool).[4]

#### Assessment of heterogeneity and inconsistency of network meta-analysis

To assess the statistical heterogeneity and inconsistency of NMA, we evaluated the transitivity assumption by visually comparing the distribution of the baseline characteristics and potential effect modifiers across the different pairwise comparisons. We assessed the presence of heterogeneity using estimated values of the heterogeneity variance parameters ($\tau 2$) and the I-squared statistic and its 95% confidence interval that measures the percentage of variability in point estimates that cannot be attributed to random error. We evaluated the inconsistency, i.e. coherence between direct and indirect evidence, using a Q statistic,[5] which measures the deviation from consistency. The random-effects model was fitted following the graph-theoretical approach and using the GMR and RR as effect estimate with 95% CI.[6]

Some individual participant level data were missing due to laboratory errors, insufficient blood sample volume or participant withdrawal. Data were not imputed and missing data were considered missing-completely-at-random. Individual participant level data were analysed according to the vaccine received.

**Supplementary Table 1. Full search terms for databases.**

| **Medline (Ovid MEDLINE® Epub Ahead of Print, In-Process & Other Non-Indexed Citations, Ovid MEDLINE® Daily and Ovid MEDLINE®) 1946 to present** |
|---|
| Streptococcus pneumoniae/ |
| (pneumococc* or s pneumoniae or strep* pneumoniae or strep* p).ti,ab. |
| 1 or 2 |
| vaccines/ or bacterial vaccines/ or streptococcal vaccines/ or Vaccines, Conjugate/ |
| (vaccin* or immuni?ation? or immuni?e? or inoculat* or conjugate).ti. |
| (conjugate adj2 vaccin*).ti,ab. |
| (7 valent or 7valent or seven valent or heptavalent or hepta-valent).ti,ab. |
| (9 valent or 9valent or nine valent or nonavalent or nona-valent).ti,ab. |
| (10 valent or 10valent or ten valent or decavalent or deca-valent).ti,ab. |
| (13 valent or 13valent or thirteen valent).ti,ab. |
| 4 or 5 or 6 or 7 or 8 or 9 or 10 |
| 3 and 11 |
| exp Pneumococcal Vaccines/ |
| ((pneumococc* or s pneumoniae or strep* pneumoniae or strep* p) adj5 (vaccin* or immuni?ation? or immuni?e? or inoculat* or conjugate)).ti,ab. |
| (pcv7 or pcv 7 or pncrm* or pnccrm* or 7vpnc or 7vcrm).ti,ab. |
| (pcv9 or pcv 9).ti,ab. |
| (pcv10 or pcv 10 or phidcv or phid cv).ti,ab. |
| (pcv13 or pcv 13 or 13vcrm).ti,ab. |
| (pneumovax or pneumopur or streptopur or streptorix or prevnar or prevenar or synflorix or gsk 1024850a or gsk1024850a).ti,ab. |
| 12 or 13 or 14 or 15 or 16 or 17 or 18 or 19 |
| exp child/ or infant/ |
| (child* or infan* or baby or babies or toddler? or preschool* or pre-school* or p?ediatric?).ti,ab. |
| 21 or 22 |
| 20 and 23 |
| randomized controlled trial.pt. |
| controlled clinical trial.pt. |
| randomized.ab. |
| placebo.ab. |

| |
|---|
| drug therapy.fs. |
| randomly.ab. |
| trial.ab. |
| groups.ab. |
| 25 or 26 or 27 or 28 or 29 or 30 or 31 or 32 |
| exp animals/ not humans.sh. |
| 33 not 34 |
| 24 and 35 |
| limit 24 to ("reviews (maximizes specificity)" or "systematic review") |
| 36 or 37 |
| (2019* or 2020* or 2021* or 2022*).ed,ez,yr. |
| 38 and 39 |
| **Embase 1974 to present** |
| Streptococcus pneumoniae/ |
| (pneumococc* or s pneumoniae or strep* pneumoniae or strep* p).ti,ab. |
| 1 or 2 |
| vaccine/ or bacterial vaccine/ or streptococcus vaccine/ |
| (vaccin* or immuni?ation? or immuni?e? or inoculat* or conjugate).ti. |
| (conjugate adj2 vaccin*).ti,ab. |
| (7 valent or 7valent or seven valent or heptavalent or hepta-valent).ti,ab. |
| (9 valent or 9valent or nine valent or nonavalent or nona-valent).ti,ab. |
| (10 valent or 10valent or ten valent or decavalent or deca-valent).ti,ab. |
| (13 valent or 13valent or thirteen valent).ti,ab. |
| 4 or 5 or 6 or 7 or 8 or 9 or 10 |
| 3 and 11 |
| pneumococcus vaccine/ |
| ((pneumococc* or s pneumoniae or strep* pneumoniae or strep* p) adj5 (vaccin* or immuni?ation? or immuni?e? or inoculat* or conjugate)).ti,ab. |
| (pcv7 or pcv 7 or pncrm* or pnccrm* or 7vpnc or 7vcrm).ti,ab. |
| (pcv9 or pcv 9).ti,ab. |
| (pcv10 or pcv 10 or phidcv or phid cv).ti,ab. |
| (pcv13 or pcv 13 or 13vcrm).ti,ab. |

| |
|---|
| (pneumovax or pneumopur or streptopur or streptorix or prevnar or prevenar or synflorix or gsk 1024850a or gsk1024850a).ti,ab. |
| 12 or 13 or 14 or 15 or 16 or 17 or 18 or 19 |
| exp child/ |
| (child* or infan* or baby or babies or toddler? or preschool* or pre-school* or p?ediatric?).ti,ab. |
| 21 or 22 |
| 20 and 23 |
| randomized controlled trial/ |
| single blind procedure/ or double blind procedure/ |
| crossover procedure/ |
| random*.tw. |
| (((singl* or doubl*) adj (blind* or mask*)) or crossover or cross over or factorial* or latin square or assign* or allocat* or volunteer*).ti,ab. |
| 25 or 26 or 27 or 28 or 29 |
| (exp animals/ or nonhuman/) not human/ |
| 30 not 31 |
| 24 and 32 |
| limit 24 to ("systematic review" or "reviews (maximizes specificity)") |
| 33 or 34 |
| (2019* or 2020* or 2021* or 2022*).yr,dd,dc. |
| 35 and 36 |
| **Global Health <1973 to 2022 Week 29>** |
| streptococcus pneumoniae/ |
| (pneumococc* or s pneumoniae or strep* pneumoniae or strep* p).ti,ab. |
| 1 or 2 |
| vaccines/ or conjugate vaccines/ |
| (vaccin* or immuni?ation? or immuni?e? or inoculat* or conjugate).ti. |
| (conjugate adj2 vaccin*).ti,ab. |
| (7 valent or 7valent or seven valent or heptavalent or hepta-valent).ti,ab. |
| (9 valent or 9valent or nine valent or nonavalent or nona-valent).ti,ab. |
| (10 valent or 10valent or ten valent or decavalent or deca-valent).ti,ab. |
| (13 valent or 13valent or thirteen valent).ti,ab. |
| 4 or 5 or 6 or 7 or 8 or 9 or 10 |

| |
|---|
| 3 and 11 |
| ((pneumococc* or s pneumoniae or strep* pneumoniae or strep* p) adj5 (vaccin* or immuni?ation? or immuni?e? or inoculat* or conjugate)).ti,ab. |
| (pcv7 or pcv 7 or pncrm* or pnccrm* or 7vpnc or 7vcrm).ti,ab. |
| (pcv9 or pcv 9).ti,ab. |
| (pcv10 or pcv 10 or phidcv or phid cv).ti,ab. |
| (pcv13 or pcv 13 or 13vcrm).ti,ab. |
| (pneumovax or pneumopur or streptopur or streptorix or prevnar or prevenar or synflorix or gsk 1024850a or gsk1024850a).ti,ab. |
| 12 or 13 or 14 or 15 or 16 or 17 or 18 |
| exp children/ or infants/ |
| (child* or infan* or baby or babies or toddler? or preschool* or pre-school* or p?ediatric?).ti,ab. |
| 20 or 21 |
| 19 and 22 |
| (random* or blind* or allocat* or assign* or trial* or placebo* or crossover* or cross-over*).mp. |
| 23 and 24 |
| (2019* or 2020* or 2021* or 2022*).yr. |
| 25 and 26 |
| **ClinicalTrials.gov – 1/6/2019** |
| (pneumococcal OR pneumococcus OR "streptococcus pneumoniae" OR "streptococcal pneumoniae" OR "streptococcus p" OR "streptococcal p") AND (vaccine OR vaccines OR vaccination OR immunisation OR immunization OR immunise OR immunisation OR inoculate) \| Child |
| (pneumococcal OR pneumococcus OR "streptococcus pneumoniae" OR "streptococcal pneumoniae" OR "streptococcus p" OR "sterptococcal p") AND (conjugate OR valent) \| Child |
| pcv7 or pcv 7 or pncrm or pnccrm or 7vpnc or 7vcrm or pcv9 or pcv 9 or pcv10 or pcv 10 or phidcv or phid cv or pcv13 or pcv 13 or 13vcrm |
| pneumovax or pneumopur or streptopur or streptorix or prevnar or prevenar or synflorix or gsk 1024850a or gsk1024850a \| Child |
| **ClinicalTrials.gov - 2022 - Added since 01/06/2019** |
| Other terms=(pneumococcal OR pneumococcus OR "streptococcus pneumoniae" OR "streptococcal pneumoniae" OR "streptococcus p" OR "streptococcal p") AND (vaccine OR vaccines OR vaccination OR immunisation OR immunization OR immunise OR immunisation OR inoculate) \| Child \| First posted from 06/01/2019 to 01/01/2024 |
| Title=(pneumococcal OR pneumococcus OR "streptococcus pneumoniae" OR "streptococcal pneumoniae" OR "streptococcus p" OR "streptococcal p") AND (vaccine OR vaccines OR vaccination OR immunisation OR immunization OR immunise OR immunisation OR inoculate) \| Child \| First posted from 06/01/2019 to 01/01/2024 |

| |
|---|
| Condition=(pneumococcal OR pneumococcus OR "streptococcus pneumoniae" OR "streptococcal pneumoniae" OR "streptococcus p" OR "streptococcal p") AND Intervention=(vaccine OR vaccines OR vaccination OR immunisation OR immunization OR immunise OR immunisation OR inoculate) \| Child \| First posted from 06/01/2019 to 01/01/2024 |
| Other terms=(pneumococcal OR pneumococcus OR "streptococcus pneumoniae" OR "streptococcal pneumoniae" OR "streptococcus p" OR "streptococcal p") AND (conjugate OR valent) \| Child |
| Title=(pneumococcal OR pneumococcus OR "streptococcus pneumoniae" OR "streptococcal pneumoniae" OR "streptococcus p" OR "streptococcal p") AND (conjugate OR valent) \| Child |
| Condition=(pneumococcal OR pneumococcus OR "streptococcus pneumoniae" OR "streptococcal pneumoniae" OR "streptococcus p" OR "streptococcal p") AND Intervention=(conjugate OR valent) \| Child |
| Other terms=pcv7 or pcv 7 or pncrm or pnccrm or 7vpnc or 7vcrm or pcv9 or pcv 9 or pcv10 or pcv 10 or phidcv or phid cv or pcv13 or pcv 13 or 13vcrm \| Child |
| Title=pcv7 or pcv 7 or pncrm or pnccrm or 7vpnc or 7vcrm or pcv9 or pcv 9 or pcv10 or pcv 10 or phidcv or phid cv or pcv13 or pcv 13 or 13vcrm \| Child |
| Intervention=pcv7 or pcv 7 or pncrm or pnccrm or 7vpnc or 7vcrm or pcv9 or pcv 9 or pcv10 or pcv 10 or phidcv or phid cv or pcv13 or pcv 13 or 13vcrm \| Child |
| Other terms=pneumovax or pneumopur or streptopur or streptorix or prevnar or prevenar or synflorix or gsk 1024850a or gsk1024850a \| Child |
| Title=pneumovax or pneumopur or streptopur or streptorix or prevnar or prevenar or synflorix or gsk 1024850a or gsk1024850a \| Child |
| Intervention=pneumovax or pneumopur or streptopur or streptorix or prevnar or prevenar or synflorix or gsk 1024850a or \| Child |
| **WHO ICTRP** |
| pneumococcal vaccine OR pneumococcus vaccine OR pneumococcal conjugate OR pneumococcus conjugate OR pcv OR pneumovax OR pneumopur OR streptopur OR streptorix OR prevnar OR prevenar OR synflorix OR gsk 1024850a OR gsk1024850a - TRIALS IN CHILDREN |
| **Cochrane (CDSR - limited to Publication date: 01/06/2019-27/07/2022, CENTRAL - limited to Added to database date: 01/06/2019-27/07/2022)** |

| ID | Search |
|---|---|
| #1 | MeSH descriptor: [Streptococcus pneumoniae] explode all trees |
| #2 | (pneumococc* or "s pneumoniae" or "streptococcus pneumoniae" or "streptococcus p" or "streptococcal pneumoniae" or "streptococcal p") |
| #3 | #1 or #2 |
| #4 | MeSH descriptor: [Vaccines] this term only |
| #5 | MeSH descriptor: [Bacterial Vaccines] this term only |
| #6 | ((vaccin* or immunization* or immunize* or immunisation* or immunise* or inoculat* or conjugate or valent)):ti,ab,kw |
| #7 | #4 or #5 or #6 |
| #8 | #3 and #7 |
| #9 | MeSH descriptor: [Pneumococcal Vaccines] explode all trees |

| | |
|---|---|
| #10 | ((pcv7 or "pcv 7" or pncrm* or pnccrm* or 7vpnc or 7vcrm)):ti,ab,kw |
| #11 | ((pcv9 or "pcv 9")):ti,ab,kw |
| #12 | (pcv10 or" pcv 10" or phidcv or "phid cv"):ti,ab,kw |
| #13 | (pcv13 or "pcv 13" or 13vcrm):ti,ab,kw |
| #14 | pneumovax or pneumopur or streptopur or streptorix or prevnar or prevenar or synflorix or gsk 1024850a or gsk1024850a |
| #15 | #8 or #9 or #10 or #11 or #13 or #14 |
| #16 | MeSH descriptor: [Child] explode all trees |
| #17 | MeSH descriptor: [Infant] this term only |
| #18 | (child* or infan* or baby or babies or toddler* or preschool* or pre-school* or pediatric* or paediatric*):ti,ab,kw |
| #19 | #16 or #17 or #18 |
| #20 | #15 and #19 |

**Supplementary Table 2. Summary of eligible studies excluded from network meta-analysis due to either no closed loop or data unavailable.**

| | Author & Year* | NCT | Comparison |
|---|---|---|---|
| **Studies with no closed loop** | Trial register[7] | NCT00169481 | pcv7 vs pcv11 |
| | Dagan et al. 1996[8] | Not found | pcv7 vs ppv23 |
| | Greenberg et al. 2018[9] | NCT01215188 | pcv13 vs pcv15 |
| | Rupp et al. 2019[10] | NCT0251373<br>NCT02037984 | pcv13 vs pcv15 |
| | Thisyakorn et al. 2014[11] | NCT00594347 | pcv7 vs ppv23 |
| | Martinez et al. 2018[12] | RPCE00000173 | Cuban pcv7 vs pcv10 |
| | Platt et al. 2020[13] | NCT02987972 | pcv15 vs pcv13 |
| | Bili et al. 2021[14] | NCT03620162 | pcv15 vs pcv13 |
| | Shin et al. 2020[15] | NCTR20170109002 | pcv12 vs pcv13 |
| | Chen et al. 2016[16];<br>Zhao et al. 2022[17] | NCT02736240 | pcv7 vs Chinese pcv13 |
| | Senders et al. 2021[18];<br>Senders et al. 2020[19] | NCT03512288 | pcv13 vs pcv20 |
| | Banniettis et al. 2021[20] | NCT03885934 | pcv15 vs pcv13 |
| **Studies with data unavailable** | Martinon-Torres et al. 2012[21] | NCT00474539 | pcv7 vs pcv13 |
| | Vanderkooi et al. 2012[22] | NCT00475033 | pcv7 vs pcv13 |
| | De Los Santos et al. 2017[23] | NCT01641133 | pcv10 vs pcv13 |
| | Diez-Domingo et al. 2013[24] | NCT00368966 | pcv7 vs pcv13 |
| | Clarke et al. 2020[25]* | NCT02308540 | pcv10-SII vs pcv13 |
| | Clarke et al. 2021[26]* | NCT03197376 | pcv10 vs pcv10-SII |

* Individual participant data unavailable

PCV – Pneumococcal conjugate vaccine; PPSV - Pneumococcal polysaccharide vaccine; PCV10-SII – 10-valent Pneumococcal conjugate vaccine by Serum Institute of India

**Supplementary Table 3. Summary of the statistical heterogeneity and incoherence for immunogenicity analyses shown in Figure 2**

| Serotype | Time point | No. of study | No. and proportion of study providing direct evidence | | No. of participants providing direct evidence | No. of participants providing indirect evidence | tau | $I^2$ (%) | p-value (Q of heterogeneity and inconsistency; df) | p-value (Q of heterogeneity; df) | p-value (Q of inconsistency; df) |
|---|---|---|---|---|---|---|---|---|---|---|---|
| **4** | post prime | 28 | 9 | 47.9% | 2,356 | 9,258 | 0.089 | 86.8 (82.0-90.4) | 0.000 (197.440;26) | 0.000 (196.733;25) | 0.400 (0.707;1) |
| **6B** | post prime | 28 | 9 | 59.7% | 2,356 | 9,194 | 0.109 | 82.2 (75.0-87.3) | 0.000 (145.913;26) | 0.000 (108.043;25) | 0.000 (37.870;1) |
| **9V** | post prime | 28 | 9 | 48.0% | 2,360 | 9,241 | 0.078 | 83.8 (77.5-88.4) | 0.000 (160.708;26) | 0.000 (158.678;25) | 0.154 (2.030;1) |
| **14** | post prime | 28 | 9 | 48.6% | 2,358 | 9,186 | 0.069 | 71.1 (57.4-80.4) | 0.000 (89.983;26) | 0.000 (83.435;25) | 0.010 (6.548;1) |
| **18C** | post prime | 28 | 9 | 50.9% | 2,358 | 9,258 | 0.147 | 93.7 (91.9-95.1) | 0.000 (413.924;26) | 0.000 (405.782;25) | 0.004 (8.142;1) |
| **19F** | post prime | 28 | 9 | 51.8% | 2,358 | 9,235 | 0.199 | 96.2 (95.3-96.9) | 0.000 (679.122;26) | 0.000 (675.147;25) | 0.046 (3.975;1) |
| **23F** | post prime | 28 | 9 | 55.1% | 2,358 | 9,211 | 0.095 | 80.6 (72.5-86.3) | 0.000 (133.979;26) | 0.000 (131.123;25) | 0.091 (2.856;1) |
| **1** | post prime | 9 | 9 | 100% | 2,357 | | 0.177 | 93.8 (90.4-96.1) | 0.000 (130.029;8) | | |
| **5** | post prime | 9 | 9 | 100% | 2,356 | | 0.193 | 95.1 (92.5-96.7) | 0.000 (161.837;8) | | |
| **7F** | post prime | 9 | 9 | 100% | 2,358 | | 0.056 | 63.6 (25.3-82.3) | 0.005 (21.980;8) | | |

| | | | | | | | | | | | |
|---|---|---|---|---|---|---|---|---|---|---|---|
| **3** | post prime | 9 | 9 | 100% | 2,354 | | 0.495 | 99.2 (99.0-99.4) | 0.000 (1006.362;8) | | |
| **6A** | post prime | 9 | 9 | 100% | 2,354 | | 0.514 | 99.0 (98.8-99.2) | 0.000 (825.778;8) | | |
| **19A** | post prime | 9 | 9 | 100% | 2,356 | | 0.381 | 98.1 (97.5-98.6) | 0.000 (428.259;8) | | |
| **4** | prior booster | 17 | 7 | 68.9% | 1,816 | 4,174 | 0.098 | 83.7 (74.8-89.4) | 0.000 (92.071;15) | 0.000 (90.267;14) | 0.179 (1.805;1) |
| **6B** | prior booster | 17 | 7 | 74.3% | 1,812 | 4,140 | 0.141 | 87.7 (81.7-91.8) | 0.000 (122.370;15) | 0.000 (112.127;14) | 0.001 (10.242;1) |
| **9V** | prior booster | 17 | 7 | 68.1% | 1,810 | 4,175 | 0.112 | 88.3 (82.6-92.1) | 0.000 (127.983;15) | 0.000 (127.962;14) | 0.885 (0.021;1) |
| **14** | prior booster | 17 | 7 | 64.9% | 1,816 | 4,178 | 0.044 | 39.5 (0-66.6) | 0.053 (24.789;15) | 0.045 (24.090;14) | 0.403 (0.699;1) |
| **18C** | prior booster | 17 | 7 | 65.6% | 1,806 | 4,177 | 0.089 | 82.0 (71.8-88.5) | 0.000 (83.259;15) | 0.000 (68.689;14) | 0.000 (14.571;1) |
| **19F** | prior booster | 17 | 7 | 78.9% | 1,807 | 4,151 | 0.128 | 84.2 (75.6-89.7) | 0.000 (94.673;15) | 0.000 (69.142;14) | 0.000 (25.531;1) |
| **23F** | prior booster | 17 | 7 | 72.9% | 1,808 | 4,170 | 0.104 | 79.0 (66.5-86.8) | 0.000 (71.375;15) | 0.000 (68.632;14) | 0.098 (2.742;1) |
| **1** | prior booster | 7 | 7 | 100% | 1,820 | | 0.121 | 87.9 (77.5-93.5) | 0.000 (49.663;6) | | |
| **5** | prior booster | 7 | 7 | 100% | 1,813 | | 0.090 | 80.3 (59.9-90.3) | 0.000 (30.418;6) | | |
| **7F** | prior booster | 7 | 7 | 100% | 1,812 | | 0.075 | 73.7 (43.6-87.7) | 0.001 (22.787;6) | | |

| | | | | | | | | | | | |
|---|---|---|---|---|---|---|---|---|---|---|---|
| **3** | prior booster | 7 | 7 | 100% | 1,813 | | 0.236 | 95.0 (91.8-96.9) | 0.000 (119.248;6) | | |
| **6A** | prior booster | 7 | 7 | 100% | 1,805 | | 0.191 | 93.2 (88.4-96.0) | 0.000 (87.690;6) | | |
| **19A** | prior booster | 7 | 7 | 100% | 1,811 | | 0.115 | 79.2 (57.2-89.8) | 0.000 (28.782;6) | | |
| **4** | post booster | 25 | 6 | 53.1% | 1,706 | 8,457 | 0.063 | 72.9 (59.3-81.9) | 0.000 (84.813;23) | 0.000 (75.091;22) | 0.002 (9.722;1) |
| **6B** | post booster | 25 | 6 | 53.1% | 1,707 | 8,434 | 0.088 | 80.1 (71.1-86.3) | 0.000 (115.374;23) | 0.000 (100.081;22) | 0.000 (15.293;1) |
| **9V** | post booster | 25 | 6 | 52.2% | 1,706 | 8,457 | 0.078 | 83.0 (75.8-88.1) | 0.000 (135.599;23) | 0.000 (134.490;22) | 0.292 (1.108;1) |
| **14** | post booster | 25 | 6 | 46.8% | 1,705 | 8,446 | 0.063 | 71.9 (57.7-81.3) | 0.000 (81.916;23) | 0.000 (81.857;22) | 0.808 (0.059;1) |
| **18C** | post booster | 25 | 6 | 54.0% | 1,705 | 8,453 | 0.074 | 79.6 (70.3-86.0) | 0.000 (112.643;23) | 0.000 (112.118;22) | 0.469 (0.525;1) |
| **19F** | post booster | 25 | 6 | 53.8% | 1,707 | 8,443 | 0.113 | 87.9 (83.2-91.2) | 0.000 (189.802;23) | 0.000 (186.331;22) | 0.062 (3.471;1) |
| **23F** | post booster | 25 | 6 | 59.1% | 1,705 | 8,437 | 0.080 | 75.9 (64.4-83.7) | 0.000 (95.591;23) | 0.000 (95.039;22) | 0.457 (0.552;1) |
| **1** | post booster | 6 | 6 | 100% | 1,705 | | 0.045 | 48.7 (0-79.6) | 0.083 (9.739;5) | | |
| **5** | post booster | 6 | 6 | 100% | 1,706 | | 0.101 | 83.9 (66.4-92.3) | 0.000 (31.010;5) | | |
| **7F** | post booster | 6 | 6 | 100% | 1,705 | | 0.044 | 57.4 (0-82.8) | 0.039 (11.732;5) | | |

| | | | | | | | | | |
|---|---|---|---|---|---|---|---|---|---|
| **3** | post booster | 6 | 6 | 100% | 1,701 | | 0.299 | 96.9 (95.2-98.0) | 0.000 (162.413;5) |
| **6A** | post booster | 6 | 6 | 100% | 1,704 | | 0.239 | 94.1 (89.8-96.6) | 0.000 (85.221;5) |
| **19A** | post booster | 6 | 6 | 100% | 1,705 | | 0.158 | 87.1 (74.2-93.5) | 0.000 (38.745;5) |

df: degree of freedom; tau: square-root of between-study variance; $I^2$: heterogeneity statistic; Q of heterogeneity and inconsistency: overall heterogeneity/inconsistency statistic. Q of heterogeneity: overall heterogeneity statistic. Q of inconsistency: overall inconsistency statistic.

**Supplementary Table 4. Summary on assessment of statistical heterogeneity and incoherence for seroefficacy analysis shown in Figure 3**

| Serotype | No. of study | No. and proportion of study providing direct evidence | | No. of participants providing direct evidence | No. of participants providing indirect evidence | tau | $I^2$ (%) | p-value (Q of heterogeneity and inconsistency; df) | p-value (Q of heterogeneity; df) | p-value (Q of inconsistency; df) |
|---|---|---|---|---|---|---|---|---|---|---|
| **4** | 15 | 6 | 89.0% | 1,577 | 3,531 | 0.383 | 23.7 (0-59.5) | 0.198 (17.036;13) | 0.178 (16.286;12) | 0.386 (0.750;1) |
| **6B** | 15 | 6 | 91.6% | 1,573 | 3,466 | 0.249 | 74.4 (56.7-84.9) | 0.000 (50.756;13) | 0.000 (48.136;12) | 0.105 (2.621;1) |
| **9V** | 15 | 6 | 90.7% | 1,574 | 3,518 | 0.309 | 43.0 (0-69.6) | 0.044 (22.801;13) | 0.049 (21.065;12) | 0.188 (1.736;1) |
| **14** | 15 | 6 | 81.3% | 1,578 | 3,524 | 0.361 | 69.6 (47.4-82.5) | 0.000 (42.834;13) | 0.000 (40.642;12) | 0.139 (2.192;1) |
| **18C** | 15 | 6 | 95.1% | 1,568 | 3,537 | 0.474 | 49.9 (7.4-72.9) | 0.017 (25.961;13) | 0.017 (24.657;12) | 0.254 (1.304;1) |
| **19F** | 15 | 6 | 80.2% | 1,569 | 3,497 | 0.327 | 49.1 (5.7-72.5) | 0.020 (25.541;13) | 0.104 (18.415;12) | 0.008 (7.126;1) |
| **23F** | 15 | 6 | 93.6% | 1,570 | 3,498 | 0.557 | 83.8 (74.3-89.9) | 0.000 (80.480;13) | 0.000 (78.982;12) | 0.221 (1.498;1) |
| **1** | 6 | 6 | 100% | 1,581 | | 0.403 | 42.7 (0-77.3) | 0.121 (8.724;5) | | |
| **5** | 6 | 6 | 100% | 1,573 | | 0.975 | 82.2 (62.1-91.6) | 0.000 (28.031;5) | | |
| **7F** | 6 | 6 | 100% | 1,575 | | 0.000 | 0 (0-74.6) | 0.537 (4.084;5) | | |

| | | | | | | | |
|---|---|---|---|---|---|---|---|
| **3** | 6 | 6 | 100% | 1,571 | 0.580 | 67.4 (22.7-86.3) | 0.009 (15.356;5) |
| **6A** | 6 | 6 | 100% | 1,564 | 0.735 | 96.1 (93.6-97.6) | 0.000 (127.663;5) |
| **19A** | 6 | 6 | 100% | 1,571 | 0.836 | 91.4 (84.0-95.3) | 0.000 (57.901;5) |

df: degree of freedom; tau: square-root of between-study variance; $I^2$: heterogeneity statistic; Q of heterogeneity and inconsistency: overall heterogeneity/inconsistency statistic. Q of heterogeneity: overall heterogeneity statistic. Q of inconsistency: overall inconsistency statistic.

**Supplementary Figure 1. Network of studies included for A) all eligible cohorts, B) immunogenicity analysis cohorts and C) seroefficacy analysis cohorts**

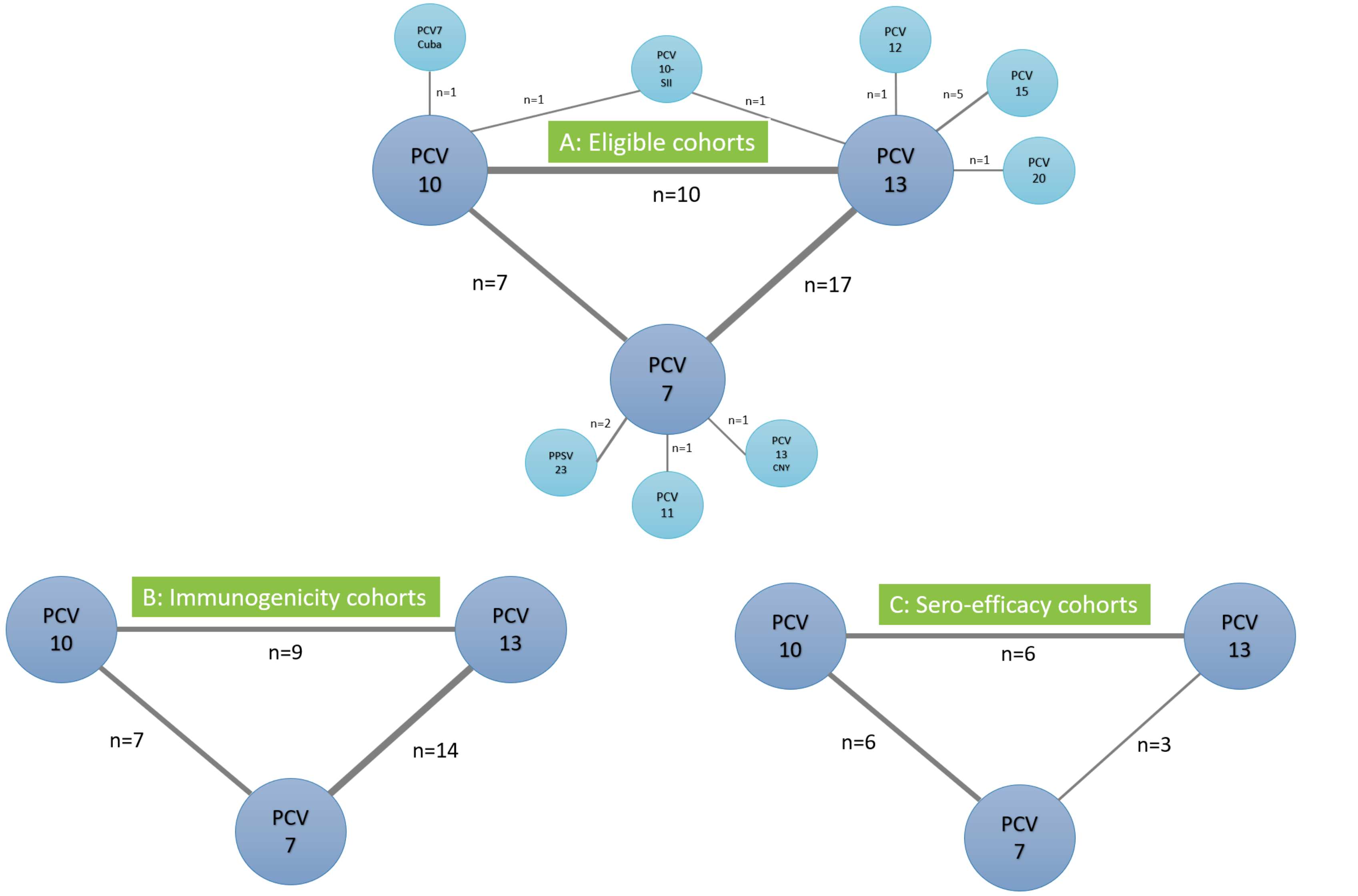

PCV – Pneumococcal conjugate vaccine; PPSV - Pneumococcal polysaccharide vaccine; PCV10-SII – 10-valent Pneumococcal conjugate vaccine by Serum Institute of India; PCV7 Cuba – a Cuban PCV7; PCV13 CNY – a Chinese PCV13

**Supplementary Figure 2. Assessment of risk of bias for included studies**

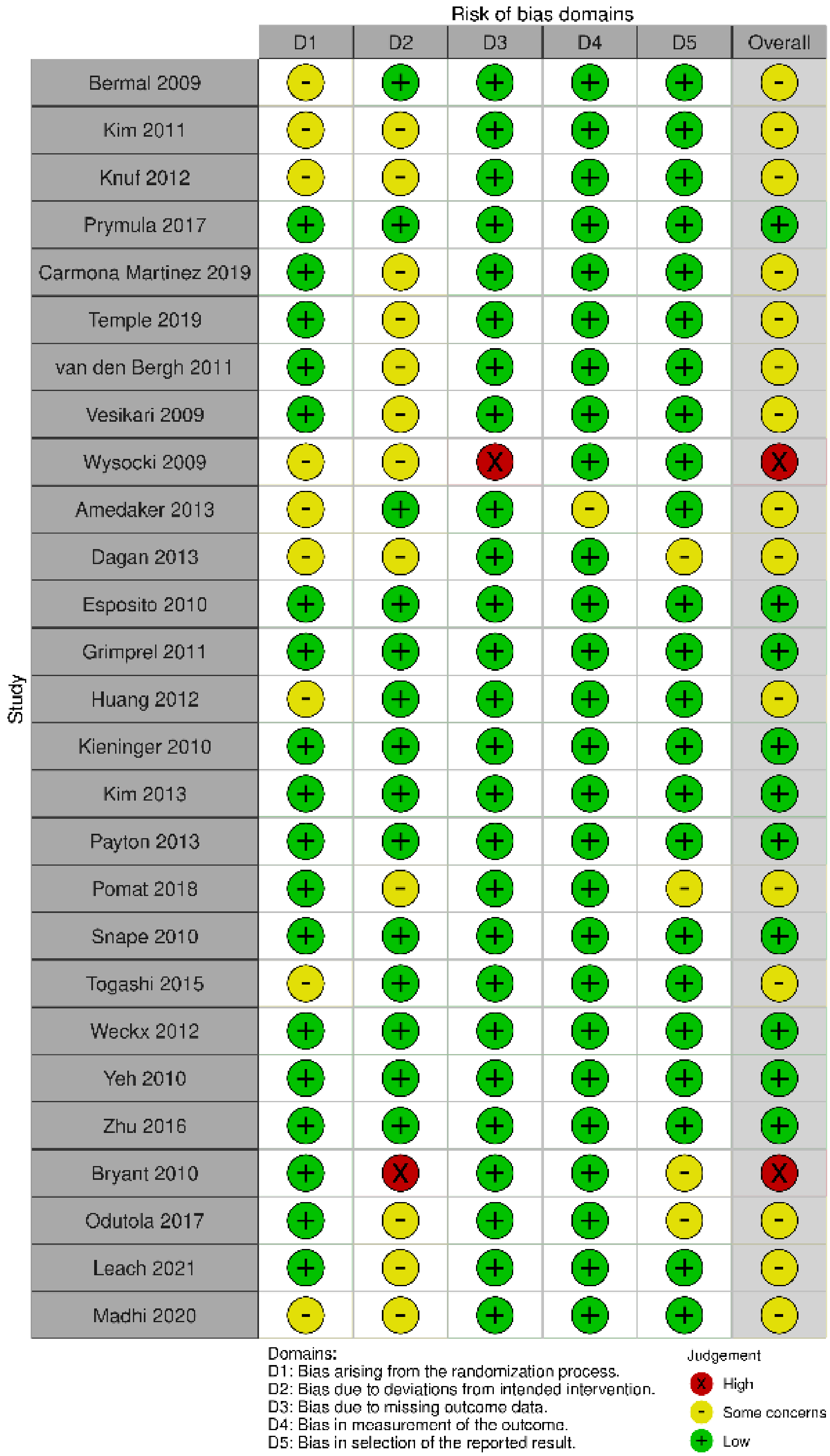

**Supplementary Figure 3. Direct and indirect evidence on geometric mean ratios comparing PCV13 vs PCV10 for serotypes in PCV7 at a) 28 days post-primary vaccination series, b) pre-booster, and c) 28 days post-booster**

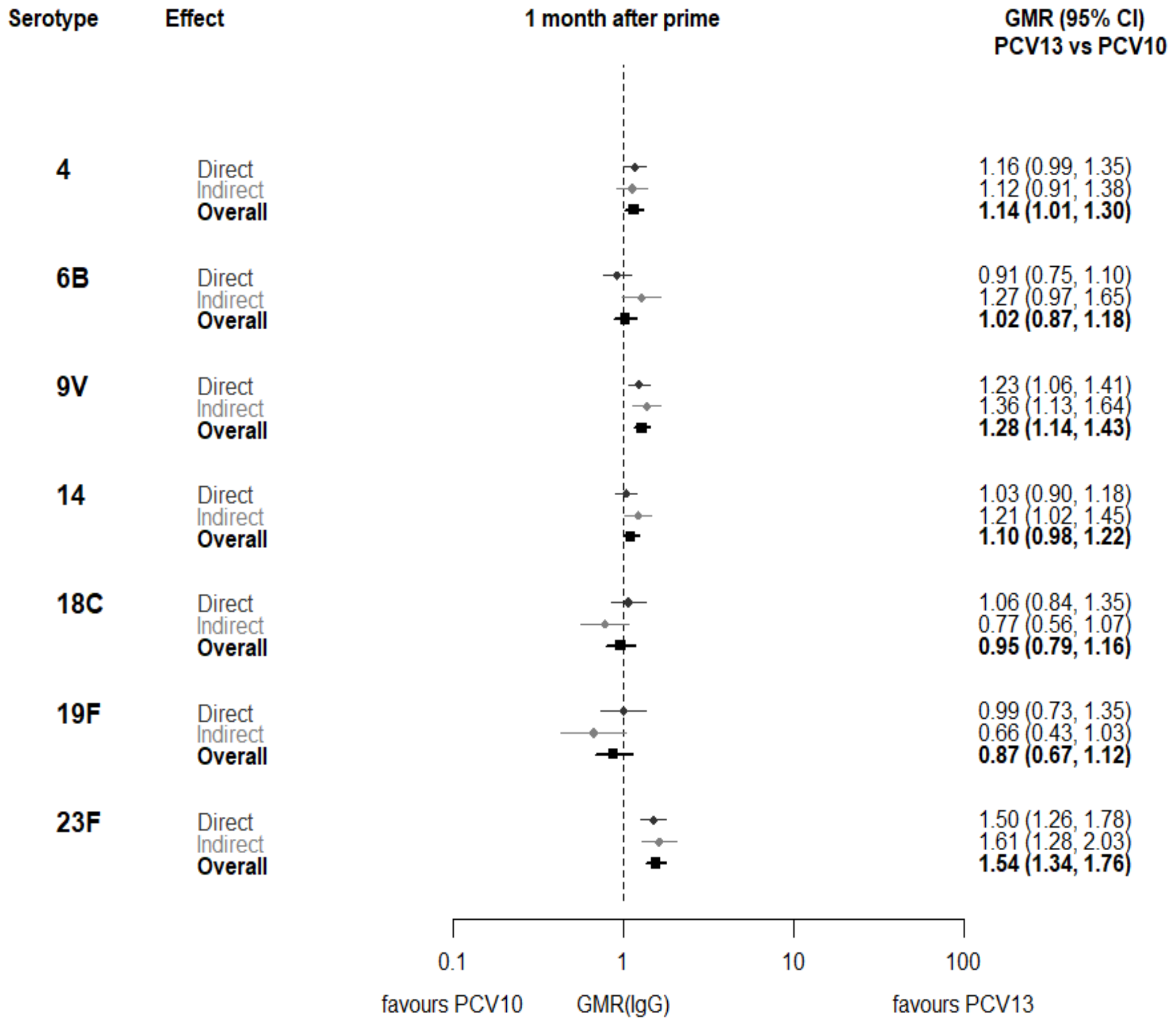

a)

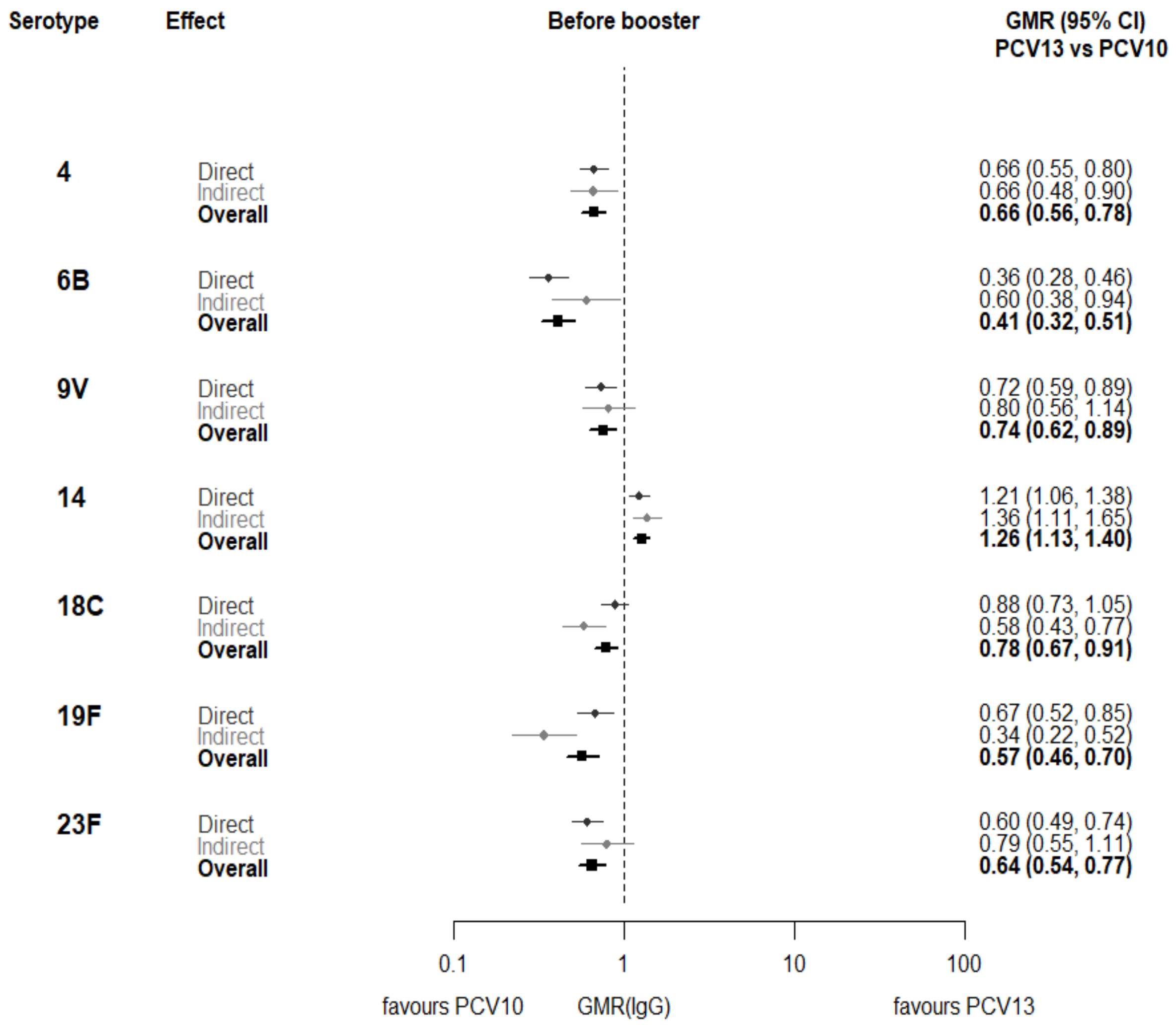
Serotype
Effect
Before booster
GMR (95% CI)
PCV13 vs PCV10
4
Direct
Indirect
Overall
0.66 (0.55, 0.80)
0.66 (0.48, 0.90)
0.66 (0.56, 0.78)
6B
Direct
Indirect
Overall
0.36 (0.28, 0.46)
0.60 (0.38, 0.94)
0.41 (0.32, 0.51)
9V
Direct
Indirect
Overall
0.72 (0.59, 0.89)
0.80 (0.56, 1.14)
0.74 (0.62, 0.89)
14
Direct
Indirect
Overall
1.21 (1.06, 1.38)
1.36 (1.11, 1.65)
1.26 (1.13, 1.40)
18C
Direct
Indirect
Overall
0.88 (0.73, 1.05)
0.58 (0.43, 0.77)
0.78 (0.67, 0.91)
19F
Direct
Indirect
Overall
0.67 (0.52, 0.85)
0.34 (0.22, 0.52)
0.57 (0.46, 0.70)
23F
Direct
Indirect
Overall
0.60 (0.49, 0.74)
0.79 (0.55, 1.11)
0.64 (0.54, 0.77)
0.1
1
10
100
favours PCV10
GMR(IgG)
favours PCV13

b)

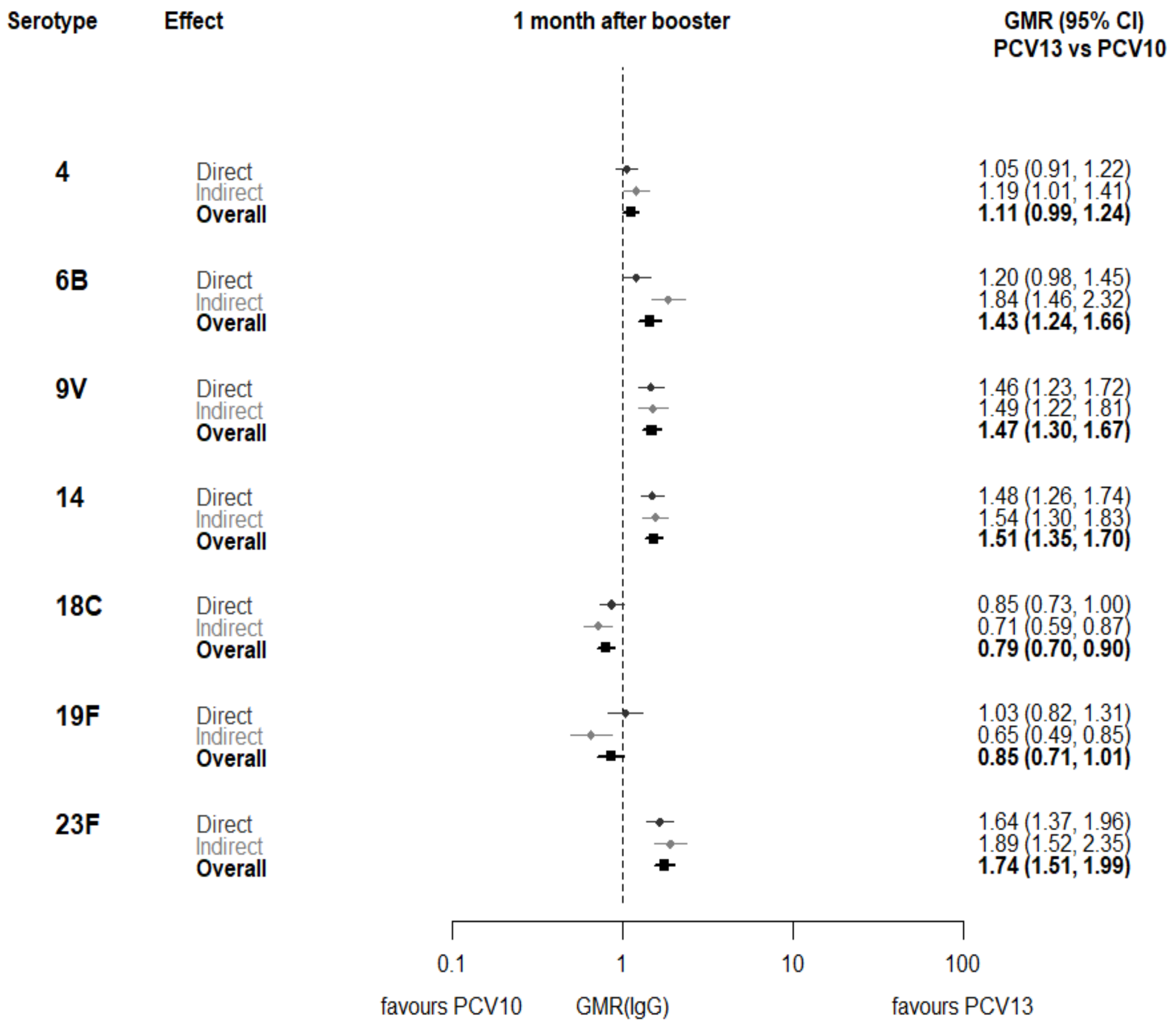

c)

GMR: Geometric mean ratio; PCV: Pneumococcal conjugate vaccine. Each line in the figure shows the output from a network meta-analyses. Dark grey diamonds and lines show the point estimates and confidence intervals for geometric mean ratios from studies directly comparing PCV13 vs PCV10. Light grey diamonds and lines show the point estimates and confidence intervals for geometric mean ratios from studies comparing PCV13 vs PCV10 through PCV7. Black boxes and lines show the point estimates and confidence intervals incorporating both direct and indirect evidence.

**Supplementary Figure 4. Trial level geometric mean ratios for serotype 4 at post-primary vaccination series.**

| ID | Comparison vac1 vs vac2 | Continent | Schedule-prime | Concomitant vaccines | Serotype 4 - post prime | GMR (95%CI) |
|---|---|---|---|---|---|---|
| 1 | *pcv7 vs pcv10* | Asia | 6-10-24 weeks | DTPw-HBV-Hib-TT + OPV | | 1.13 (0.95-1.33) |
| 1 | *pcv7 vs pcv10* | Europe | 2-4-6 months | DTPw-HBV-Hib-TT + IPV | | 1.29 (1.10-1.50) |
| 2 | *pcv7 vs pcv10* | Asia | 2-4-6 months | Hib-TT | | 1.31 (1.12-1.54) |
| 3 | *pcv7 vs pcv10* | Europe | 2-3-4 months | DTPa-HBV-Hib-TT/IPV | | 1.81 (1.39-2.36) |
| 7 | *pcv7 vs pcv10* | Europe | 2-3-4 months | DTPa-(HBV)-Hib-TT/IPV | | 1.52 (1.32-1.76) |
| 8 | *pcv7 vs pcv10* | Europe | 2-3-4 months | DTPa-(HBV)-Hib-TT/IPV | | 1.94 (1.78-2.12) |
| 9 | *pcv7 vs pcv10* | Europe | 2-4-6 months | DTPa-(HBV)-Hib-TT/IPV + MenC | | 1.50 (1.30-1.71) |
| 4 | *pcv13 vs pcv10* | Europe | 2-3-4 months | DTPa-HBV-Hib-TT/IPV | | 1.38 (1.10-1.73) |
| 5 | *pcv13 vs pcv10* | Europe | 2–3–4 months | DTPa-HBV-Hib-TT/IPV + (MenC-TT) | | 1.66 (1.42-1.94) |
| 6 | *pcv13 vs pcv10* | Asia | 2-4 months | DTPa-HBV-Hib-TT/IPV | | 1.50 (1.31-1.73) |
| 18 | *pcv13 vs pcv10* | Oceania | 1-2-3 months | DTPw-HBV-Hib-TT + OPV | | 1.14 (0.91-1.43) |
| 25 | *pcv13 vs pcv10* | Africa | 2–3–4 months | DTPw-HBV-Hib-TT + OPV | | 1.67 (1.41-1.97) |
| 26 | *pcv13 vs pcv10* | Oceania | 2-4-6 months | DTPa-HBV-Hib-TT/IPV/Rotavirus | | 1.27 (1.02-1.59) |
| 27 | *pcv13 vs pcv10* | Africa | 6 weeks | DTPa-HBV-Hib-TT/IPV/Rotavirus | | 0.60 (0.42-0.86) |
| 27 | *pcv13 vs pcv10* | Africa | 14 weeks | DTPa-HBV-Hib-TT/IPV/Rotavirus | | 0.53 (0.36-0.77) |
| 27 | *pcv13 vs pcv10* | Africa | 6-14 weeks | DTPa-HBV-Hib-TT/IPV/Rotavirus | | 0.76 (0.55-1.05) |
| 10 | *pcv13 vs pcv7* | Asia | 6-10-14 weeks | DTwP-Hib-HBV+OPV | | 0.85 (0.72-1.00) |
| 11 | *pcv13 vs pcv7* | Asia | 2-4-6 months | NA | | 1.07 (0.99-1.15) |
| 14 | *pcv13 vs pcv7* | Asia | 2-4-6 months | DTPa-(HBV)-Hib-TT/IPV | | 0.62 (0.50-0.78) |
| 15 | *pcv13 vs pcv7* | Europe | 2-3-4 months | DTPa-HBV-Hib-TT/IPV | | 0.73 (0.64-0.84) |
| 16 | *pcv13 vs pcv7* | Asia | 2-4-6 months | DTPa-HBV-Hib-TT/IPV | | 0.79 (0.64-0.98) |
| 17 | *pcv13 vs pcv7* | North America | 2-4-6 months | DTPa-HBV-Hib-TT/IPV | | 0.76 (0.68-0.86) |
| 19 | *pcv13 vs pcv7* | Europe | 2-4 months | DTaP-IPV-Hib/MenC | | 0.53 (0.36-0.78) |
| 20 | *pcv13 vs pcv7* | Asia | enr 3-6 m, 4-8 w int | DTPa | | 0.81 (0.71-0.93) |
| 21 | *pcv13 vs pcv7* | South America | 2-4-6 months | HBV-DTwP-Hib/OPV/Rotavirus | | 0.75 (0.65-0.87) |
| 22 | *pcv13 vs pcv7* | North America | 2-4-6 months | DTPa-HBV-Hib-TT/IPV | | 0.67 (0.58-0.76) |
| 23 | *pcv13 vs pcv7* | Asia | 3-4-5 months | NA | | 0.78 (0.70-0.87) |
| 24 | *pcv13 vs pcv7* | North America | 2-4-6 months | DTPa-HBV-Hib-TT/IPV | | 0.72 (0.57-0.91) |

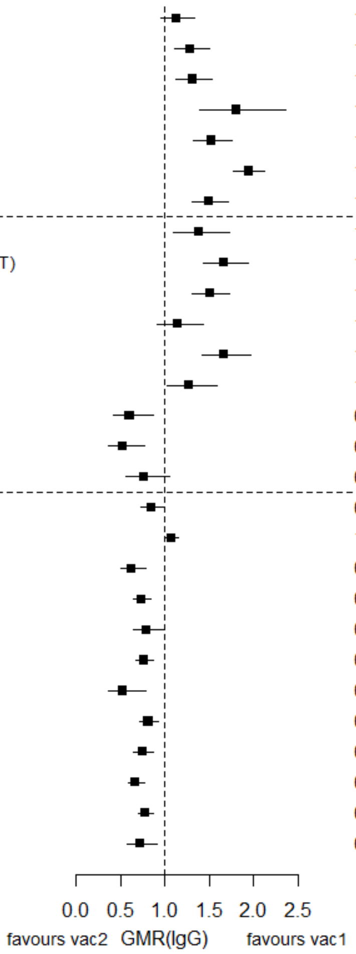

GMR: Geometric mean ratio; pcv: Pneumococcal conjugate vaccine; DTaP – diphtheria and tetanus toxoids, and acellular pertussis vaccine; DTwP – diphtheria and tetanus toxoids, and whole-cell pertussis vaccine; Hib-TT – Haemophilus influenzae type b vaccine (tetanus toxoid conjugate); HB – Hepatitis B vaccine; IPV – Inactivated polio vaccine; OPV – Oral polio vaccine; MenC – Meningococcal C vaccine; TT – tetanus toxoid conjugate; NA: not applicable; enr 3-6 m, 4-8 w int: enrolment at 3-6 months of age, and at 4-8 weeks interval of a 3 doses primary vaccines in total.

Each solid line in the figure shows the GMR from each trial. Black boxes and lines show the point estimates and confidence intervals for geometric mean ratios comparing vac1 vs vac2. Concomitant vaccines are vaccines co-administered with PCV primary vaccine series. Information on co-administered vaccine is not always available (e.g. study ID 11 and 23). Concomitant vaccines in the bracket are those administered in some but not all of the study sites.

**Supplementary Figure 5. Trial level geometric mean ratios for serotypes 6B at post-primary vaccination series.**

| ID | Comparison vac1 vs vac2 | Continent | Schedule-prime | Concomitant vaccines | Serotype 6B - post prime | GMR (95%CI) |
|---|---|---|---|---|---|---|
| 1 | *pcv7 vs pcv10* | Asia | 6-10-24 weeks | DTPw-HBV-Hib-TT + OPV | | 0.89 (0.66-1.20) |
| 1 | *pcv7 vs pcv10* | Europe | 2-4-6 months | DTPw-HBV-Hib-TT + IPV | | 1.60 (1.22-2.11) |
| 2 | *pcv7 vs pcv10* | Asia | 2-4-6 months | Hib-TT | | 1.45 (1.19-1.77) |
| 3 | *pcv7 vs pcv10* | Europe | 2-3-4 months | DTPa-HBV-Hib-TT/IPV | | 2.25 (1.39-3.64) |
| 7 | *pcv7 vs pcv10* | Europe | 2-3-4 months | DTPa-(HBV)-Hib-TT/IPV | | 1.20 (0.94-1.55) |
| 8 | *pcv7 vs pcv10* | Europe | 2-3-4 months | DTPa-(HBV)-Hib-TT/IPV | | 1.80 (1.55-2.09) |
| 9 | *pcv7 vs pcv10* | Europe | 2-4-6 months | DTPa-(HBV)-Hib-TT/IPV + MenC | | 1.55 (1.28-1.88) |
| 4 | *pcv13 vs pcv10* | Europe | 2-3-4 months | DTPa-HBV-Hib-TT/IPV | | 1.18 (0.87-1.61) |
| 5 | *pcv13 vs pcv10* | Europe | 2–3–4 months | DTPa-HBV-Hib-TT/IPV + (MenC-TT) | | 1.05 (0.82-1.33) |
| 6 | *pcv13 vs pcv10* | Asia | 2-4 months | DTPa-HBV-Hib-TT/IPV | | 0.61 (0.50-0.73) |
| 18 | *pcv13 vs pcv10* | Oceania | 1-2-3 months | DTPw-HBV-Hib-TT + OPV | | 1.20 (0.94-1.54) |
| 25 | *pcv13 vs pcv10* | Africa | 2–3–4 months | DTPw-HBV-Hib-TT + OPV | | 1.35 (1.00-1.82) |
| 26 | *pcv13 vs pcv10* | Oceania | 2-4-6 months | DTPa-HBV-Hib-TT/IPV/Rotavirus | | 1.15 (0.84-1.57) |
| 27 | *pcv13 vs pcv10* | Africa | 6 weeks | DTPa-HBV-Hib-TT/IPV/Rotavirus | | 0.71 (0.55-0.92) |
| 27 | *pcv13 vs pcv10* | Africa | 14 weeks | DTPa-HBV-Hib-TT/IPV/Rotavirus | | 0.89 (0.73-1.09) |
| 27 | *pcv13 vs pcv10* | Africa | 6-14 weeks | DTPa-HBV-Hib-TT/IPV/Rotavirus | | 0.43 (0.28-0.66) |
| 10 | *pcv13 vs pcv7* | Asia | 6-10-14 weeks | DTwP-Hib-HBV+OPV | | 0.92 (0.72-1.19) |
| 11 | *pcv13 vs pcv7* | Asia | 2-4-6 months | NA | | 0.98 (0.87-1.10) |
| 14 | *pcv13 vs pcv7* | Asia | 2-4-6 months | DTPa-(HBV)-Hib-TT/IPV | | 0.91 (0.70-1.17) |
| 15 | *pcv13 vs pcv7* | Europe | 2-3-4 months | DTPa-HBV-Hib-TT/IPV | | 0.64 (0.51-0.79) |
| 16 | *pcv13 vs pcv7* | Asia | 2-4-6 months | DTPa-HBV-Hib-TT/IPV | | 1.14 (0.86-1.52) |
| 17 | *pcv13 vs pcv7* | North America | 2-4-6 months | DTPa-HBV-Hib-TT/IPV | | 0.89 (0.74-1.06) |
| 19 | *pcv13 vs pcv7* | Europe | 2-4 months | DTaP-IPV-Hib/MenC | | 0.48 (0.29-0.80) |
| 20 | *pcv13 vs pcv7* | Asia | enr 3-6 m, 4-8 w int | DTPa | | 0.79 (0.65-0.97) |
| 21 | *pcv13 vs pcv7* | South America | 2-4-6 months | HBV-DTwP-Hib/OPV/Rotavirus | | 1.24 (0.97-1.59) |
| 22 | *pcv13 vs pcv7* | North America | 2-4-6 months | DTPa-HBV-Hib-TT/IPV | | 0.68 (0.53-0.86) |
| 23 | *pcv13 vs pcv7* | Asia | 3-4-5 months | NA | | 0.93 (0.79-1.08) |
| 24 | *pcv13 vs pcv7* | North America | 2-4-6 months | DTPa-HBV-Hib-TT/IPV | | 0.93 (0.61-1.41) |

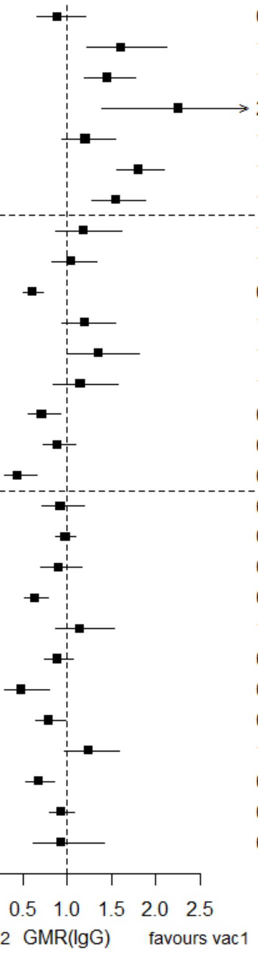

GMR: Geometric mean ratio; pcv: Pneumococcal conjugate vaccine; DTaP – diphtheria and tetanus toxoids, and acellular pertussis vaccine; DTwP – diphtheria and tetanus toxoids, and whole-cell pertussis vaccine; Hib-TT – Haemophilus influenzae type b vaccine (tetanus toxoid conjugate); HB – Hepatitis B vaccine; IPV – Inactivated polio vaccine; OPV – Oral polio vaccine; MenC – Meningococcal C vaccine; TT – tetanus toxoid conjugate; NA: not applicable; enr 3-6 m, 4-8 w int: enrolment at 3-6 months of age, and at 4-8 weeks interval of a 3 doses primary vaccines in total.

Each solid line in the figure shows the GMR from each trial. Black boxes and lines show the point estimates and confidence intervals for geometric mean ratios comparing vac1 vs vac2. Concomitant vaccines are vaccines co-administered with PCV primary vaccine series. Information on co-administered vaccine is not always available (e.g. study ID 11 and 23). Concomitant vaccines in the bracket are those administered in some but not all of the study sites.

**Supplementary Figure 6. Trial level geometric mean ratios for serotypes 9V at post-primary vaccination series.**

| ID | Comparison vac1 vs vac2 | Continent | Schedule-prime | Concomitant vaccines | GMR (95%CI) |
|---|---|---|---|---|---|
| 1 | *pcv7 vs pcv10* | Asia | 6-10-24 weeks | DTPw-HBV-Hib-TT + OPV | 1.24 (1.03-1.49) |
| 1 | *pcv7 vs pcv10* | Europe | 2-4-6 months | DTPw-HBV-Hib-TT + IPV | 1.71 (1.44-2.04) |
| 2 | *pcv7 vs pcv10* | Asia | 2-4-6 months | Hib-TT | 1.47 (1.23-1.76) |
| 3 | *pcv7 vs pcv10* | Europe | 2-3-4 months | DTPa-HBV-Hib-TT/IPV | 2.37 (1.78-3.16) |
| 7 | *pcv7 vs pcv10* | Europe | 2-3-4 months | DTPa-(HBV)-Hib-TT/IPV | 1.47 (1.27-1.70) |
| 8 | *pcv7 vs pcv10* | Europe | 2-3-4 months | DTPa-(HBV)-Hib-TT/IPV | 2.09 (1.91-2.29) |
| 9 | *pcv7 vs pcv10* | Europe | 2-4-6 months | DTPa-(HBV)-Hib-TT/IPV + MenC | 1.85 (1.59-2.16) |
| 4 | *pcv13 vs pcv10* | Europe | 2-3-4 months | DTPa-HBV-Hib-TT/IPV | 1.19 (0.96-1.48) |
| 5 | *pcv13 vs pcv10* | Europe | 2–3–4 months | DTPa-HBV-Hib-TT/IPV + (MenC-TT) | 1.57 (1.36-1.82) |
| 6 | *pcv13 vs pcv10* | Asia | 2-4 months | DTPa-HBV-Hib-TT/IPV | 2.00 (1.72-2.33) |
| 18 | *pcv13 vs pcv10* | Oceania | 1-2-3 months | DTPw-HBV-Hib-TT + OPV | 1.19 (0.95-1.49) |
| 25 | *pcv13 vs pcv10* | Africa | 2–3–4 months | DTPw-HBV-Hib-TT + OPV | 1.23 (1.00-1.52) |
| 26 | *pcv13 vs pcv10* | Oceania | 2-4-6 months | DTPa-HBV-Hib-TT/IPV/Rotavirus | 1.55 (1.26-1.92) |
| 27 | *pcv13 vs pcv10* | Africa | 6 weeks | DTPa-HBV-Hib-TT/IPV/Rotavirus | 0.56 (0.41-0.75) |
| 27 | *pcv13 vs pcv10* | Africa | 14 weeks | DTPa-HBV-Hib-TT/IPV/Rotavirus | 0.88 (0.60-1.29) |
| 27 | *pcv13 vs pcv10* | Africa | 6-14 weeks | DTPa-HBV-Hib-TT/IPV/Rotavirus | 0.88 (0.61-1.28) |
| 10 | *pcv13 vs pcv7* | Asia | 6-10-14 weeks | DTwP-Hib-HBV+OPV | 0.92 (0.79-1.08) |
| 11 | *pcv13 vs pcv7* | Asia | 2-4-6 months | NA | 0.98 (0.91-1.05) |
| 14 | *pcv13 vs pcv7* | Asia | 2-4-6 months | DTPa-(HBV)-Hib-TT/IPV | 0.67 (0.56-0.82) |
| 15 | *pcv13 vs pcv7* | Europe | 2-3-4 months | DTPa-HBV-Hib-TT/IPV | 0.83 (0.73-0.94) |
| 16 | *pcv13 vs pcv7* | Asia | 2-4-6 months | DTPa-HBV-Hib-TT/IPV | 0.89 (0.74-1.08) |
| 17 | *pcv13 vs pcv7* | North America | 2-4-6 months | DTPa-HBV-Hib-TT/IPV | 0.81 (0.74-0.90) |
| 19 | *pcv13 vs pcv7* | Europe | 2-4 months | DTaP-IPV-Hib/MenC | 0.62 (0.41-0.93) |
| 20 | *pcv13 vs pcv7* | Asia | enr 3-6 m, 4-8 w int | DTPa | 0.74 (0.64-0.86) |
| 21 | *pcv13 vs pcv7* | South America | 2-4-6 months | HBV-DTwP-Hib/OPV/Rotavirus | 0.87 (0.75-1.01) |
| 22 | *pcv13 vs pcv7* | North America | 2-4-6 months | DTPa-HBV-Hib-TT/IPV | 0.70 (0.61-0.80) |
| 23 | *pcv13 vs pcv7* | Asia | 3-4-5 months | NA | 0.77 (0.70-0.85) |
| 24 | *pcv13 vs pcv7* | North America | 2-4-6 months | DTPa-HBV-Hib-TT/IPV | 0.81 (0.66-0.99) |

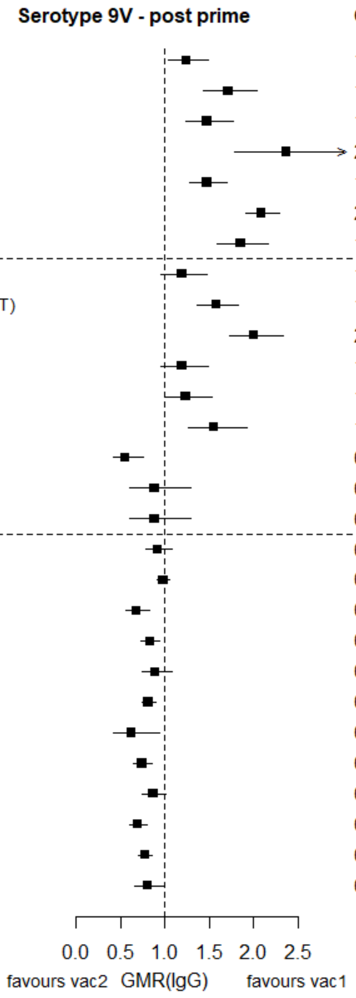

GMR: Geometric mean ratio; pcv: Pneumococcal conjugate vaccine; DTaP – diphtheria and tetanus toxoids, and acellular pertussis vaccine; DTwP – diphtheria and tetanus toxoids, and whole-cell pertussis vaccine; Hib-TT – Haemophilus influenzae type b vaccine (tetanus toxoid conjugate); HB – Hepatitis B vaccine; IPV – Inactivated polio vaccine; OPV – Oral polio vaccine; MenC – Meningococcal C vaccine; TT – tetanus toxoid conjugate; NA: not applicable; enr 3-6 m, 4-8 w int: enrolment at 3-6 months of age, and at 4-8 weeks interval of a 3 doses primary vaccines in total.

Each solid line in the figure shows the GMR from each trial. Black boxes and lines show the point estimates and confidence intervals for geometric mean ratios comparing vac1 vs vac2. Concomitant vaccines are vaccines co-administered with PCV primary vaccine series. Information on co-administered vaccine is not always available (e.g. study ID 11 and 23). Concomitant vaccines in the bracket are those administered in some but not all of the study sites.

**Supplementary Figure 7. Trial level geometric mean ratios for serotypes 14 at post-primary vaccination series.**

| ID | Comparison vac1 vs vac2 | Continent | Schedule-prime | Concomitant vaccines | GMR (95%CI) |
|---|---|---|---|---|---|
| 1 | *pcv7 vs pcv10* | Asia | 6-10-24 weeks | DTPw-HBV-Hib-TT + OPV | 0.90 (0.70-1.16) |
| 1 | *pcv7 vs pcv10* | Europe | 2-4-6 months | DTPw-HBV-Hib-TT + IPV | 1.55 (1.27-1.89) |
| 2 | *pcv7 vs pcv10* | Asia | 2-4-6 months | Hib-TT | 1.53 (1.27-1.83) |
| 3 | *pcv7 vs pcv10* | Europe | 2-3-4 months | DTPa-HBV-Hib-TT/IPV | 1.80 (1.29-2.51) |
| 7 | *pcv7 vs pcv10* | Europe | 2-3-4 months | DTPa-(HBV)-Hib-TT/IPV | 1.09 (0.95-1.26) |
| 8 | *pcv7 vs pcv10* | Europe | 2-3-4 months | DTPa-(HBV)-Hib-TT/IPV | 1.59 (1.43-1.77) |
| 9 | *pcv7 vs pcv10* | Europe | 2-4-6 months | DTPa-(HBV)-Hib-TT/IPV + MenC | 1.61 (1.34-1.94) |
| 4 | *pcv13 vs pcv10* | Europe | 2-3-4 months | DTPa-HBV-Hib-TT/IPV | 1.11 (0.88-1.41) |
| 5 | *pcv13 vs pcv10* | Europe | 2–3–4 months | DTPa-HBV-Hib-TT/IPV + (MenC-TT) | 1.01 (0.85-1.20) |
| 6 | *pcv13 vs pcv10* | Asia | 2-4 months | DTPa-HBV-Hib-TT/IPV | 1.36 (1.11-1.68) |
| 18 | *pcv13 vs pcv10* | Oceania | 1-2-3 months | DTPw-HBV-Hib-TT + OPV | 0.73 (0.55-0.98) |
| 25 | *pcv13 vs pcv10* | Africa | 2–3–4 months | DTPw-HBV-Hib-TT + OPV | 1.12 (0.89-1.42) |
| 26 | *pcv13 vs pcv10* | Oceania | 2-4-6 months | DTPa-HBV-Hib-TT/IPV/Rotavirus | 1.37 (1.02-1.84) |
| 27 | *pcv13 vs pcv10* | Africa | 6 weeks | DTPa-HBV-Hib-TT/IPV/Rotavirus | 0.73 (0.52-1.03) |
| 27 | *pcv13 vs pcv10* | Africa | 14 weeks | DTPa-HBV-Hib-TT/IPV/Rotavirus | 1.44 (1.04-1.99) |
| 27 | *pcv13 vs pcv10* | Africa | 6-14 weeks | DTPa-HBV-Hib-TT/IPV/Rotavirus | 0.48 (0.31-0.74) |
| 10 | *pcv13 vs pcv7* | Asia | 6-10-14 weeks | DTwP-Hib-HBV+OPV | 1.20 (0.93-1.55) |
| 11 | *pcv13 vs pcv7* | Asia | 2-4-6 months | NA | 0.89 (0.79-0.99) |
| 14 | *pcv13 vs pcv7* | Asia | 2-4-6 months | DTPa-(HBV)-Hib-TT/IPV | 0.84 (0.67-1.06) |
| 15 | *pcv13 vs pcv7* | Europe | 2-3-4 months | DTPa-HBV-Hib-TT/IPV | 0.87 (0.74-1.04) |
| 16 | *pcv13 vs pcv7* | Asia | 2-4-6 months | DTPa-HBV-Hib-TT/IPV | 0.88 (0.67-1.16) |
| 17 | *pcv13 vs pcv7* | North America | 2-4-6 months | DTPa-HBV-Hib-TT/IPV | 0.83 (0.72-0.96) |
| 19 | *pcv13 vs pcv7* | Europe | 2-4 months | DTaP-IPV-Hib/MenC | 0.51 (0.32-0.84) |
| 20 | *pcv13 vs pcv7* | Asia | enr 3-6 m, 4-8 w int | DTPa | 0.82 (0.70-0.97) |
| 21 | *pcv13 vs pcv7* | South America | 2-4-6 months | HBV-DTwP-Hib/OPV/Rotavirus | 0.92 (0.73-1.16) |
| 22 | *pcv13 vs pcv7* | North America | 2-4-6 months | DTPa-HBV-Hib-TT/IPV | 0.84 (0.71-1.00) |
| 23 | *pcv13 vs pcv7* | Asia | 3-4-5 months | NA | 0.91 (0.79-1.04) |
| 24 | *pcv13 vs pcv7* | North America | 2-4-6 months | DTPa-HBV-Hib-TT/IPV | 0.80 (0.60-1.08) |

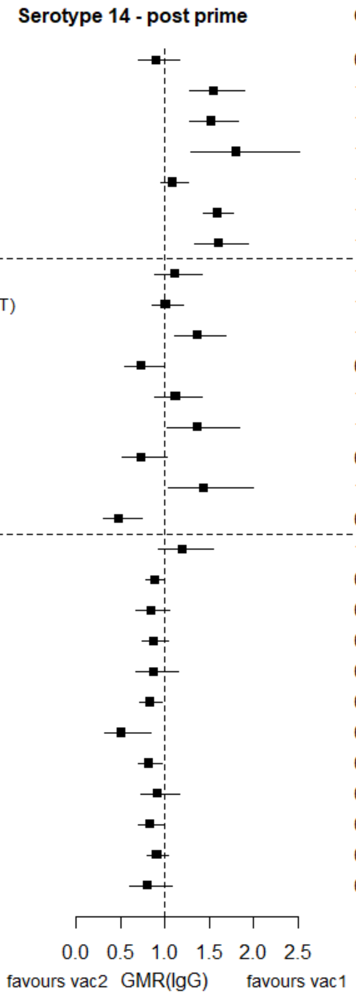

GMR: Geometric mean ratio; pcv: Pneumococcal conjugate vaccine; DTaP – diphtheria and tetanus toxoids, and acellular pertussis vaccine; DTwP – diphtheria and tetanus toxoids, and whole-cell pertussis vaccine; Hib-TT – Haemophilus influenzae type b vaccine (tetanus toxoid conjugate); HB – Hepatitis B vaccine; IPV – Inactivated polio vaccine; OPV – Oral polio vaccine; MenC – Meningococcal C vaccine; TT – tetanus toxoid conjugate; NA: not applicable; enr 3-6 m, 4-8 w int: enrolment at 3-6 months of age, and at 4-8 weeks interval of a 3 doses primary vaccines in total.

Each solid line in the figure shows the GMR from each trial. Black boxes and lines show the point estimates and confidence intervals for geometric mean ratios comparing vac1 vs vac2. Concomitant vaccines are vaccines co-administered with PCV primary vaccine series. Information on co-administered vaccine is not always available (e.g. study ID 11 and 23). Concomitant vaccines in the bracket are those administered in some but not all of the study sites.

**Supplementary Figure 8. Trial level geometric mean ratios for serotypes 18C at post-primary vaccination series.**

| ID | Comparison vac1 vs vac2 | Continent | Schedule-prime | Concomitant vaccines | GMR (95%CI) |
|---|---|---|---|---|---|
| 1 | *pcv7 vs pcv10* | Asia | 6-10-24 weeks | DTPw-HBV-Hib-TT + OPV | 0.32 (0.26-0.39) |
| 1 | *pcv7 vs pcv10* | Europe | 2-4-6 months | DTPw-HBV-Hib-TT + IPV | 0.69 (0.57-0.85) |
| 2 | *pcv7 vs pcv10* | Asia | 2-4-6 months | Hib-TT | 0.86 (0.70-1.06) |
| 3 | *pcv7 vs pcv10* | Europe | 2-3-4 months | DTPa-HBV-Hib-TT/IPV | 0.93 (0.67-1.28) |
| 7 | *pcv7 vs pcv10* | Europe | 2-3-4 months | DTPa-(HBV)-Hib-TT/IPV | 1.53 (1.29-1.82) |
| 8 | *pcv7 vs pcv10* | Europe | 2-3-4 months | DTPa-(HBV)-Hib-TT/IPV | 1.48 (1.33-1.65) |
| 9 | *pcv7 vs pcv10* | Europe | 2-4-6 months | DTPa-(HBV)-Hib-TT/IPV + MenC | 1.06 (0.91-1.23) |
| 4 | *pcv13 vs pcv10* | Europe | 2-3-4 months | DTPa-HBV-Hib-TT/IPV | 1.05 (0.82-1.35) |
| 5 | *pcv13 vs pcv10* | Europe | 2–3–4 months | DTPa-HBV-Hib-TT/IPV + (MenC-TT) | 1.25 (1.05-1.50) |
| 6 | *pcv13 vs pcv10* | Asia | 2-4 months | DTPa-HBV-Hib-TT/IPV | 1.69 (1.44-1.98) |
| 18 | *pcv13 vs pcv10* | Oceania | 1-2-3 months | DTPw-HBV-Hib-TT + OPV | 0.88 (0.68-1.16) |
| 25 | *pcv13 vs pcv10* | Africa | 2–3–4 months | DTPw-HBV-Hib-TT + OPV | 0.37 (0.30-0.45) |
| 26 | *pcv13 vs pcv10* | Oceania | 2-4-6 months | DTPa-HBV-Hib-TT/IPV/Rotavirus | 0.93 (0.72-1.20) |
| 27 | *pcv13 vs pcv10* | Africa | 6 weeks | DTPa-HBV-Hib-TT/IPV/Rotavirus | 1.50 (1.15-1.97) |
| 27 | *pcv13 vs pcv10* | Africa | 14 weeks | DTPa-HBV-Hib-TT/IPV/Rotavirus | 1.19 (0.92-1.56) |
| 27 | *pcv13 vs pcv10* | Africa | 6-14 weeks | DTPa-HBV-Hib-TT/IPV/Rotavirus | 1.49 (1.07-2.08) |
| 10 | *pcv13 vs pcv7* | Asia | 6-10-14 weeks | DTwP-Hib-HBV+OPV | 1.02 (0.85-1.22) |
| 11 | *pcv13 vs pcv7* | Asia | 2-4-6 months | NA | 1.02 (0.93-1.11) |
| 14 | *pcv13 vs pcv7* | Asia | 2-4-6 months | DTPa-(HBV)-Hib-TT/IPV | 0.78 (0.63-0.97) |
| 15 | *pcv13 vs pcv7* | Europe | 2-3-4 months | DTPa-HBV-Hib-TT/IPV | 0.87 (0.76-1.00) |
| 16 | *pcv13 vs pcv7* | Asia | 2-4-6 months | DTPa-HBV-Hib-TT/IPV | 0.98 (0.81-1.19) |
| 17 | *pcv13 vs pcv7* | North America | 2-4-6 months | DTPa-HBV-Hib-TT/IPV | 0.86 (0.77-0.97) |
| 19 | *pcv13 vs pcv7* | Europe | 2-4 months | DTaP-IPV-Hib/MenC | 1.10 (0.76-1.60) |
| 20 | *pcv13 vs pcv7* | Asia | enr 3-6 m, 4-8 w int | DTPa | 0.72 (0.62-0.84) |
| 21 | *pcv13 vs pcv7* | South America | 2-4-6 months | HBV-DTwP-Hib/OPV/Rotavirus | 0.90 (0.76-1.07) |
| 22 | *pcv13 vs pcv7* | North America | 2-4-6 months | DTPa-HBV-Hib-TT/IPV | 0.75 (0.66-0.86) |
| 23 | *pcv13 vs pcv7* | Asia | 3-4-5 months | NA | 0.95 (0.84-1.07) |
| 24 | *pcv13 vs pcv7* | North America | 2-4-6 months | DTPa-HBV-Hib-TT/IPV | 0.60 (0.49-0.77) |

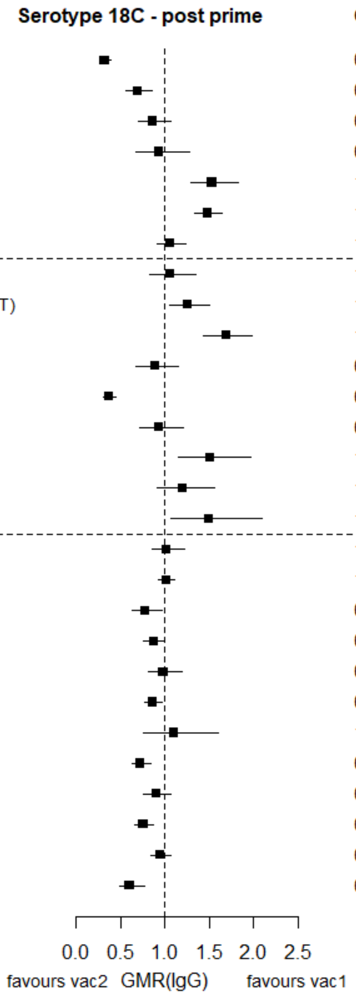

GMR: Geometric mean ratio; pcv: Pneumococcal conjugate vaccine; DTaP – diphtheria and tetanus toxoids, and acellular pertussis vaccine; DTwP – diphtheria and tetanus toxoids, and whole-cell pertussis vaccine; Hib-TT – Haemophilus influenzae type b vaccine (tetanus toxoid conjugate); HB – Hepatitis B vaccine; IPV – Inactivated polio vaccine; OPV – Oral polio vaccine; MenC – Meningococcal C vaccine; TT – tetanus toxoid conjugate; NA: not applicable; enr 3-6 m, 4-8 w int: enrolment at 3-6 months of age, and at 4-8 weeks interval of a 3 doses primary vaccines in total.

Each solid line in the figure shows the GMR from each trial. Black boxes and lines show the point estimates and confidence intervals for geometric mean ratios comparing vac1 vs vac2. Concomitant vaccines are vaccines co-administered with PCV primary vaccine series. Information on co-administered vaccine is not always available (e.g. study ID 11 and 23). Concomitant vaccines in the bracket are those administered in some but not all of the study sites.

**Supplementary Figure 9. Trial level geometric mean ratios for serotypes 19F at post-primary vaccination series.**

| ID | Comparison vac1 vs vac2 | Continent | Schedule-prime | Concomitant vaccines | Serotype 19F - post prime | GMR (95%CI) |
|---|---|---|---|---|---|---|
| 1 | *pcv7 vs pcv10* | Asia | 6-10-24 weeks | DTPw-HBV-Hib-TT + OPV | | 0.45 (0.37-0.54) |
| 1 | *pcv7 vs pcv10* | Europe | 2-4-6 months | DTPw-HBV-Hib-TT + IPV | | 0.44 (0.36-0.53) |
| 2 | *pcv7 vs pcv10* | Asia | 2-4-6 months | Hib-TT | | 0.37 (0.31-0.43) |
| 3 | *pcv7 vs pcv10* | Europe | 2-3-4 months | DTPa-HBV-Hib-TT/IPV | | 1.22 (0.88-1.69) |
| 7 | *pcv7 vs pcv10* | Europe | 2-3-4 months | DTPa-(HBV)-Hib-TT/IPV | | 1.50 (1.27-1.77) |
| 8 | *pcv7 vs pcv10* | Europe | 2-3-4 months | DTPa-(HBV)-Hib-TT/IPV | | 1.83 (1.65-2.03) |
| 9 | *pcv7 vs pcv10* | Europe | 2-4-6 months | DTPa-(HBV)-Hib-TT/IPV + MenC | | 0.57 (0.50-0.65) |
| 4 | *pcv13 vs pcv10* | Europe | 2-3-4 months | DTPa-HBV-Hib-TT/IPV | | 0.79 (0.62-1.00) |
| 5 | *pcv13 vs pcv10* | Europe | 2–3–4 months | DTPa-HBV-Hib-TT/IPV + (MenC-TT) | | 0.81 (0.69-0.96) |
| 6 | *pcv13 vs pcv10* | Asia | 2-4 months | DTPa-HBV-Hib-TT/IPV | | 0.80 (0.67-0.96) |
| 18 | *pcv13 vs pcv10* | Oceania | 1-2-3 months | DTPw-HBV-Hib-TT + OPV | | 1.25 (1.00-1.55) |
| 25 | *pcv13 vs pcv10* | Africa | 2–3–4 months | DTPw-HBV-Hib-TT + OPV | | 0.60 (0.49-0.72) |
| 26 | *pcv13 vs pcv10* | Oceania | 2-4-6 months | DTPa-HBV-Hib-TT/IPV/Rotavirus | | 0.82 (0.62-1.09) |
| 27 | *pcv13 vs pcv10* | Africa | 6 weeks | DTPa-HBV-Hib-TT/IPV/Rotavirus | | 1.01 (0.81-1.27) |
| 27 | *pcv13 vs pcv10* | Africa | 14 weeks | DTPa-HBV-Hib-TT/IPV/Rotavirus | | 1.64 (1.26-2.14) |
| 27 | *pcv13 vs pcv10* | Africa | 6-14 weeks | DTPa-HBV-Hib-TT/IPV/Rotavirus | | 1.90 (1.39-2.61) |
| 10 | *pcv13 vs pcv7* | Asia | 6-10-14 weeks | DTwP-Hib-HBV+OPV | | 0.91 (0.75-1.10) |
| 11 | *pcv13 vs pcv7* | Asia | 2-4-6 months | NA | | 1.30 (1.19-1.42) |
| 14 | *pcv13 vs pcv7* | Asia | 2-4-6 months | DTPa-(HBV)-Hib-TT/IPV | | 0.75 (0.60-0.94) |
| 15 | *pcv13 vs pcv7* | Europe | 2-3-4 months | DTPa-HBV-Hib-TT/IPV | | 0.59 (0.51-0.69) |
| 16 | *pcv13 vs pcv7* | Asia | 2-4-6 months | DTPa-HBV-Hib-TT/IPV | | 0.96 (0.74-1.24) |
| 17 | *pcv13 vs pcv7* | North America | 2-4-6 months | DTPa-HBV-Hib-TT/IPV | | 0.96 (0.84-1.10) |
| 19 | *pcv13 vs pcv7* | Europe | 2-4 months | DTaP-IPV-Hib/MenC | | 0.80 (0.50-1.30) |
| 20 | *pcv13 vs pcv7* | Asia | enr 3-6 m, 4-8 w int | DTPa | | 0.87 (0.71-1.07) |
| 21 | *pcv13 vs pcv7* | South America | 2-4-6 months | HBV-DTwP-Hib/OPV/Rotavirus | | 0.74 (0.61-0.90) |
| 22 | *pcv13 vs pcv7* | North America | 2-4-6 months | DTPa-HBV-Hib-TT/IPV | | 0.82 (0.71-0.94) |
| 23 | *pcv13 vs pcv7* | Asia | 3-4-5 months | NA | | 1.29 (1.07-1.55) |
| 24 | *pcv13 vs pcv7* | North America | 2-4-6 months | DTPa-HBV-Hib-TT/IPV | | 0.82 (0.65-1.04) |

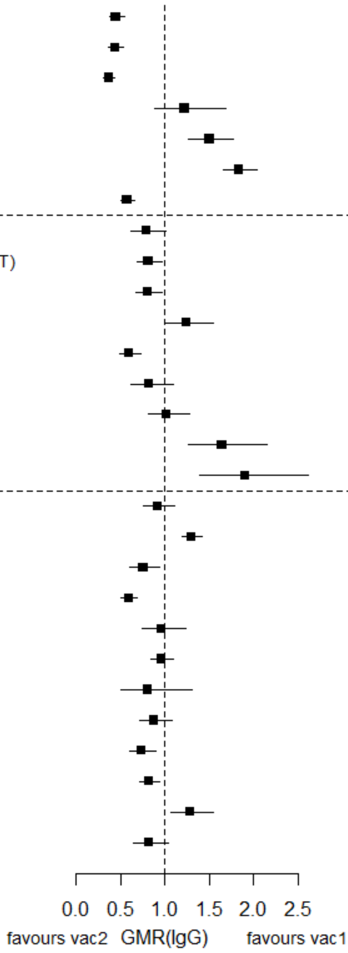

GMR: Geometric mean ratio; pcv: Pneumococcal conjugate vaccine; DTaP – diphtheria and tetanus toxoids, and acellular pertussis vaccine; DTwP – diphtheria and tetanus toxoids, and whole-cell pertussis vaccine; Hib-TT – Haemophilus influenzae type b vaccine (tetanus toxoid conjugate); HB – Hepatitis B vaccine; IPV – Inactivated polio vaccine; OPV – Oral polio vaccine; MenC – Meningococcal C vaccine; TT – tetanus toxoid conjugate; NA: not applicable; enr 3-6 m, 4-8 w int: enrolment at 3-6 months of age, and at 4-8 weeks interval of a 3 doses primary vaccines in total.

Each solid line in the figure shows the GMR from each trial. Black boxes and lines show the point estimates and confidence intervals for geometric mean ratios comparing vac1 vs vac2. Concomitant vaccines are vaccines co-administered with PCV primary vaccine series. Information on co-administered vaccine is not always available (e.g. study ID 11 and 23). Concomitant vaccines in the bracket are those administered in some but not all of the study sites.

**Supplementary Figure 10. Trial level geometric mean ratios for serotypes 23F at post-primary vaccination series.**

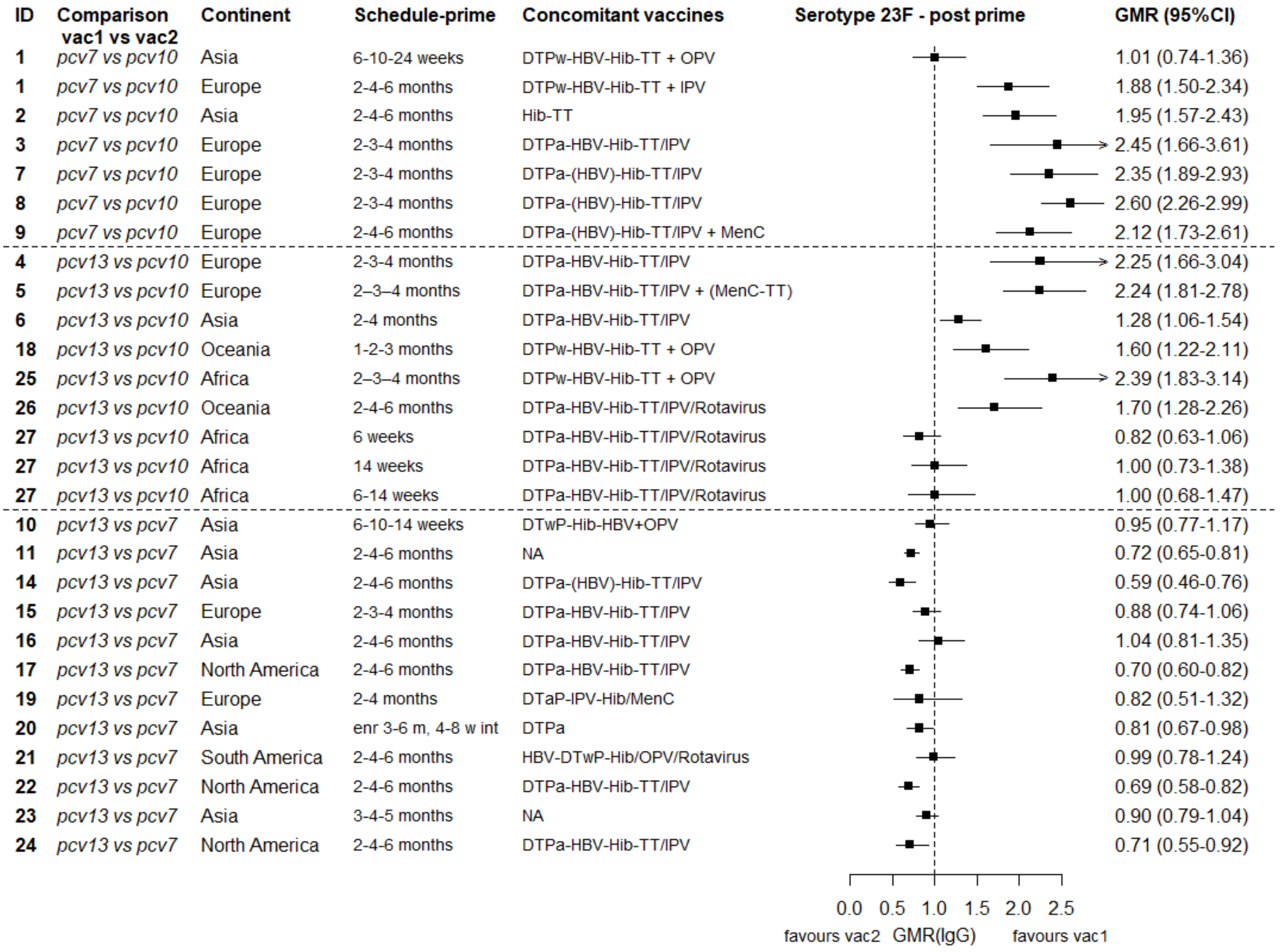

GMR: Geometric mean ratio; pcv: Pneumococcal conjugate vaccine; DTaP – diphtheria and tetanus toxoids, and acellular pertussis vaccine; DTwP – diphtheria and tetanus toxoids, and whole-cell pertussis vaccine; Hib-TT – Haemophilus influenzae type b vaccine (tetanus toxoid conjugate); HB – Hepatitis B vaccine; IPV – Inactivated polio vaccine; OPV – Oral polio vaccine; MenC – Meningococcal C vaccine; TT – tetanus toxoid conjugate; NA: not applicable; enr 3-6 m, 4-8 w int: enrolment at 3-6 months of age, and at 4-8 weeks interval of a 3 doses primary vaccines in total.

Each solid line in the figure shows the GMR from each trial. Black boxes and lines show the point estimates and confidence intervals for geometric mean ratios comparing vac1 vs vac2. Concomitant vaccines are vaccines co-administered with PCV primary vaccine series. Information on co-administered vaccine is not always available (e.g. study ID 11 and 23). Concomitant vaccines in the bracket are those administered in some but not all of the study sites.

**Supplementary Figure 11. Trial level geometric mean ratios for serotypes 1 at post-primary vaccination series.**

| ID | Comparison vac1 vs vac2 | Continent | Schedule-prime | Concomitant vaccines | GMR (95%CI) |
|---|---|---|---|---|---|
| **4** | *pcv13 vs pcv10* | Europe | 2-3-4 months | DTPa-HBV-Hib-TT/IPV | 1.49 (1.20-1.86) |
| **5** | *pcv13 vs pcv10* | Europe | 2–3–4 months | DTPa-HBV-Hib-TT/IPV + (MenC-TT) | 1.62 (1.39-1.88) |
| **6** | *pcv13 vs pcv10* | Asia | 2-4 months | DTPa-HBV-Hib-TT/IPV | 2.21 (1.89-2.57) |
| **18** | *pcv13 vs pcv10* | Oceania | 1-2-3 months | DTPw-HBV-Hib-TT + OPV | 1.15 (0.93-1.42) |
| **25** | *pcv13 vs pcv10* | Africa | 2–3–4 months | DTPw-HBV-Hib-TT + OPV | 1.63 (1.37-1.93) |
| **26** | *pcv13 vs pcv10* | Oceania | 2-4-6 months | DTPa-HBV-Hib-TT/IPV/Rotavirus | 1.81 (1.47-2.24) |
| **27** | *pcv13 vs pcv10* | Africa | 6 weeks | DTPa-HBV-Hib-TT/IPV/Rotavirus | 0.37 (0.27-0.51) |
| **27** | *pcv13 vs pcv10* | Africa | 14 weeks | DTPa-HBV-Hib-TT/IPV/Rotavirus | 0.82 (0.61-1.11) |
| **27** | *pcv13 vs pcv10* | Africa | 6-14 weeks | DTPa-HBV-Hib-TT/IPV/Rotavirus | 1.04 (0.81-1.34) |

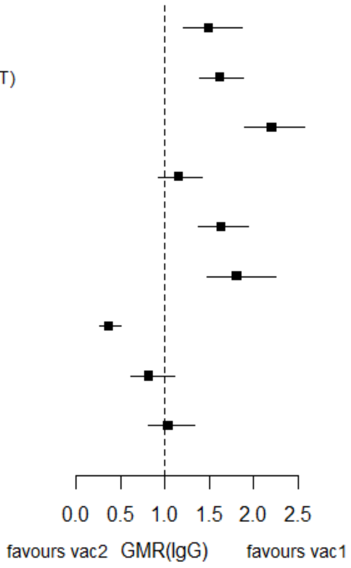

GMR: Geometric mean ratio; pcv: Pneumococcal conjugate vaccine; DTaP – diphtheria and tetanus toxoids, and acellular pertussis vaccine; DTwP – diphtheria and tetanus toxoids, and whole-cell pertussis vaccine; Hib-TT – Haemophilus influenzae type b vaccine (tetanus toxoid conjugate); HB – Hepatitis B vaccine; IPV – Inactivated polio vaccine; OPV – Oral polio vaccine; MenC – Meningococcal C vaccine; TT – tetanus toxoid conjugate; NA: not applicable; enr 3-6 m, 4-8 w int: enrolment at 3-6 months of age, and at 4-8 weeks interval of a 3 doses primary vaccines in total.

Each solid line in the figure shows the GMR from each trial. Black boxes and lines show the point estimates and confidence intervals for geometric mean ratios comparing vac1 vs vac2. Concomitant vaccines are vaccines co-administered with PCV primary vaccine series. Information on co-administered vaccine is not always available. Concomitant vaccines in the bracket are those administered in some but not all of the study sites.

**Supplementary Figure 12. Trial level geometric mean ratios for serotypes 5 at post-primary vaccination series.**

| ID | Comparison vac1 vs vac2 | Continent | Schedule-prime | Concomitant vaccines | GMR (95%CI) |
|---|---|---|---|---|---|
| **4** | *pcv13 vs pcv10* | Europe | 2-3-4 months | DTPa-HBV-Hib-TT/IPV | 1.23 (0.97-1.56) |
| **5** | *pcv13 vs pcv10* | Europe | 2–3–4 months | DTPa-HBV-Hib-TT/IPV + (MenC-TT) | 1.30 (1.12-1.51) |
| **6** | *pcv13 vs pcv10* | Asia | 2-4 months | DTPa-HBV-Hib-TT/IPV | 1.88 (1.66-2.13) |
| **18** | *pcv13 vs pcv10* | Oceania | 1-2-3 months | DTPw-HBV-Hib-TT + OPV | 1.01 (0.85-1.20) |
| **25** | *pcv13 vs pcv10* | Africa | 2–3–4 months | DTPw-HBV-Hib-TT + OPV | 1.32 (1.10-1.59) |
| **26** | *pcv13 vs pcv10* | Oceania | 2-4-6 months | DTPa-HBV-Hib-TT/IPV/Rotavirus | 2.49 (2.01-3.08) |
| **27** | *pcv13 vs pcv10* | Africa | 6 weeks | DTPa-HBV-Hib-TT/IPV/Rotavirus | 0.36 (0.25-0.50) |
| **27** | *pcv13 vs pcv10* | Africa | 14 weeks | DTPa-HBV-Hib-TT/IPV/Rotavirus | 0.53 (0.37-0.75) |
| **27** | *pcv13 vs pcv10* | Africa | 6-14 weeks | DTPa-HBV-Hib-TT/IPV/Rotavirus | 0.94 (0.71-1.24) |

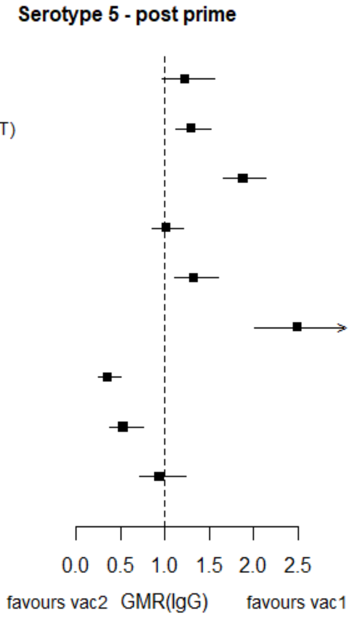

GMR: Geometric mean ratio; pcv: Pneumococcal conjugate vaccine; DTaP – diphtheria and tetanus toxoids, and acellular pertussis vaccine; DTwP – diphtheria and tetanus toxoids, and whole-cell pertussis vaccine; Hib-TT – Haemophilus influenzae type b vaccine (tetanus toxoid conjugate); HB – Hepatitis B vaccine; IPV – Inactivated polio vaccine; OPV – Oral polio vaccine; MenC – Meningococcal C vaccine; TT – tetanus toxoid conjugate; NA: not applicable; enr 3-6 m, 4-8 w int: enrolment at 3-6 months of age, and at 4-8 weeks interval of a 3 doses primary vaccines in total.

Each solid line in the figure shows the GMR from each trial. Black boxes and lines show the point estimates and confidence intervals for geometric mean ratios comparing vac1 vs vac2. Concomitant vaccines are vaccines co-administered with PCV primary vaccine series. Information on co-administered vaccine is not always available. Concomitant vaccines in the bracket are those administered in some but not all of the study sites.

**Supplementary Figure 13. Trial level geometric mean ratios for serotypes 7F at post-primary vaccination series.**

| ID | Comparison vac1 vs vac2 | Continent | Schedule-prime | Concomitant vaccines | GMR (95%CI) |
|---|---|---|---|---|---|
| **4** | *pcv13 vs pcv10* | Europe | 2-3-4 months | DTPa-HBV-Hib-TT/IPV | 1.37 (1.12-1.67) |
| **5** | *pcv13 vs pcv10* | Europe | 2–3–4 months | DTPa-HBV-Hib-TT/IPV + (MenC-TT) | 1.44 (1.26-1.64) |
| **6** | *pcv13 vs pcv10* | Asia | 2-4 months | DTPa-HBV-Hib-TT/IPV | 1.60 (1.42-1.82) |
| **18** | *pcv13 vs pcv10* | Oceania | 1-2-3 months | DTPw-HBV-Hib-TT + OPV | 1.29 (1.08-1.55) |
| **25** | *pcv13 vs pcv10* | Africa | 2–3–4 months | DTPw-HBV-Hib-TT + OPV | 1.29 (1.09-1.53) |
| **26** | *pcv13 vs pcv10* | Oceania | 2-4-6 months | DTPa-HBV-Hib-TT/IPV/Rotavirus | 1.72 (1.41-2.10) |
| **27** | *pcv13 vs pcv10* | Africa | 6 weeks | DTPa-HBV-Hib-TT/IPV/Rotavirus | 0.75 (0.52-1.09) |
| **27** | *pcv13 vs pcv10* | Africa | 14 weeks | DTPa-HBV-Hib-TT/IPV/Rotavirus | 1.18 (0.83-1.67) |
| **27** | *pcv13 vs pcv10* | Africa | 6-14 weeks | DTPa-HBV-Hib-TT/IPV/Rotavirus | 1.37 (1.02-1.84) |

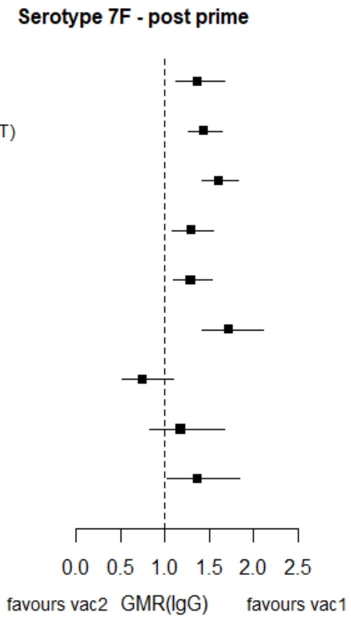

GMR: Geometric mean ratio; pcv: Pneumococcal conjugate vaccine; DTaP – diphtheria and tetanus toxoids, and acellular pertussis vaccine; DTwP – diphtheria and tetanus toxoids, and whole-cell pertussis vaccine; Hib-TT – Haemophilus influenzae type b vaccine (tetanus toxoid conjugate); HB – Hepatitis B vaccine; IPV – Inactivated polio vaccine; OPV – Oral polio vaccine; MenC – Meningococcal C vaccine; TT – tetanus toxoid conjugate; NA: not applicable; enr 3-6 m, 4-8 w int: enrolment at 3-6 months of age, and at 4-8 weeks interval of a 3 doses primary vaccines in total.

Each solid line in the figure shows the GMR from each trial. Black boxes and lines show the point estimates and confidence intervals for geometric mean ratios comparing vac1 vs vac2. Concomitant vaccines are vaccines co-administered with PCV primary vaccine series. Information on co-administered vaccine is not always available. Concomitant vaccines in the bracket are those administered in some but not all of the study sites.

**Supplementary Figure 14. Trial level geometric mean ratios for serotypes 3 at post-primary vaccination series.**

| ID | Comparison vac1 vs vac2 | Continent | Schedule-prime | Concomitant vaccines | GMR (95%CI) |
|---|---|---|---|---|---|
| **4** | *pcv13 vs pcv10* | Europe | 2-3-4 months | DTPa-HBV-Hib-TT/IPV | 43.33 (33.78-55.58) |
| **5** | *pcv13 vs pcv10* | Europe | 2–3–4 months | DTPa-HBV-Hib-TT/IPV + (MenC-TT) | 44.21 (38.12-51.27) |
| **6** | *pcv13 vs pcv10* | Asia | 2-4 months | DTPa-HBV-Hib-TT/IPV | 15.16 (13.17-17.46) |
| **18** | *pcv13 vs pcv10* | Oceania | 1-2-3 months | DTPw-HBV-Hib-TT + OPV | 2.19 (1.72-2.78) |
| **25** | *pcv13 vs pcv10* | Africa | 2–3–4 months | DTPw-HBV-Hib-TT + OPV | 90.36 (72.44-112.73) |
| **26** | *pcv13 vs pcv10* | Oceania | 2-4-6 months | DTPa-HBV-Hib-TT/IPV/Rotavirus | 5.60 (4.55-6.90) |
| **27** | *pcv13 vs pcv10* | Africa | 6 weeks | DTPa-HBV-Hib-TT/IPV/Rotavirus | 4.84 (3.92-5.99) |
| **27** | *pcv13 vs pcv10* | Africa | 14 weeks | DTPa-HBV-Hib-TT/IPV/Rotavirus | 11.99 (9.53-15.10) |
| **27** | *pcv13 vs pcv10* | Africa | 6-14 weeks | DTPa-HBV-Hib-TT/IPV/Rotavirus | 7.50 (5.88-9.57) |

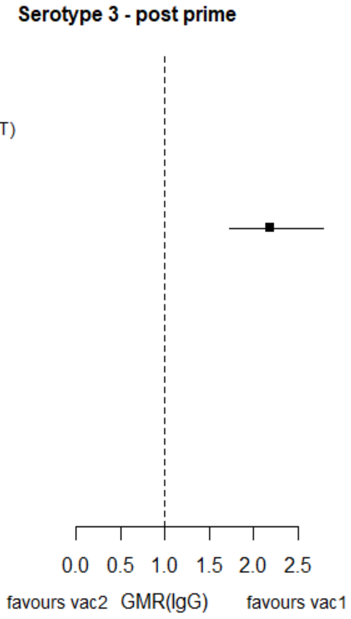

GMR: Geometric mean ratio; pcv: Pneumococcal conjugate vaccine; DTaP – diphtheria and tetanus toxoids, and acellular pertussis vaccine; DTwP – diphtheria and tetanus toxoids, and whole-cell pertussis vaccine; Hib-TT – Haemophilus influenzae type b vaccine (tetanus toxoid conjugate); HB – Hepatitis B vaccine; IPV – Inactivated polio vaccine; OPV – Oral polio vaccine; MenC – Meningococcal C vaccine; TT – tetanus toxoid conjugate; NA: not applicable; enr 3-6 m, 4-8 w int: enrolment at 3-6 months of age, and at 4-8 weeks interval of a 3 doses primary vaccines in total.

Each solid line in the figure shows the GMR from each trial. Black boxes and lines show the point estimates and confidence intervals for geometric mean ratios comparing vac1 vs vac2. Concomitant vaccines are vaccines co-administered with PCV primary vaccine series. Information on co-administered vaccine is not always available. Concomitant vaccines in the bracket are those administered in some but not all of the study sites.

**Supplementary Figure 15. Trial level geometric mean ratios for serotypes 6A at post-primary vaccination series.**

| ID | Comparison vac1 vs vac2 | Continent | Schedule-prime | Concomitant vaccines | GMR (95%CI) |
|---|---|---|---|---|---|
| **4** | *pcv13 vs pcv10* | Europe | 2-3-4 months | DTPa-HBV-Hib-TT/IPV | 18.67 (14.07-24.76) |
| **5** | *pcv13 vs pcv10* | Europe | 2–3–4 months | DTPa-HBV-Hib-TT/IPV + (MenC-TT) | 17.14 (13.99-20.99) |
| **6** | *pcv13 vs pcv10* | Asia | 2-4 months | DTPa-HBV-Hib-TT/IPV | 6.19 (5.21-7.36) |
| **18** | *pcv13 vs pcv10* | Oceania | 1-2-3 months | DTPw-HBV-Hib-TT + OPV | 3.54 (2.73-4.60) |
| **25** | *pcv13 vs pcv10* | Africa | 2–3–4 months | DTPw-HBV-Hib-TT + OPV | 23.17 (18.75-28.65) |
| **26** | *pcv13 vs pcv10* | Oceania | 2-4-6 months | DTPa-HBV-Hib-TT/IPV/Rotavirus | 12.47 (9.76-15.94) |
| **27** | *pcv13 vs pcv10* | Africa | 6 weeks | DTPa-HBV-Hib-TT/IPV/Rotavirus | 1.00 (0.78-1.28) |
| **27** | *pcv13 vs pcv10* | Africa | 14 weeks | DTPa-HBV-Hib-TT/IPV/Rotavirus | 1.18 (0.97-1.45) |
| **27** | *pcv13 vs pcv10* | Africa | 6-14 weeks | DTPa-HBV-Hib-TT/IPV/Rotavirus | 10.77 (7.74-14.96) |

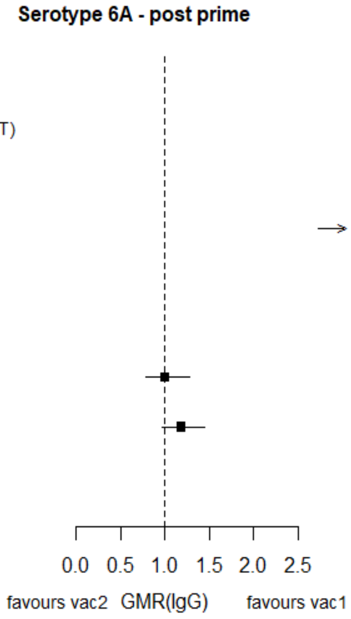

GMR: Geometric mean ratio; pcv: Pneumococcal conjugate vaccine; DTaP – diphtheria and tetanus toxoids, and acellular pertussis vaccine; DTwP – diphtheria and tetanus toxoids, and whole-cell pertussis vaccine; Hib-TT – Haemophilus influenzae type b vaccine (tetanus toxoid conjugate); HB – Hepatitis B vaccine; IPV – Inactivated polio vaccine; OPV – Oral polio vaccine; MenC – Meningococcal C vaccine; TT – tetanus toxoid conjugate; NA: not applicable; enr 3-6 m, 4-8 w int: enrolment at 3-6 months of age, and at 4-8 weeks interval of a 3 doses primary vaccines in total.

Each solid line in the figure shows the GMR from each trial. Black boxes and lines show the point estimates and confidence intervals for geometric mean ratios comparing vac1 vs vac2. Concomitant vaccines are vaccines co-administered with PCV primary vaccine series. Information on co-administered vaccine is not always available. Concomitant vaccines in the bracket are those administered in some but not all of the study sites.

**Supplementary Figure 16. Trial level geometric mean ratios for serotypes 19A at post-primary vaccination series.**

| ID | Comparison vac1 vs vac2 | Continent | Schedule-prime | Concomitant vaccines | GMR (95%CI) |
|---|---|---|---|---|---|
| 4 | *pcv13 vs pcv10* | Europe | 2-3-4 months | DTPa-HBV-Hib-TT/IPV | 16.30 (12.77-20.80) |
| 5 | *pcv13 vs pcv10* | Europe | 2–3–4 months | DTPa-HBV-Hib-TT/IPV + (MenC-TT) | 14.23 (11.70-17.32) |
| 6 | *pcv13 vs pcv10* | Asia | 2-4 months | DTPa-HBV-Hib-TT/IPV | 6.93 (5.81-8.26) |
| 18 | *pcv13 vs pcv10* | Oceania | 1-2-3 months | DTPw-HBV-Hib-TT + OPV | 4.78 (3.78-6.04) |
| 25 | *pcv13 vs pcv10* | Africa | 2–3–4 months | DTPw-HBV-Hib-TT + OPV | 22.91 (17.79-29.50) |
| 26 | *pcv13 vs pcv10* | Oceania | 2-4-6 months | DTPa-HBV-Hib-TT/IPV/Rotavirus | 3.59 (2.78-4.65) |
| 27 | *pcv13 vs pcv10* | Africa | 6 weeks | DTPa-HBV-Hib-TT/IPV/Rotavirus | 1.30 (0.99-1.71) |
| 27 | *pcv13 vs pcv10* | Africa | 14 weeks | DTPa-HBV-Hib-TT/IPV/Rotavirus | 2.50 (1.90-3.29) |
| 27 | *pcv13 vs pcv10* | Africa | 6-14 weeks | DTPa-HBV-Hib-TT/IPV/Rotavirus | 11.04 (8.23-14.80) |

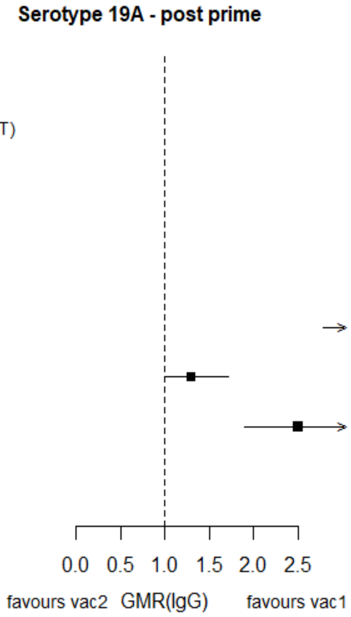

GMR: Geometric mean ratio; pcv: Pneumococcal conjugate vaccine; DTaP – diphtheria and tetanus toxoids, and acellular pertussis vaccine; DTwP – diphtheria and tetanus toxoids, and whole-cell pertussis vaccine; Hib-TT – Haemophilus influenzae type b vaccine (tetanus toxoid conjugate); HB – Hepatitis B vaccine; IPV – Inactivated polio vaccine; OPV – Oral polio vaccine; MenC – Meningococcal C vaccine; TT – tetanus toxoid conjugate; NA: not applicable; enr 3-6 m, 4-8 w int: enrolment at 3-6 months of age, and at 4-8 weeks interval of a 3 doses primary vaccines in total.

Each solid line in the figure shows the GMR from each trial. Black boxes and lines show the point estimates and confidence intervals for geometric mean ratios comparing vac1 vs vac2. Concomitant vaccines are vaccines co-administered with PCV primary vaccine series. Information on co-administered vaccine is not always available. Concomitant vaccines in the bracket are those administered in some but not all of the study sites.

**Supplementary Figure 17. Trial level geometric mean ratios for serotypes 4 at pre-booster.**

| ID | Comparison vac1 vs vac2 | Continent | Schedule-prime | Concomitant vaccines | GMR (95%CI) |
|---|---|---|---|---|---|
| 1 | *pcv7 vs pcv10* | Asia | 6-10-24 weeks | DTPw-HBV-Hib-TT + OPV | 0.60 (0.43-0.84) |
| 1 | *pcv7 vs pcv10* | Europe | 2-4-6 months | DTPw-HBV-Hib-TT + IPV | 0.91 (0.68-1.20) |
| 3 | *pcv7 vs pcv10* | Europe | 2-3-4 months | NA | 1.02 (0.74-1.42) |
| 7 | *pcv7 vs pcv10* | Europe | 2-3-4 months | DTPa-(HBV)-Hib-TT/IPV | 0.92 (0.79-1.08) |
| 8 | *pcv7 vs pcv10* | Europe | 2-3-4 months | DTPa-(HBV)-Hib-TT/IPV | 1.08 (0.94-1.25) |
| 9 | *pcv7 vs pcv10* | Europe | 2-4-6 months | DTPa-(HBV)-Hib-IPV + MenC | 1.14 (0.94-1.38) |
| 4 | *pcv13 vs pcv10* | Europe | 2-3-4 months | DTPa-HBV-Hib-TT/IPV | 0.68 (0.54-0.84) |
| 5 | *pcv13 vs pcv10* | Europe | 2–3–4 months | DTPa-HBV-Hib-TT/IPV + (MenC-TT) | 0.83 (0.71-0.96) |
| 6 | *pcv13 vs pcv10* | Asia | 2-4 months | DTPa-HBV-Hib-TT/IPV | 1.04 (0.90-1.20) |
| 18 | *pcv13 vs pcv10* | Oceania | 1-2-3 months | Measles, PPV | 0.91 (0.74-1.12) |
| 27 | *pcv13 vs pcv10* | Africa | 6 weeks | NA | 0.29 (0.21-0.40) |
| 27 | *pcv13 vs pcv10* | Africa | 14 weeks | NA | 0.48 (0.34-0.67) |
| 27 | *pcv13 vs pcv10* | Africa | 6-14 weeks | NA | 0.52 (0.38-0.71) |
| 13 | *pcv13 vs pcv7* | Europe | 2-3-4 months | DTPa-Hib-TT/IPV | 0.68 (0.59-0.80) |
| 15 | *pcv13 vs pcv7* | Europe | 2-3-4 months | DTPa-HBV-Hib-TT/IPV | 0.81 (0.70-0.92) |
| 19 | *pcv13 vs pcv7* | Europe | 2-4 months | Hib-MenC-TT | 0.61 (0.46-0.80) |
| 22 | *pcv13 vs pcv7* | North America | 2-4-6 months | MMRV, Hib, HAV | 0.68 (0.58-0.79) |

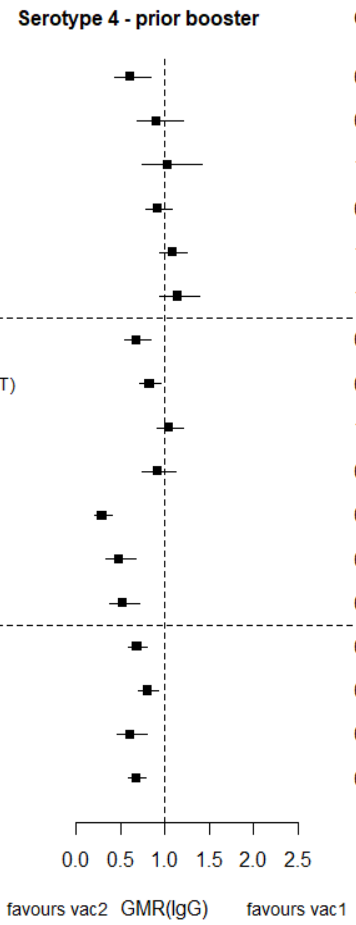

GMR: Geometric mean ratio; pcv: Pneumococcal conjugate vaccine; DTaP – diphtheria and tetanus toxoids, and acellular pertussis vaccine; DTwP – diphtheria and tetanus toxoids, and whole-cell pertussis vaccine; Hib-TT – Haemophilus influenzae type b vaccine (tetanus toxoid conjugate); HB – Hepatitis B vaccine; IPV – Inactivated polio vaccine; OPV – Oral polio vaccine; MenC – Meningococcal C vaccine; TT – tetanus toxoid conjugate; NA: not applicable; enr 3-6 m, 4-8 w int: enrolment at 3-6 months of age, and at 4-8 weeks interval of a 3 doses primary vaccines in total.

Each solid line in the figure shows the GMR from each trial. Black boxes and lines show the point estimates and confidence intervals for geometric mean ratios comparing vac1 vs vac2. Concomitant vaccines are vaccines co-administered with PCV primary vaccine series. Information on co-administered vaccine is not always available. Concomitant vaccines in the bracket are those administered in some but not all of the study sites.

**Supplementary Figure 18. Trial level geometric mean ratios for serotypes 6B at pre-booster.**

| ID | Comparison vac1 vs vac2 | Continent | Schedule-prime | Concomitant vaccines | Serotype 6B - prior booster | GMR (95%CI) |
|---|---|---|---|---|---|---|
| 1 | *pcv7 vs pcv10* | Asia | 6-10-24 weeks | DTPw-HBV-Hib-TT + OPV | | 0.55 (0.32-0.95) |
| 1 | *pcv7 vs pcv10* | Europe | 2-4-6 months | DTPw-HBV-Hib-TT + IPV | | 1.01 (0.59-1.72) |
| 3 | *pcv7 vs pcv10* | Europe | 2-3-4 months | NA | | 0.84 (0.56-1.25) |
| 7 | *pcv7 vs pcv10* | Europe | 2-3-4 months | DTPa-(HBV)-Hib-TT/IPV | | 0.41 (0.33-0.51) |
| 8 | *pcv7 vs pcv10* | Europe | 2-3-4 months | DTPa-(HBV)-Hib-TT/IPV | | 0.59 (0.48-0.72) |
| 9 | *pcv7 vs pcv10* | Europe | 2-4-6 months | DTPa-(HBV)-Hib-IPV + MenC | | 0.67 (0.52-0.85) |
| 4 | *pcv13 vs pcv10* | Europe | 2-3-4 months | DTPa-HBV-Hib-TT/IPV | | 0.57 (0.44-0.74) |
| 5 | *pcv13 vs pcv10* | Europe | 2–3–4 months | DTPa-HBV-Hib-TT/IPV + (MenC-TT) | | 0.38 (0.31-0.46) |
| 6 | *pcv13 vs pcv10* | Asia | 2-4 months | DTPa-HBV-Hib-TT/IPV | | 0.39 (0.33-0.45) |
| 18 | *pcv13 vs pcv10* | Oceania | 1-2-3 months | Measles, PPV | | 0.78 (0.62-0.99) |
| 27 | *pcv13 vs pcv10* | Africa | 6 weeks | NA | | 0.23 (0.18-0.31) |
| 27 | *pcv13 vs pcv10* | Africa | 14 weeks | NA | | 0.18 (0.13-0.24) |
| 27 | *pcv13 vs pcv10* | Africa | 6-14 weeks | NA | | 0.25 (0.18-0.35) |
| 13 | *pcv13 vs pcv7* | Europe | 2-3-4 months | DTPa-Hib-TT/IPV | | 0.92 (0.76-1.12) |
| 15 | *pcv13 vs pcv7* | Europe | 2-3-4 months | DTPa-HBV-Hib-TT/IPV | | 0.92 (0.79-1.07) |
| 19 | *pcv13 vs pcv7* | Europe | 2-4 months | Hib-MenC-TT | | 1.35 (0.86-2.12) |
| 22 | *pcv13 vs pcv7* | North America | 2-4-6 months | MMRV, Hib, HAV | | 0.77 (0.64-0.91) |

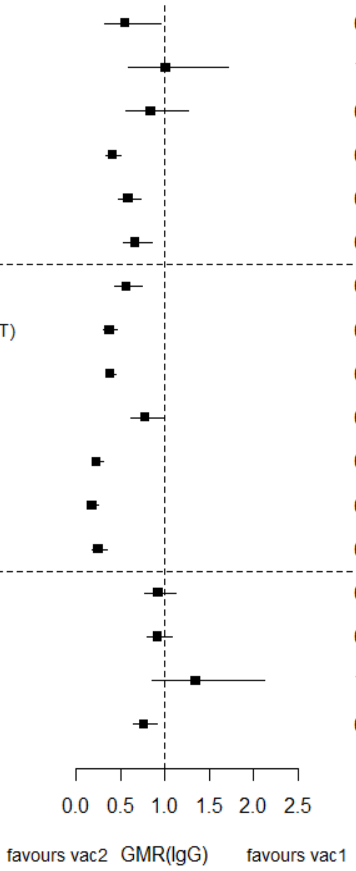

GMR: Geometric mean ratio; pcv: Pneumococcal conjugate vaccine; DTaP – diphtheria and tetanus toxoids, and acellular pertussis vaccine; DTwP – diphtheria and tetanus toxoids, and whole-cell pertussis vaccine; Hib-TT – Haemophilus influenzae type b vaccine (tetanus toxoid conjugate); HB – Hepatitis B vaccine; IPV – Inactivated polio vaccine; OPV – Oral polio vaccine; MenC – Meningococcal C vaccine; TT – tetanus toxoid conjugate; NA: not applicable; enr 3-6 m, 4-8 w int: enrolment at 3-6 months of age, and at 4-8 weeks interval of a 3 doses primary vaccines in total.

Each solid line in the figure shows the GMR from each trial. Black boxes and lines show the point estimates and confidence intervals for geometric mean ratios comparing vac1 vs vac2. Concomitant vaccines are vaccines co-administered with PCV primary vaccine series. Information on co-administered vaccine is not always available. Concomitant vaccines in the bracket are those administered in some but not all of the study sites.

**Supplementary Figure 19. Trial level geometric mean ratios for serotypes 9V at pre-booster.**

| ID | Comparison vac1 vs vac2 | Continent | Schedule-prime | Concomitant vaccines | GMR (95%CI) |
|---|---|---|---|---|---|
| 1 | *pcv7 vs pcv10* | Asia | 6-10-24 weeks | DTPw-HBV-Hib-TT + OPV | 0.64 (0.48-0.85) |
| 1 | *pcv7 vs pcv10* | Europe | 2-4-6 months | DTPw-HBV-Hib-TT + IPV | 1.04 (0.80-1.34) |
| 3 | *pcv7 vs pcv10* | Europe | 2-3-4 months | NA | 1.36 (1.04-1.77) |
| 7 | *pcv7 vs pcv10* | Europe | 2-3-4 months | DTPa-(HBV)-Hib-TT/IPV | 0.75 (0.64-0.87) |
| 8 | *pcv7 vs pcv10* | Europe | 2-3-4 months | DTPa-(HBV)-Hib-TT/IPV | 1.17 (1.02-1.33) |
| 9 | *pcv7 vs pcv10* | Europe | 2-4-6 months | DTPa-(HBV)-Hib-IPV + MenC | 1.19 (0.98-1.43) |
| 4 | *pcv13 vs pcv10* | Europe | 2-3-4 months | DTPa-HBV-Hib-TT/IPV | 0.60 (0.48-0.74) |
| 5 | *pcv13 vs pcv10* | Europe | 2–3–4 months | DTPa-HBV-Hib-TT/IPV + (MenC-TT) | 0.74 (0.64-0.86) |
| 6 | *pcv13 vs pcv10* | Asia | 2-4 months | DTPa-HBV-Hib-TT/IPV | 1.21 (1.06-1.39) |
| 18 | *pcv13 vs pcv10* | Oceania | 1-2-3 months | Measles, PPV | 0.97 (0.80-1.17) |
| 27 | *pcv13 vs pcv10* | Africa | 6 weeks | NA | 0.34 (0.26-0.45) |
| 27 | *pcv13 vs pcv10* | Africa | 14 weeks | NA | 0.65 (0.47-0.89) |
| 27 | *pcv13 vs pcv10* | Africa | 6-14 weeks | NA | 0.79 (0.58-1.09) |
| 13 | *pcv13 vs pcv7* | Europe | 2-3-4 months | DTPa-Hib-TT/IPV | 0.80 (0.68-0.93) |
| 15 | *pcv13 vs pcv7* | Europe | 2-3-4 months | DTPa-HBV-Hib-TT/IPV | 0.88 (0.79-1.00) |
| 19 | *pcv13 vs pcv7* | Europe | 2-4 months | Hib-MenC-TT | 0.87 (0.63-1.19) |
| 22 | *pcv13 vs pcv7* | North America | 2-4-6 months | MMRV, Hib, HAV | 0.71 (0.62-0.81) |

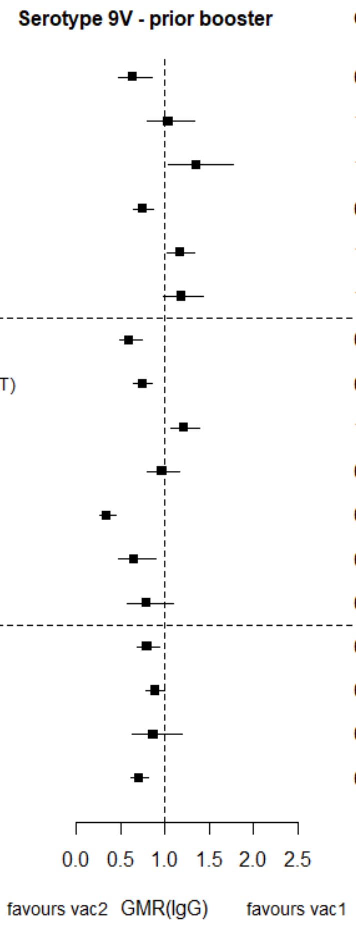

GMR: Geometric mean ratio; pcv: Pneumococcal conjugate vaccine; DTaP – diphtheria and tetanus toxoids, and acellular pertussis vaccine; DTwP – diphtheria and tetanus toxoids, and whole-cell pertussis vaccine; Hib-TT – Haemophilus influenzae type b vaccine (tetanus toxoid conjugate); HB – Hepatitis B vaccine; IPV – Inactivated polio vaccine; OPV – Oral polio vaccine; MenC – Meningococcal C vaccine; TT – tetanus toxoid conjugate; NA: not applicable; enr 3-6 m, 4-8 w int: enrolment at 3-6 months of age, and at 4-8 weeks interval of a 3 doses primary vaccines in total.

Each solid line in the figure shows the GMR from each trial. Black boxes and lines show the point estimates and confidence intervals for geometric mean ratios comparing vac1 vs vac2. Concomitant vaccines are vaccines co-administered with PCV primary vaccine series. Information on co-administered vaccine is not always available. Concomitant vaccines in the bracket are those administered in some but not all of the study sites.

**Supplementary Figure 20. Trial level geometric mean ratios for serotypes 14 at pre-booster.**

| ID | Comparison vac1 vs vac2 | Continent | Schedule-prime | Concomitant vaccines | GMR (95%CI) |
|---|---|---|---|---|---|
| 1 | *pcv7 vs pcv10* | Asia | 6-10-24 weeks | DTPw-HBV-Hib-TT + OPV | 1.33 (0.89-1.99) |
| 1 | *pcv7 vs pcv10* | Europe | 2-4-6 months | DTPw-HBV-Hib-TT + IPV | 1.29 (0.89-1.86) |
| 3 | *pcv7 vs pcv10* | Europe | 2-3-4 months | NA | 1.93 (1.30-2.86) |
| 7 | *pcv7 vs pcv10* | Europe | 2-3-4 months | DTPa-(HBV)-Hib-TT/IPV | 1.41 (1.17-1.69) |
| 8 | *pcv7 vs pcv10* | Europe | 2-3-4 months | DTPa-(HBV)-Hib-TT/IPV | 1.64 (1.37-1.97) |
| 9 | *pcv7 vs pcv10* | Europe | 2-4-6 months | DTPa-(HBV)-Hib-IPV + MenC | 1.90 (1.56-2.32) |
| 4 | *pcv13 vs pcv10* | Europe | 2-3-4 months | DTPa-HBV-Hib-TT/IPV | 1.33 (1.02-1.73) |
| 5 | *pcv13 vs pcv10* | Europe | 2–3–4 months | DTPa-HBV-Hib-TT/IPV + (MenC-TT) | 1.49 (1.22-1.83) |
| 6 | *pcv13 vs pcv10* | Asia | 2-4 months | DTPa-HBV-Hib-TT/IPV | 1.30 (1.07-1.58) |
| 18 | *pcv13 vs pcv10* | Oceania | 1-2-3 months | Measles, PPV | 1.25 (0.94-1.65) |
| 27 | *pcv13 vs pcv10* | Africa | 6 weeks | NA | 0.82 (0.57-1.19) |
| 27 | *pcv13 vs pcv10* | Africa | 14 weeks | NA | 0.77 (0.51-1.18) |
| 27 | *pcv13 vs pcv10* | Africa | 6-14 weeks | NA | 1.13 (0.70-1.82) |
| 13 | *pcv13 vs pcv7* | Europe | 2-3-4 months | DTPa-Hib-TT/IPV | 0.87 (0.72-1.05) |
| 15 | *pcv13 vs pcv7* | Europe | 2-3-4 months | DTPa-HBV-Hib-TT/IPV | 0.84 (0.71-0.99) |
| 19 | *pcv13 vs pcv7* | Europe | 2-4 months | Hib-MenC-TT | 1.10 (0.75-1.60) |
| 22 | *pcv13 vs pcv7* | North America | 2-4-6 months | MMRV, Hib, HAV | 0.77 (0.64-0.92) |

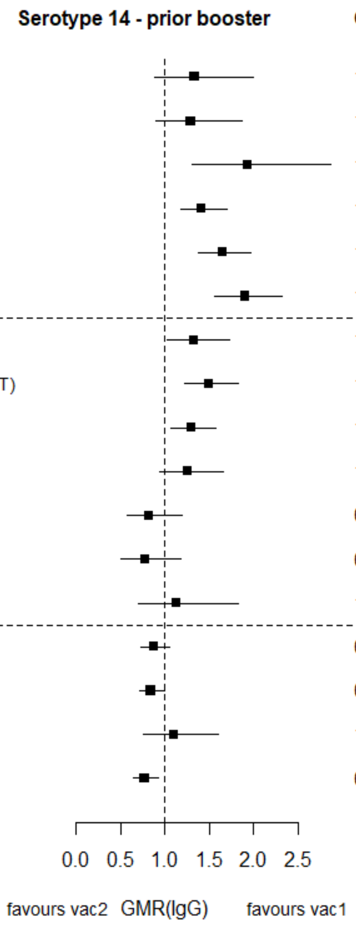

GMR: Geometric mean ratio; pcv: Pneumococcal conjugate vaccine; DTaP – diphtheria and tetanus toxoids, and acellular pertussis vaccine; DTwP – diphtheria and tetanus toxoids, and whole-cell pertussis vaccine; Hib-TT – Haemophilus influenzae type b vaccine (tetanus toxoid conjugate); HB – Hepatitis B vaccine; IPV – Inactivated polio vaccine; OPV – Oral polio vaccine; MenC – Meningococcal C vaccine; TT – tetanus toxoid conjugate; NA: not applicable; enr 3-6 m, 4-8 w int: enrolment at 3-6 months of age, and at 4-8 weeks interval of a 3 doses primary vaccines in total.

Each solid line in the figure shows the GMR from each trial. Black boxes and lines show the point estimates and confidence intervals for geometric mean ratios comparing vac1 vs vac2. Concomitant vaccines are vaccines co-administered with PCV primary vaccine series. Information on co-administered vaccine is not always available. Concomitant vaccines in the bracket are those administered in some but not all of the study sites.

**Supplementary Figure 21. Trial level geometric mean ratios for serotypes 18C at pre-booster.**

| ID | Comparison vac1 vs vac2 | Continent | Schedule-prime | Concomitant vaccines | GMR (95%CI) |
|---|---|---|---|---|---|
| 1 | *pcv7 vs pcv10* | Asia | 6-10-24 weeks | DTPw-HBV-Hib-TT + OPV | 0.30 (0.22-0.41) |
| 1 | *pcv7 vs pcv10* | Europe | 2-4-6 months | DTPw-HBV-Hib-TT + IPV | 0.65 (0.47-0.90) |
| 3 | *pcv7 vs pcv10* | Europe | 2-3-4 months | NA | 0.60 (0.44-0.81) |
| 7 | *pcv7 vs pcv10* | Europe | 2-3-4 months | DTPa-(HBV)-Hib-TT/IPV | 0.81 (0.68-0.96) |
| 8 | *pcv7 vs pcv10* | Europe | 2-3-4 months | DTPa-(HBV)-Hib-TT/IPV | 0.95 (0.83-1.09) |
| 9 | *pcv7 vs pcv10* | Europe | 2-4-6 months | DTPa-(HBV)-Hib-IPV + MenC | 0.83 (0.69-0.99) |
| 4 | *pcv13 vs pcv10* | Europe | 2-3-4 months | DTPa-HBV-Hib-TT/IPV | 0.84 (0.68-1.04) |
| 5 | *pcv13 vs pcv10* | Europe | 2–3–4 months | DTPa-HBV-Hib-TT/IPV + (MenC-TT) | 0.82 (0.70-0.97) |
| 6 | *pcv13 vs pcv10* | Asia | 2-4 months | DTPa-HBV-Hib-TT/IPV | 0.84 (0.73-0.96) |
| 18 | *pcv13 vs pcv10* | Oceania | 1-2-3 months | Measles, PPV | 0.68 (0.55-0.84) |
| 27 | *pcv13 vs pcv10* | Africa | 6 weeks | NA | 1.27 (0.88-1.84) |
| 27 | *pcv13 vs pcv10* | Africa | 14 weeks | NA | 1.08 (0.77-1.53) |
| 27 | *pcv13 vs pcv10* | Africa | 6-14 weeks | NA | 0.88 (0.64-1.20) |
| 13 | *pcv13 vs pcv7* | Europe | 2-3-4 months | DTPa-Hib-TT/IPV | 0.85 (0.74-0.99) |
| 15 | *pcv13 vs pcv7* | Europe | 2-3-4 months | DTPa-HBV-Hib-TT/IPV | 0.88 (0.78-0.99) |
| 19 | *pcv13 vs pcv7* | Europe | 2-4 months | Hib-MenC-TT | 0.98 (0.78-1.23) |
| 22 | *pcv13 vs pcv7* | North America | 2-4-6 months | MMRV, Hib, HAV | 0.73 (0.63-0.83) |

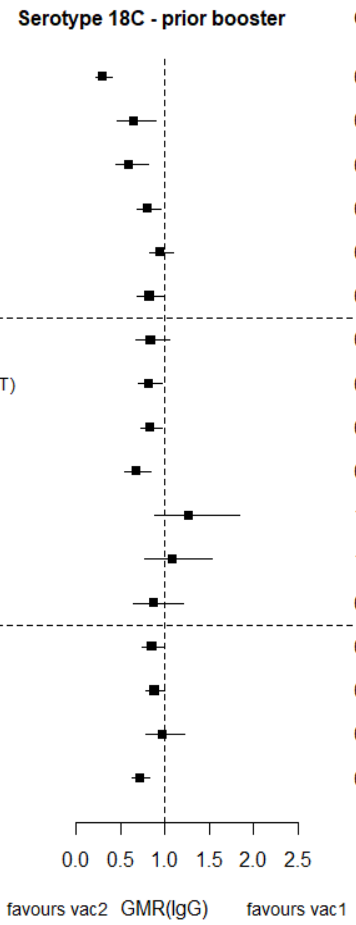

GMR: Geometric mean ratio; pcv: Pneumococcal conjugate vaccine; DTaP – diphtheria and tetanus toxoids, and acellular pertussis vaccine; DTwP – diphtheria and tetanus toxoids, and whole-cell pertussis vaccine; Hib-TT – Haemophilus influenzae type b vaccine (tetanus toxoid conjugate); HB – Hepatitis B vaccine; IPV – Inactivated polio vaccine; OPV – Oral polio vaccine; MenC – Meningococcal C vaccine; TT – tetanus toxoid conjugate; NA: not applicable; enr 3-6 m, 4-8 w int: enrolment at 3-6 months of age, and at 4-8 weeks interval of a 3 doses primary vaccines in total.

Each solid line in the figure shows the GMR from each trial. Black boxes and lines show the point estimates and confidence intervals for geometric mean ratios comparing vac1 vs vac2. Concomitant vaccines are vaccines co-administered with PCV primary vaccine series. Information on co-administered vaccine is not always available. Concomitant vaccines in the bracket are those administered in some but not all of the study sites.

**Supplementary Figure 22. Trial level geometric mean ratios for serotypes 19F at pre-booster.**

| ID | Comparison vac1 vs vac2 | Continent | Schedule-prime | Concomitant vaccines | GMR (95%CI) |
|---|---|---|---|---|---|
| 1 | *pcv7 vs pcv10* | Asia | 6-10-24 weeks | DTPw-HBV-Hib-TT + OPV | 0.11 (0.07-0.19) |
| 1 | *pcv7 vs pcv10* | Europe | 2-4-6 months | DTPw-HBV-Hib-TT + IPV | 0.32 (0.19-0.56) |
| 3 | *pcv7 vs pcv10* | Europe | 2-3-4 months | NA | 0.43 (0.26-0.71) |
| 7 | *pcv7 vs pcv10* | Europe | 2-3-4 months | DTPa-(HBV)-Hib-TT/IPV | 0.35 (0.28-0.45) |
| 8 | *pcv7 vs pcv10* | Europe | 2-3-4 months | DTPa-(HBV)-Hib-TT/IPV | 0.47 (0.38-0.59) |
| 9 | *pcv7 vs pcv10* | Europe | 2-4-6 months | DTPa-(HBV)-Hib-IPV + MenC | 0.25 (0.19-0.33) |
| 4 | *pcv13 vs pcv10* | Europe | 2-3-4 months | DTPa-HBV-Hib-TT/IPV | 0.49 (0.37-0.65) |
| 5 | *pcv13 vs pcv10* | Europe | 2–3–4 months | DTPa-HBV-Hib-TT/IPV + (MenC-TT) | 0.45 (0.37-0.54) |
| 6 | *pcv13 vs pcv10* | Asia | 2-4 months | DTPa-HBV-Hib-TT/IPV | 0.55 (0.48-0.62) |
| 18 | *pcv13 vs pcv10* | Oceania | 1-2-3 months | Measles, PPV | 0.75 (0.60-0.95) |
| 27 | *pcv13 vs pcv10* | Africa | 6 weeks | NA | 1.10 (0.74-1.63) |
| 27 | *pcv13 vs pcv10* | Africa | 14 weeks | NA | 0.88 (0.64-1.21) |
| 27 | *pcv13 vs pcv10* | Africa | 6-14 weeks | NA | 0.83 (0.57-1.22) |
| 13 | *pcv13 vs pcv7* | Europe | 2-3-4 months | DTPa-Hib-TT/IPV | 0.92 (0.71-1.19) |
| 15 | *pcv13 vs pcv7* | Europe | 2-3-4 months | DTPa-HBV-Hib-TT/IPV | 1.14 (0.96-1.35) |
| 19 | *pcv13 vs pcv7* | Europe | 2-4 months | Hib-MenC-TT | 1.33 (0.80-2.23) |
| 22 | *pcv13 vs pcv7* | North America | 2-4-6 months | MMRV, Hib, HAV | 1.11 (0.95-1.31) |

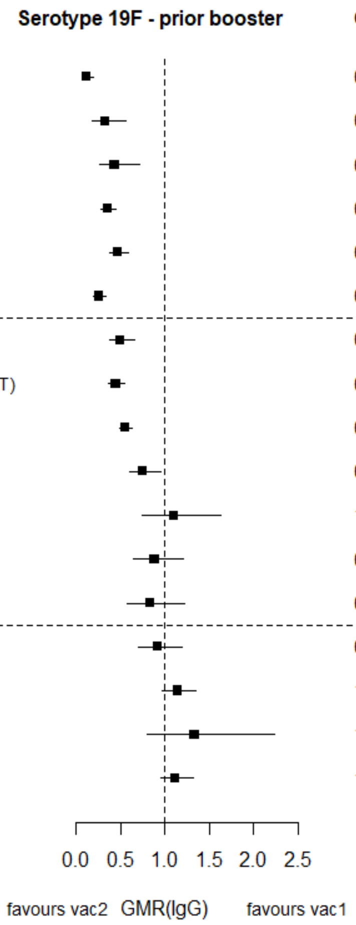

GMR: Geometric mean ratio; pcv: Pneumococcal conjugate vaccine; DTaP – diphtheria and tetanus toxoids, and acellular pertussis vaccine; DTwP – diphtheria and tetanus toxoids, and whole-cell pertussis vaccine; Hib-TT – Haemophilus influenzae type b vaccine (tetanus toxoid conjugate); HB – Hepatitis B vaccine; IPV – Inactivated polio vaccine; OPV – Oral polio vaccine; MenC – Meningococcal C vaccine; TT – tetanus toxoid conjugate; NA: not applicable; enr 3-6 m, 4-8 w int: enrolment at 3-6 months of age, and at 4-8 weeks interval of a 3 doses primary vaccines in total.

Each solid line in the figure shows the GMR from each trial. Black boxes and lines show the point estimates and confidence intervals for geometric mean ratios comparing vac1 vs vac2. Concomitant vaccines are vaccines co-administered with PCV primary vaccine series. Information on co-administered vaccine is not always available. Concomitant vaccines in the bracket are those administered in some but not all of the study sites.

**Supplementary Figure 23. Trial level geometric mean ratios for serotypes 23F at pre-booster.**

| ID | Comparison vac1 vs vac2 | Continent | Schedule-prime | Concomitant vaccines | GMR (95%CI) |
|---|---|---|---|---|---|
| **1** | *pcv7 vs pcv10* | Asia | 6-10-24 weeks | DTPw-HBV-Hib-TT + OPV | 0.46 (0.29-0.73) |
| **1** | *pcv7 vs pcv10* | Europe | 2-4-6 months | DTPw-HBV-Hib-TT + IPV | 1.93 (1.30-2.86) |
| **3** | *pcv7 vs pcv10* | Europe | 2-3-4 months | NA | 0.72 (0.49-1.08) |
| **7** | *pcv7 vs pcv10* | Europe | 2-3-4 months | DTPa-(HBV)-Hib-TT/IPV | 0.71 (0.58-0.87) |
| **8** | *pcv7 vs pcv10* | Europe | 2-3-4 months | DTPa-(HBV)-Hib-TT/IPV | 0.98 (0.81-1.19) |
| **9** | *pcv7 vs pcv10* | Europe | 2-4-6 months | DTPa-(HBV)-Hib-IPV + MenC | 0.96 (0.75-1.23) |
| **4** | *pcv13 vs pcv10* | Europe | 2-3-4 months | DTPa-HBV-Hib-TT/IPV | 0.62 (0.48-0.80) |
| **5** | *pcv13 vs pcv10* | Europe | 2–3–4 months | DTPa-HBV-Hib-TT/IPV + (MenC-TT) | 0.70 (0.58-0.85) |
| **6** | *pcv13 vs pcv10* | Asia | 2-4 months | DTPa-HBV-Hib-TT/IPV | 0.67 (0.57-0.79) |
| **18** | *pcv13 vs pcv10* | Oceania | 1-2-3 months | Measles, PPV | 1.00 (0.77-1.30) |
| **27** | *pcv13 vs pcv10* | Africa | 6 weeks | NA | 0.50 (0.36-0.69) |
| **27** | *pcv13 vs pcv10* | Africa | 14 weeks | NA | 0.37 (0.28-0.50) |
| **27** | *pcv13 vs pcv10* | Africa | 6-14 weeks | NA | 0.46 (0.31-0.66) |
| **13** | *pcv13 vs pcv7* | Europe | 2-3-4 months | DTPa-Hib-TT/IPV | 0.79 (0.65-0.96) |
| **15** | *pcv13 vs pcv7* | Europe | 2-3-4 months | DTPa-HBV-Hib-TT/IPV | 0.86 (0.73-1.02) |
| **19** | *pcv13 vs pcv7* | Europe | 2-4 months | Hib-MenC-TT | 1.45 (0.96-2.19) |
| **22** | *pcv13 vs pcv7* | North America | 2-4-6 months | MMRV, Hib, HAV | 0.77 (0.64-0.92) |

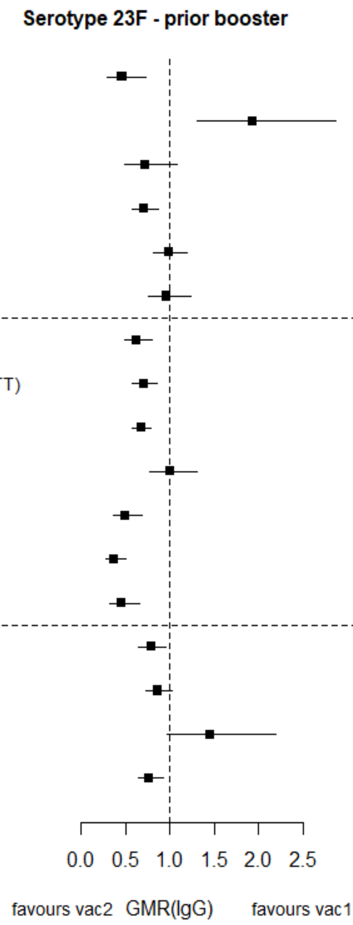

GMR: Geometric mean ratio; pcv: Pneumococcal conjugate vaccine; DTaP – diphtheria and tetanus toxoids, and acellular pertussis vaccine; DTwP – diphtheria and tetanus toxoids, and whole-cell pertussis vaccine; Hib-TT – Haemophilus influenzae type b vaccine (tetanus toxoid conjugate); HB – Hepatitis B vaccine; IPV – Inactivated polio vaccine; OPV – Oral polio vaccine; MenC – Meningococcal C vaccine; TT – tetanus toxoid conjugate; NA: not applicable; enr 3-6 m, 4-8 w int: enrolment at 3-6 months of age, and at 4-8 weeks interval of a 3 doses primary vaccines in total.

Each solid line in the figure shows the GMR from each trial. Black boxes and lines show the point estimates and confidence intervals for geometric mean ratios comparing vac1 vs vac2. Concomitant vaccines are vaccines co-administered with PCV primary vaccine series. Information on co-administered vaccine is not always available. Concomitant vaccines in the bracket are those administered in some but not all of the study sites.

**Supplementary Figure 24. Trial level geometric mean ratios for serotypes 1 at pre-booster.**

| ID | Comparison vac1 vs vac2 | Continent | Schedule-prime | Concomitant vaccines | GMR (95%CI) |
|---|---|---|---|---|---|
| **4** | *pcv13 vs pcv10* | Europe | 2-3-4 months | DTPa-HBV-Hib-TT/IPV | 1.81 (1.48-2.21) |
| **5** | *pcv13 vs pcv10* | Europe | 2–3–4 months | DTPa-HBV-Hib-TT/IPV + (MenC-TT) | 1.70 (1.48-1.96) |
| **6** | *pcv13 vs pcv10* | Asia | 2-4 months | DTPa-HBV-Hib-TT/IPV | 1.96 (1.71-2.25) |
| **18** | *pcv13 vs pcv10* | Oceania | 1-2-3 months | Measles, PPV | 1.44 (1.16-1.77) |
| **27** | *pcv13 vs pcv10* | Africa | 6 weeks | NA | 0.69 (0.48-0.99) |
| **27** | *pcv13 vs pcv10* | Africa | 14 weeks | NA | 0.89 (0.67-1.20) |
| **27** | *pcv13 vs pcv10* | Africa | 6-14 weeks | NA | 1.30 (0.98-1.72) |

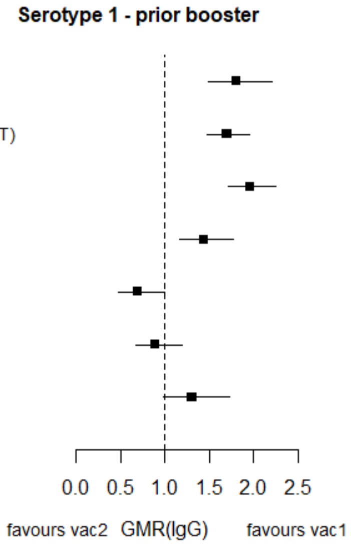

GMR: Geometric mean ratio; pcv: Pneumococcal conjugate vaccine; DTaP – diphtheria and tetanus toxoids, and acellular pertussis vaccine; DTwP – diphtheria and tetanus toxoids, and whole-cell pertussis vaccine; Hib-TT – Haemophilus influenzae type b vaccine (tetanus toxoid conjugate); HB – Hepatitis B vaccine; IPV – Inactivated polio vaccine; OPV – Oral polio vaccine; MenC – Meningococcal C vaccine; TT – tetanus toxoid conjugate; NA: not applicable; enr 3-6 m, 4-8 w int: enrolment at 3-6 months of age, and at 4-8 weeks interval of a 3 doses primary vaccines in total.

Each solid line in the figure shows the GMR from each trial. Black boxes and lines show the point estimates and confidence intervals for geometric mean ratios comparing vac1 vs vac2. Concomitant vaccines are vaccines co-administered with PCV primary vaccine series. Information on co-administered vaccine is not always available. Concomitant vaccines in the bracket are those administered in some but not all of the study sites.

**Supplementary Figure 25. Trial level geometric mean ratios for serotypes 5 at pre-booster.**

| ID | Comparison vac1 vs vac2 | Continent | Schedule-prime | Concomitant vaccines | GMR (95%CI) |
|---|---|---|---|---|---|
| **4** | *pcv13 vs pcv10* | Europe | 2-3-4 months | DTPa-HBV-Hib-TT/IPV | 1.51 (1.22-1.87) |
| **5** | *pcv13 vs pcv10* | Europe | 2–3–4 months | DTPa-HBV-Hib-TT/IPV + (MenC-TT) | 1.62 (1.40-1.87) |
| **6** | *pcv13 vs pcv10* | Asia | 2-4 months | DTPa-HBV-Hib-TT/IPV | 1.58 (1.40-1.79) |
| **18** | *pcv13 vs pcv10* | Oceania | 1-2-3 months | Measles, PPV | 1.26 (1.04-1.53) |
| **27** | *pcv13 vs pcv10* | Africa | 6 weeks | NA | 0.61 (0.43-0.87) |
| **27** | *pcv13 vs pcv10* | Africa | 14 weeks | NA | 1.32 (0.95-1.84) |
| **27** | *pcv13 vs pcv10* | Africa | 6-14 weeks | NA | 1.23 (0.90-1.66) |

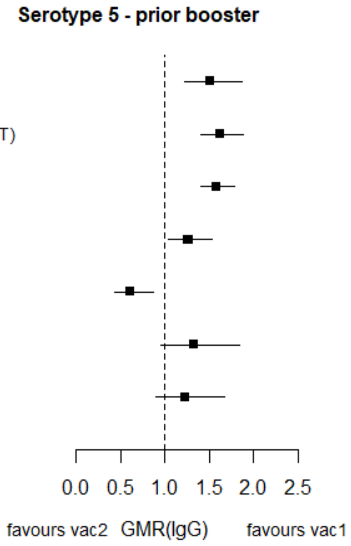

GMR: Geometric mean ratio; pcv: Pneumococcal conjugate vaccine; DTaP – diphtheria and tetanus toxoids, and acellular pertussis vaccine; DTwP – diphtheria and tetanus toxoids, and whole-cell pertussis vaccine; Hib-TT – Haemophilus influenzae type b vaccine (tetanus toxoid conjugate); HB – Hepatitis B vaccine; IPV – Inactivated polio vaccine; OPV – Oral polio vaccine; MenC – Meningococcal C vaccine; TT – tetanus toxoid conjugate; NA: not applicable; enr 3-6 m, 4-8 w int: enrolment at 3-6 months of age, and at 4-8 weeks interval of a 3 doses primary vaccines in total.

Each solid line in the figure shows the GMR from each trial. Black boxes and lines show the point estimates and confidence intervals for geometric mean ratios comparing vac1 vs vac2. Concomitant vaccines are vaccines co-administered with PCV primary vaccine series. Information on co-administered vaccine is not always available. Concomitant vaccines in the bracket are those administered in some but not all of the study sites.

**Supplementary Figure 26. Trial level geometric mean ratios for serotypes 7F at pre-booster.**

| ID | Comparison vac1 vs vac2 | Continent | Schedule-prime | Concomitant vaccines | GMR (95%CI) |
|---|---|---|---|---|---|
| 4 | *pcv13 vs pcv10* | Europe | 2-3-4 months | DTPa-HBV-Hib-TT/IPV | 1.39 (1.15-1.68) |
| 5 | *pcv13 vs pcv10* | Europe | 2–3–4 months | DTPa-HBV-Hib-TT/IPV + (MenC-TT) | 1.30 (1.14-1.50) |
| 6 | *pcv13 vs pcv10* | Asia | 2-4 months | DTPa-HBV-Hib-TT/IPV | 1.29 (1.13-1.47) |
| 18 | *pcv13 vs pcv10* | Oceania | 1-2-3 months | Measles, PPV | 1.35 (1.10-1.65) |
| 27 | *pcv13 vs pcv10* | Africa | 6 weeks | NA | 0.57 (0.40-0.80) |
| 27 | *pcv13 vs pcv10* | Africa | 14 weeks | NA | 1.10 (0.79-1.53) |
| 27 | *pcv13 vs pcv10* | Africa | 6-14 weeks | NA | 1.18 (0.85-1.65) |

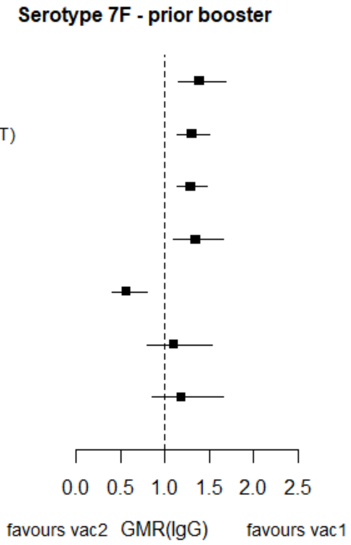

GMR: Geometric mean ratio; pcv: Pneumococcal conjugate vaccine; DTaP – diphtheria and tetanus toxoids, and acellular pertussis vaccine; DTwP – diphtheria and tetanus toxoids, and whole-cell pertussis vaccine; Hib-TT – Haemophilus influenzae type b vaccine (tetanus toxoid conjugate); HB – Hepatitis B vaccine; IPV – Inactivated polio vaccine; OPV – Oral polio vaccine; MenC – Meningococcal C vaccine; TT – tetanus toxoid conjugate; NA: not applicable; enr 3-6 m, 4-8 w int: enrolment at 3-6 months of age, and at 4-8 weeks interval of a 3 doses primary vaccines in total.

Each solid line in the figure shows the GMR from each trial. Black boxes and lines show the point estimates and confidence intervals for geometric mean ratios comparing vac1 vs vac2. Concomitant vaccines are vaccines co-administered with PCV primary vaccine series. Information on co-administered vaccine is not always available. Concomitant vaccines in the bracket are those administered in some but not all of the study sites.

**Supplementary Figure 27. Trial level geometric mean ratios for serotypes 3 at pre-booster.**

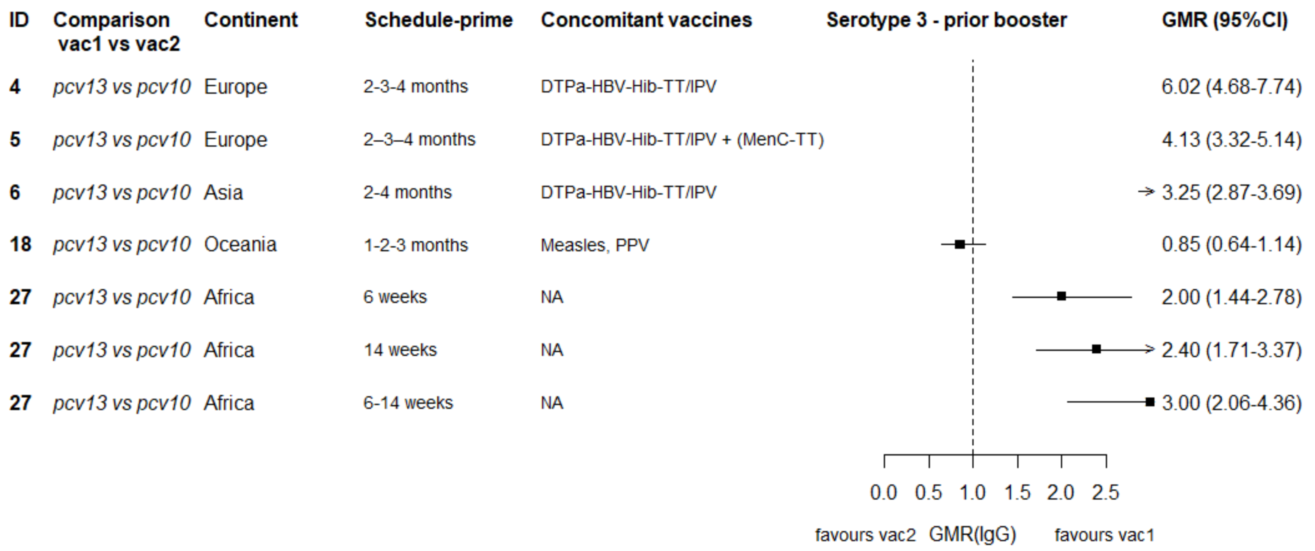

GMR: Geometric mean ratio; pcv: Pneumococcal conjugate vaccine; DTaP – diphtheria and tetanus toxoids, and acellular pertussis vaccine; DTwP – diphtheria and tetanus toxoids, and whole-cell pertussis vaccine; Hib-TT – Haemophilus influenzae type b vaccine (tetanus toxoid conjugate); HB – Hepatitis B vaccine; IPV – Inactivated polio vaccine; OPV – Oral polio vaccine; MenC – Meningococcal C vaccine; TT – tetanus toxoid conjugate; NA: not applicable; enr 3-6 m, 4-8 w int: enrolment at 3-6 months of age, and at 4-8 weeks interval of a 3 doses primary vaccines in total.

Each solid line in the figure shows the GMR from each trial. Black boxes and lines show the point estimates and confidence intervals for geometric mean ratios comparing vac1 vs vac2. Concomitant vaccines are vaccines co-administered with PCV primary vaccine series. Information on co-administered vaccine is not always available. Concomitant vaccines in the bracket are those administered in some but not all of the study sites.

**Supplementary Figure 28. Trial level geometric mean ratios for serotypes 6A at pre-booster.**

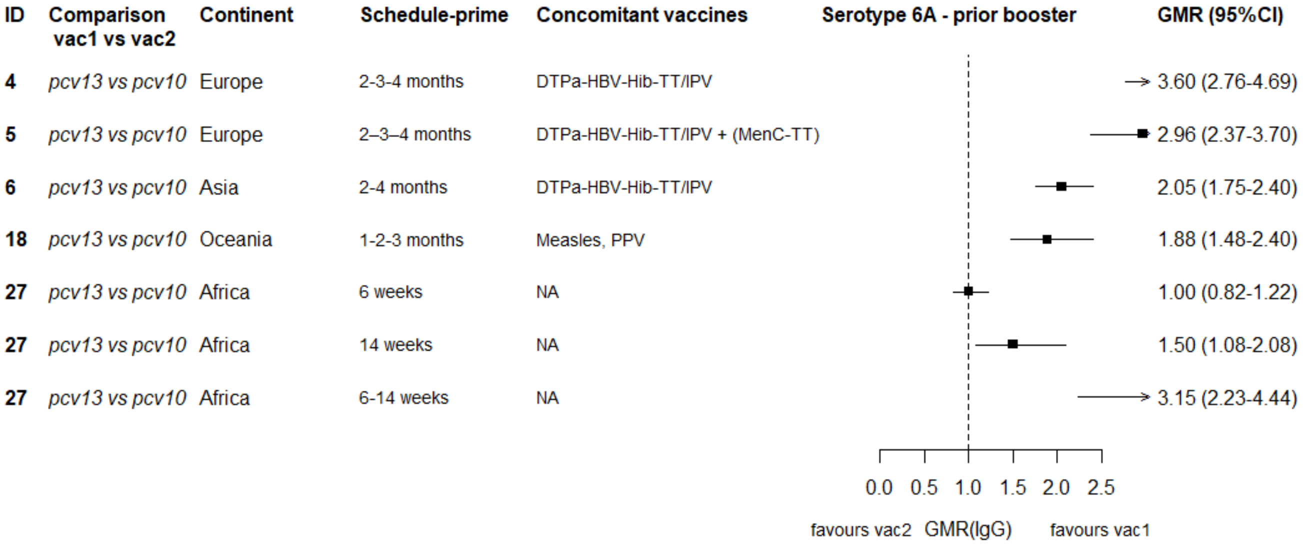

GMR: Geometric mean ratio; pcv: Pneumococcal conjugate vaccine; DTaP – diphtheria and tetanus toxoids, and acellular pertussis vaccine; DTwP – diphtheria and tetanus toxoids, and whole-cell pertussis vaccine; Hib-TT – Haemophilus influenzae type b vaccine (tetanus toxoid conjugate); HB – Hepatitis B vaccine; IPV – Inactivated polio vaccine; OPV – Oral polio vaccine; MenC – Meningococcal C vaccine; TT – tetanus toxoid conjugate; NA: not applicable; enr 3-6 m, 4-8 w int: enrolment at 3-6 months of age, and at 4-8 weeks interval of a 3 doses primary vaccines in total.

Each solid line in the figure shows the GMR from each trial. Black boxes and lines show the point estimates and confidence intervals for geometric mean ratios comparing vac1 vs vac2. Concomitant vaccines are vaccines co-administered with PCV primary vaccine series. Information on co-administered vaccine is not always available. Concomitant vaccines in the bracket are those administered in some but not all of the study sites.

**Supplementary Figure 29. Trial level geometric mean ratios for serotypes 19A at pre-booster.**

| ID | Comparison vac1 vs vac2 | Continent | Schedule-prime | Concomitant vaccines | Serotype 19A - prior booster | GMR (95%CI) |
|---|---|---|---|---|---|---|
| 4 | *pcv13 vs pcv10* | Europe | 2-3-4 months | DTPa-HBV-Hib-TT/IPV | | 3.21 (2.34-4.40) |
| 5 | *pcv13 vs pcv10* | Europe | 2–3–4 months | DTPa-HBV-Hib-TT/IPV + (MenC-TT) | | 2.25 (1.79-2.83) |
| 6 | *pcv13 vs pcv10* | Asia | 2-4 months | DTPa-HBV-Hib-TT/IPV | | 1.89 (1.63-2.19) |
| 18 | *pcv13 vs pcv10* | Oceania | 1-2-3 months | Measles, PPV | | 1.43 (1.11-1.84) |
| 27 | *pcv13 vs pcv10* | Africa | 6 weeks | NA | | 2.08 (0.48-8.91) |
| 27 | *pcv13 vs pcv10* | Africa | 14 weeks | NA | | 2.00 (1.45-2.77) |
| 27 | *pcv13 vs pcv10* | Africa | 6-14 weeks | NA | | 3.46 (2.56-4.68) |

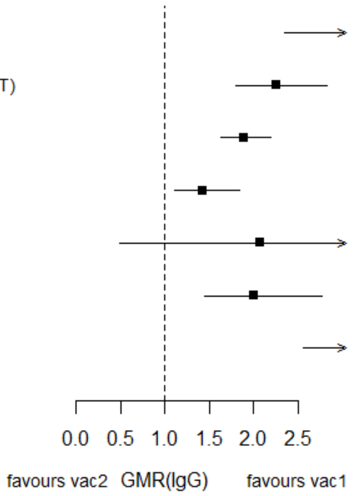

GMR: Geometric mean ratio; pcv: Pneumococcal conjugate vaccine; DTaP – diphtheria and tetanus toxoids, and acellular pertussis vaccine; DTwP – diphtheria and tetanus toxoids, and whole-cell pertussis vaccine; Hib-TT – Haemophilus influenzae type b vaccine (tetanus toxoid conjugate); HB – Hepatitis B vaccine; IPV – Inactivated polio vaccine; OPV – Oral polio vaccine; MenC – Meningococcal C vaccine; TT – tetanus toxoid conjugate; NA: not applicable; enr 3-6 m, 4-8 w int: enrolment at 3-6 months of age, and at 4-8 weeks interval of a 3 doses primary vaccines in total.

Each solid line in the figure shows the GMR from each trial. Black boxes and lines show the point estimates and confidence intervals for geometric mean ratios comparing vac1 vs vac2. Concomitant vaccines are vaccines co-administered with PCV primary vaccine series. Information on co-administered vaccine is not always available. Concomitant vaccines in the bracket are those administered in some but not all of the study sites.

**Supplementary Figure 30. Trial level geometric mean ratios for serotypes 4 at post-booster.**

| ID | Comparison vac1 vs vac2 | Continent | Schedule-prime | Concomitant vaccines | GMR (95%CI) |
|---|---|---|---|---|---|
| 1 | *pcv7 vs pcv10* | Asia | 6-10-24 weeks | DTPw-HBV-Hib-TT + OPV | 0.89 (0.65-1.21) |
| 1 | *pcv7 vs pcv10* | Europe | 2-4-6 months | DTPw-HBV-Hib-TT + IPV | 1.58 (1.21-2.06) |
| 2 | *pcv7 vs pcv10* | Asia | 2-4-6 months | Hib-TT | 2.11 (1.73-2.57) |
| 7 | *pcv7 vs pcv10* | Europe | 2-3-4 months | DTPa-(HBV)-Hib-TT/IPV | 1.25 (1.08-1.46) |
| 8 | *pcv7 vs pcv10* | Europe | 2-3-4 months | DTPa-(HBV)-Hib-TT/IPV | 1.60 (1.41-1.81) |
| 9 | *pcv7 vs pcv10* | Europe | 2-4-6 months | DTPa-(HBV)-Hib-IPV + MenC | 1.39 (1.17-1.65) |
| 4 | *pcv13 vs pcv10* | Europe | 2-3-4 months | DTPa-HBV-Hib-TT/IPV | 1.09 (0.91-1.32) |
| 5 | *pcv13 vs pcv10* | Europe | 2–3–4 months | DTPa-HBV-Hib-TT/IPV + (MenC-TT) | 0.97 (0.84-1.11) |
| 6 | *pcv13 vs pcv10* | Asia | 2-4 months | DTPa-HBV-Hib-TT/IPV | 1.12 (0.96-1.31) |
| 27 | *pcv13 vs pcv10* | Africa | 6 weeks | NA | 1.36 (1.02-1.83) |
| 27 | *pcv13 vs pcv10* | Africa | 14 weeks | NA | 0.84 (0.64-1.11) |
| 27 | *pcv13 vs pcv10* | Africa | 6-14 weeks | NA | 1.00 (0.75-1.33) |
| 10 | *pcv13 vs pcv7* | Asia | 6-10-14 weeks | NA | 0.90 (0.75-1.08) |
| 11 | *pcv13 vs pcv7* | Asia | 2-4-6 months | NA | 0.99 (0.90-1.08) |
| 12 | *pcv13 vs pcv7* | Europe | 3–5 months | DTPa-HBV-Hib-TT/IPV | 0.67 (0.58-0.78) |
| 13 | *pcv13 vs pcv7* | Europe | 2-3-4 months | DTPa-Hib-TT/IPV | 0.83 (0.70-0.98) |
| 14 | *pcv13 vs pcv7* | Asia | 2-4-6 months | NA | 0.64 (0.49-0.83) |
| 15 | *pcv13 vs pcv7* | Europe | 2-3-4 months | DTPa-HBV-Hib-TT/IPV | 0.83 (0.71-0.97) |
| 16 | *pcv13 vs pcv7* | Asia | 2-4-6 months | NA | 0.82 (0.63-1.07) |
| 17 | *pcv13 vs pcv7* | North America | 2-4-6 months | MMR | 0.92 (0.80-1.05) |
| 19 | *pcv13 vs pcv7* | Europe | 2-4 months | Hib-MenC-TT | 0.61 (0.41-0.91) |
| 20 | *pcv13 vs pcv7* | Asia | enr 3-6 m, 4-8 w int | DTPa | 0.92 (0.76-1.13) |
| 21 | *pcv13 vs pcv7* | South America | 2-4-6 months | DTwP-Hib, OPV | 0.91 (0.75-1.10) |
| 22 | *pcv13 vs pcv7* | North America | 2-4-6 months | MMRV, Hib, HAV | 0.69 (0.59-0.81) |
| 23 | *pcv13 vs pcv7* | Asia | 3-4-5 months | NA | 0.85 (0.75-0.96) |

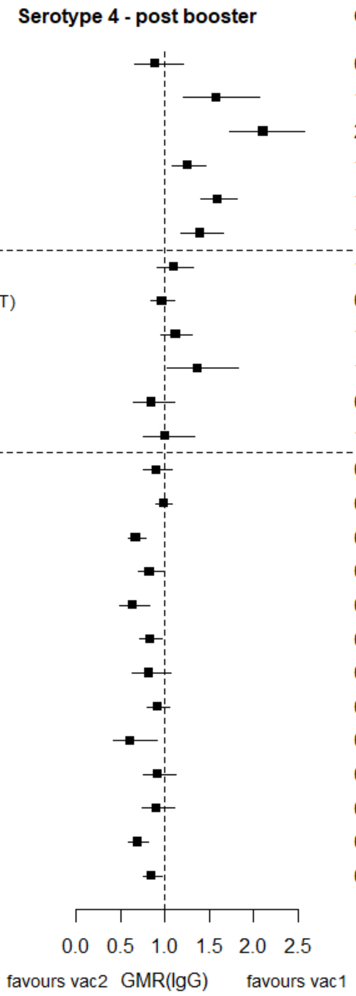

GMR: Geometric mean ratio; pcv: Pneumococcal conjugate vaccine; DTaP – diphtheria and tetanus toxoids, and acellular pertussis vaccine; DTwP – diphtheria and tetanus toxoids, and whole-cell pertussis vaccine; Hib-TT – Haemophilus influenzae type b vaccine (tetanus toxoid conjugate); HB – Hepatitis B vaccine; IPV – Inactivated polio vaccine; OPV – Oral polio vaccine; MenC – Meningococcal C vaccine; TT – tetanus toxoid conjugate; NA: not applicable; enr 3-6 m, 4-8 w int: enrolment at 3-6 months of age, and at 4-8 weeks interval of a 3 doses primary vaccines in total.

Each solid line in the figure shows the GMR from each trial. Black boxes and lines show the point estimates and confidence intervals for geometric mean ratios comparing vac1 vs vac2. Concomitant vaccines are vaccines co-administered with PCV primary vaccine series. Information on co-administered vaccine is not always available. Concomitant vaccines in the bracket are those administered in some but not all of the study sites.

**Supplementary Figure 31. Trial level geometric mean ratios for serotypes 6B at post-booster.**

| ID | Comparison vac1 vs vac2 | Continent | Schedule-prime | Concomitant vaccines | GMR (95%CI) |
|---|---|---|---|---|---|
| 1 | *pcv7 vs pcv10* | Asia | 6-10-24 weeks | DTPw-HBV-Hib-TT + OPV | 1.31 (0.92-1.87) |
| 1 | *pcv7 vs pcv10* | Europe | 2-4-6 months | DTPw-HBV-Hib-TT + IPV | 2.68 (1.79-4.00) |
| 2 | *pcv7 vs pcv10* | Asia | 2-4-6 months | Hib-TT | 2.45 (1.97-3.06) |
| 7 | *pcv7 vs pcv10* | Europe | 2-3-4 months | DTPa-(HBV)-Hib-TT/IPV | 1.72 (1.41-2.08) |
| 8 | *pcv7 vs pcv10* | Europe | 2-3-4 months | DTPa-(HBV)-Hib-TT/IPV | 1.32 (1.12-1.56) |
| 9 | *pcv7 vs pcv10* | Europe | 2-4-6 months | DTPa-(HBV)-Hib-IPV + MenC | 2.26 (1.89-2.71) |
| 4 | *pcv13 vs pcv10* | Europe | 2-3-4 months | DTPa-HBV-Hib-TT/IPV | 1.38 (1.11-1.72) |
| 5 | *pcv13 vs pcv10* | Europe | 2–3–4 months | DTPa-HBV-Hib-TT/IPV + (MenC-TT) | 1.52 (1.28-1.80) |
| 6 | *pcv13 vs pcv10* | Asia | 2-4 months | DTPa-HBV-Hib-TT/IPV | 1.54 (1.27-1.87) |
| 27 | *pcv13 vs pcv10* | Africa | 6 weeks | NA | 0.48 (0.34-0.68) |
| 27 | *pcv13 vs pcv10* | Africa | 14 weeks | NA | 0.85 (0.59-1.22) |
| 27 | *pcv13 vs pcv10* | Africa | 6-14 weeks | NA | 1.60 (1.18-2.15) |
| 10 | *pcv13 vs pcv7* | Asia | 6-10-14 weeks | NA | 1.06 (0.86-1.30) |
| 11 | *pcv13 vs pcv7* | Asia | 2-4-6 months | NA | 0.91 (0.82-1.00) |
| 12 | *pcv13 vs pcv7* | Europe | 3–5 months | DTPa-HBV-Hib-TT/IPV | 0.95 (0.79-1.14) |
| 13 | *pcv13 vs pcv7* | Europe | 2-3-4 months | DTPa-Hib-TT/IPV | 0.91 (0.76-1.10) |
| 14 | *pcv13 vs pcv7* | Asia | 2-4-6 months | NA | 1.03 (0.78-1.36) |
| 15 | *pcv13 vs pcv7* | Europe | 2-3-4 months | DTPa-HBV-Hib-TT/IPV | 0.94 (0.79-1.11) |
| 16 | *pcv13 vs pcv7* | Asia | 2-4-6 months | NA | 1.12 (0.85-1.47) |
| 17 | *pcv13 vs pcv7* | North America | 2-4-6 months | MMR | 1.07 (0.93-1.22) |
| 19 | *pcv13 vs pcv7* | Europe | 2-4 months | Hib-MenC-TT | 1.10 (0.64-1.90) |
| 20 | *pcv13 vs pcv7* | Asia | enr 3-6 m, 4-8 w int | DTPa | 1.21 (0.97-1.50) |
| 21 | *pcv13 vs pcv7* | South America | 2-4-6 months | DTwP-Hib, OPV | 1.13 (0.89-1.43) |
| 22 | *pcv13 vs pcv7* | North America | 2-4-6 months | MMRV, Hib, HAV | 0.73 (0.61-0.88) |
| 23 | *pcv13 vs pcv7* | Asia | 3-4-5 months | NA | 0.94 (0.81-1.08) |

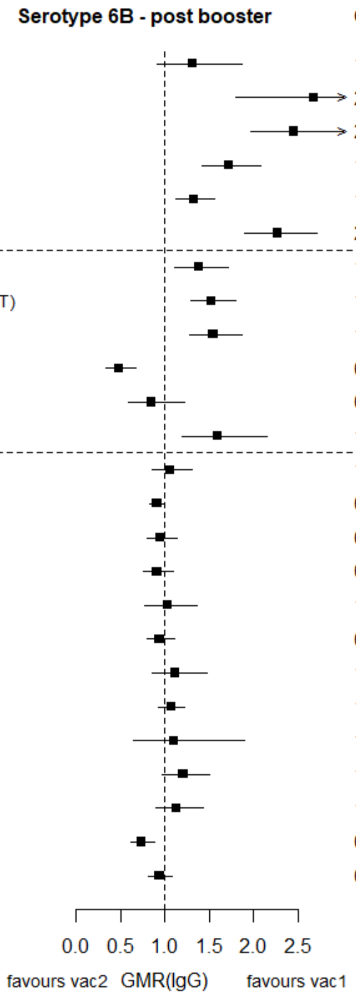

GMR: Geometric mean ratio; pcv: Pneumococcal conjugate vaccine; DTaP – diphtheria and tetanus toxoids, and acellular pertussis vaccine; DTwP – diphtheria and tetanus toxoids, and whole-cell pertussis vaccine; Hib-TT – Haemophilus influenzae type b vaccine (tetanus toxoid conjugate); HB – Hepatitis B vaccine; IPV – Inactivated polio vaccine; OPV – Oral polio vaccine; MenC – Meningococcal C vaccine; TT – tetanus toxoid conjugate; NA: not applicable; enr 3-6 m, 4-8 w int: enrolment at 3-6 months of age, and at 4-8 weeks interval of a 3 doses primary vaccines in total.

Each solid line in the figure shows the GMR from each trial. Black boxes and lines show the point estimates and confidence intervals for geometric mean ratios comparing vac1 vs vac2. Concomitant vaccines are vaccines co-administered with PCV primary vaccine series. Information on co-administered vaccine is not always available. Concomitant vaccines in the bracket are those administered in some but not all of the study sites.

**Supplementary Figure 32. Trial level geometric mean ratios for serotypes 9V at post-booster.**

| ID | Comparison vac1 vs vac2 | Continent | Schedule-prime | Concomitant vaccines | GMR (95%CI) |
|---|---|---|---|---|---|
| 1 | *pcv7 vs pcv10* | Asia | 6-10-24 weeks | DTPw-HBV-Hib-TT + OPV | 1.37 (1.03-1.83) |
| 1 | *pcv7 vs pcv10* | Europe | 2-4-6 months | DTPw-HBV-Hib-TT + IPV | 2.86 (2.26-3.62) |
| 2 | *pcv7 vs pcv10* | Asia | 2-4-6 months | Hib-TT | 2.53 (2.12-3.01) |
| 7 | *pcv7 vs pcv10* | Europe | 2-3-4 months | DTPa-(HBV)-Hib-TT/IPV | 1.47 (1.28-1.68) |
| 8 | *pcv7 vs pcv10* | Europe | 2-3-4 months | DTPa-(HBV)-Hib-TT/IPV | 1.05 (0.92-1.20) |
| 9 | *pcv7 vs pcv10* | Europe | 2-4-6 months | DTPa-(HBV)-Hib-IPV + MenC | 1.91 (1.63-2.24) |
| 4 | *pcv13 vs pcv10* | Europe | 2-3-4 months | DTPa-HBV-Hib-TT/IPV | 1.26 (1.04-1.52) |
| 5 | *pcv13 vs pcv10* | Europe | 2–3–4 months | DTPa-HBV-Hib-TT/IPV + (MenC-TT) | 1.57 (1.37-1.80) |
| 6 | *pcv13 vs pcv10* | Asia | 2-4 months | DTPa-HBV-Hib-TT/IPV | 1.57 (1.36-1.80) |
| 27 | *pcv13 vs pcv10* | Africa | 6 weeks | NA | 1.47 (1.11-1.95) |
| 27 | *pcv13 vs pcv10* | Africa | 14 weeks | NA | 1.53 (1.21-1.94) |
| 27 | *pcv13 vs pcv10* | Africa | 6-14 weeks | NA | 1.33 (1.00-1.78) |
| 10 | *pcv13 vs pcv7* | Asia | 6-10-14 weeks | NA | 0.91 (0.77-1.07) |
| 11 | *pcv13 vs pcv7* | Asia | 2-4-6 months | NA | 0.94 (0.87-1.02) |
| 12 | *pcv13 vs pcv7* | Europe | 3–5 months | DTPa-HBV-Hib-TT/IPV | 0.73 (0.64-0.84) |
| 13 | *pcv13 vs pcv7* | Europe | 2-3-4 months | DTPa-Hib-TT/IPV | 0.79 (0.68-0.92) |
| 14 | *pcv13 vs pcv7* | Asia | 2-4-6 months | NA | 0.83 (0.65-1.06) |
| 15 | *pcv13 vs pcv7* | Europe | 2-3-4 months | DTPa-HBV-Hib-TT/IPV | 0.83 (0.73-0.94) |
| 16 | *pcv13 vs pcv7* | Asia | 2-4-6 months | NA | 0.99 (0.79-1.23) |
| 17 | *pcv13 vs pcv7* | North America | 2-4-6 months | MMR | 0.99 (0.88-1.11) |
| 19 | *pcv13 vs pcv7* | Europe | 2-4 months | Hib-MenC-TT | 0.78 (0.52-1.16) |
| 20 | *pcv13 vs pcv7* | Asia | enr 3-6 m, 4-8 w int | DTPa | 0.81 (0.67-0.97) |
| 21 | *pcv13 vs pcv7* | South America | 2-4-6 months | DTwP-Hib, OPV | 1.12 (0.94-1.34) |
| 22 | *pcv13 vs pcv7* | North America | 2-4-6 months | MMRV, Hib, HAV | 0.72 (0.62-0.84) |
| 23 | *pcv13 vs pcv7* | Asia | 3-4-5 months | NA | 0.80 (0.71-0.90) |

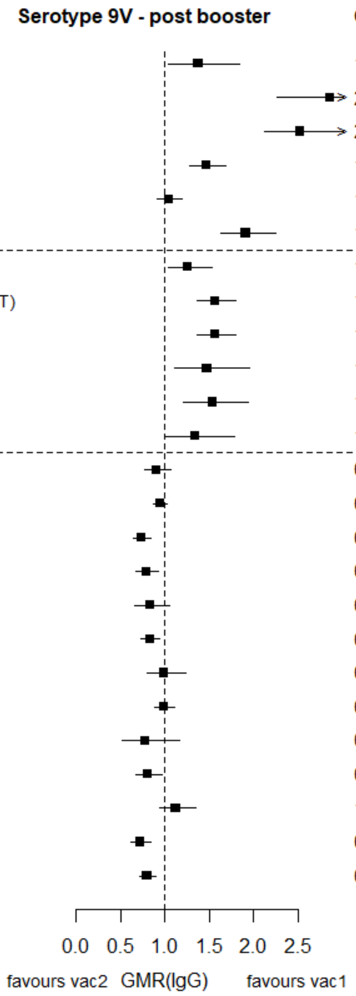

GMR: Geometric mean ratio; pcv: Pneumococcal conjugate vaccine; DTaP – diphtheria and tetanus toxoids, and acellular pertussis vaccine; DTwP – diphtheria and tetanus toxoids, and whole-cell pertussis vaccine; Hib-TT – Haemophilus influenzae type b vaccine (tetanus toxoid conjugate); HB – Hepatitis B vaccine; IPV – Inactivated polio vaccine; OPV – Oral polio vaccine; MenC – Meningococcal C vaccine; TT – tetanus toxoid conjugate; NA: not applicable; enr 3-6 m, 4-8 w int: enrolment at 3-6 months of age, and at 4-8 weeks interval of a 3 doses primary vaccines in total.

Each solid line in the figure shows the GMR from each trial. Black boxes and lines show the point estimates and confidence intervals for geometric mean ratios comparing vac1 vs vac2. Concomitant vaccines are vaccines co-administered with PCV primary vaccine series. Information on co-administered vaccine is not always available. Concomitant vaccines in the bracket are those administered in some but not all of the study sites.

**Supplementary Figure 33. Trial level geometric mean ratios for serotypes 14 at post-booster.**

| ID | Comparison vac1 vs vac2 | Continent | Schedule-prime | Concomitant vaccines | GMR (95%CI) |
|---|---|---|---|---|---|
| 1 | *pcv7 vs pcv10* | Asia | 6-10-24 weeks | DTPw-HBV-Hib-TT + OPV | 1.61 (1.18-2.20) |
| 1 | *pcv7 vs pcv10* | Europe | 2-4-6 months | DTPw-HBV-Hib-TT + IPV | 2.35 (1.74-3.18) |
| 2 | *pcv7 vs pcv10* | Asia | 2-4-6 months | Hib-TT | 2.71 (2.26-3.24) |
| 7 | *pcv7 vs pcv10* | Europe | 2-3-4 months | DTPa-(HBV)-Hib-TT/IPV | 1.56 (1.33-1.83) |
| 8 | *pcv7 vs pcv10* | Europe | 2-3-4 months | DTPa-(HBV)-Hib-TT/IPV | 1.24 (1.08-1.43) |
| 9 | *pcv7 vs pcv10* | Europe | 2-4-6 months | DTPa-(HBV)-Hib-IPV + MenC | 1.94 (1.62-2.32) |
| 4 | *pcv13 vs pcv10* | Europe | 2-3-4 months | DTPa-HBV-Hib-TT/IPV | 1.73 (1.39-2.16) |
| 5 | *pcv13 vs pcv10* | Europe | 2–3–4 months | DTPa-HBV-Hib-TT/IPV + (MenC-TT) | 1.72 (1.47-2.02) |
| 6 | *pcv13 vs pcv10* | Asia | 2-4 months | DTPa-HBV-Hib-TT/IPV | 1.31 (1.11-1.54) |
| 27 | *pcv13 vs pcv10* | Africa | 6 weeks | NA | 1.21 (0.76-1.93) |
| 27 | *pcv13 vs pcv10* | Africa | 14 weeks | NA | 1.11 (0.79-1.56) |
| 27 | *pcv13 vs pcv10* | Africa | 6-14 weeks | NA | 1.76 (1.18-2.62) |
| 10 | *pcv13 vs pcv7* | Asia | 6-10-14 weeks | NA | 0.97 (0.79-1.19) |
| 11 | *pcv13 vs pcv7* | Asia | 2-4-6 months | NA | 0.89 (0.81-0.98) |
| 12 | *pcv13 vs pcv7* | Europe | 3–5 months | DTPa-HBV-Hib-TT/IPV | 0.86 (0.74-1.00) |
| 13 | *pcv13 vs pcv7* | Europe | 2-3-4 months | DTPa-Hib-TT/IPV | 0.89 (0.75-1.06) |
| 14 | *pcv13 vs pcv7* | Asia | 2-4-6 months | NA | 0.69 (0.51-0.92) |
| 15 | *pcv13 vs pcv7* | Europe | 2-3-4 months | DTPa-HBV-Hib-TT/IPV | 0.77 (0.66-0.90) |
| 16 | *pcv13 vs pcv7* | Asia | 2-4-6 months | NA | 0.72 (0.57-0.92) |
| 17 | *pcv13 vs pcv7* | North America | 2-4-6 months | MMR | 0.85 (0.75-0.95) |
| 19 | *pcv13 vs pcv7* | Europe | 2-4 months | Hib-MenC-TT | 0.89 (0.61-1.30) |
| 20 | *pcv13 vs pcv7* | Asia | enr 3-6 m, 4-8 w int | DTPa | 0.95 (0.80-1.12) |
| 21 | *pcv13 vs pcv7* | South America | 2-4-6 months | DTwP-Hib, OPV | 1.06 (0.86-1.31) |
| 22 | *pcv13 vs pcv7* | North America | 2-4-6 months | MMRV, Hib, HAV | 0.74 (0.62-0.88) |
| 23 | *pcv13 vs pcv7* | Asia | 3-4-5 months | NA | 0.88 (0.79-0.98) |

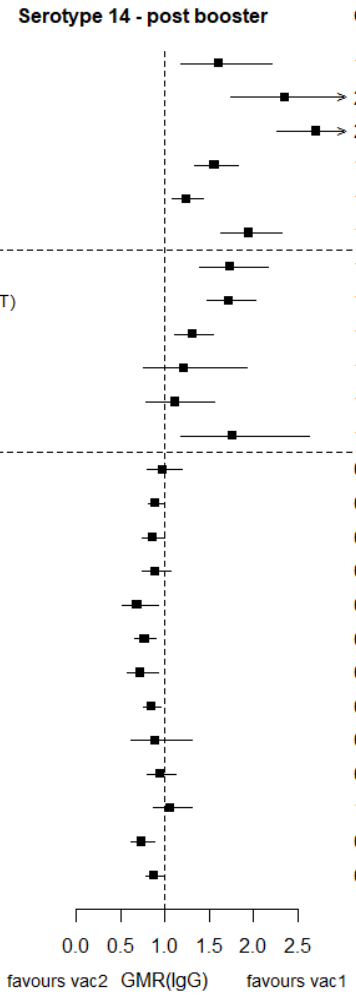

GMR: Geometric mean ratio; pcv: Pneumococcal conjugate vaccine; DTaP – diphtheria and tetanus toxoids, and acellular pertussis vaccine; DTwP – diphtheria and tetanus toxoids, and whole-cell pertussis vaccine; Hib-TT – Haemophilus influenzae type b vaccine (tetanus toxoid conjugate); HB – Hepatitis B vaccine; IPV – Inactivated polio vaccine; OPV – Oral polio vaccine; MenC – Meningococcal C vaccine; TT – tetanus toxoid conjugate; NA: not applicable; enr 3-6 m, 4-8 w int: enrolment at 3-6 months of age, and at 4-8 weeks interval of a 3 doses primary vaccines in total.

Each solid line in the figure shows the GMR from each trial. Black boxes and lines show the point estimates and confidence intervals for geometric mean ratios comparing vac1 vs vac2. Concomitant vaccines are vaccines co-administered with PCV primary vaccine series. Information on co-administered vaccine is not always available. Concomitant vaccines in the bracket are those administered in some but not all of the study sites.

**Supplementary Figure 34. Trial level geometric mean ratios for serotypes 18C at post-booster.**

| ID | Comparison vac1 vs vac2 | Continent | Schedule-prime | Concomitant vaccines | GMR (95%CI) |
|---|---|---|---|---|---|
| 1 | *pcv7 vs pcv10* | Asia | 6-10-24 weeks | DTPw-HBV-Hib-TT + OPV | 0.31 (0.23-0.41) |
| 1 | *pcv7 vs pcv10* | Europe | 2-4-6 months | DTPw-HBV-Hib-TT + IPV | 0.90 (0.67-1.19) |
| 2 | *pcv7 vs pcv10* | Asia | 2-4-6 months | Hib-TT | 1.09 (0.89-1.33) |
| 7 | *pcv7 vs pcv10* | Europe | 2-3-4 months | DTPa-(HBV)-Hib-TT/IPV | 0.95 (0.81-1.10) |
| 8 | *pcv7 vs pcv10* | Europe | 2-3-4 months | DTPa-(HBV)-Hib-TT/IPV | 1.11 (0.97-1.27) |
| 9 | *pcv7 vs pcv10* | Europe | 2-4-6 months | DTPa-(HBV)-Hib-IPV + MenC | 0.95 (0.81-1.11) |
| 4 | *pcv13 vs pcv10* | Europe | 2-3-4 months | DTPa-HBV-Hib-TT/IPV | 0.85 (0.71-1.03) |
| 5 | *pcv13 vs pcv10* | Europe | 2–3–4 months | DTPa-HBV-Hib-TT/IPV + (MenC-TT) | 0.79 (0.68-0.92) |
| 6 | *pcv13 vs pcv10* | Asia | 2-4 months | DTPa-HBV-Hib-TT/IPV | 0.84 (0.72-0.96) |
| 27 | *pcv13 vs pcv10* | Africa | 6 weeks | NA | 1.10 (0.85-1.43) |
| 27 | *pcv13 vs pcv10* | Africa | 14 weeks | NA | 0.71 (0.54-0.93) |
| 27 | *pcv13 vs pcv10* | Africa | 6-14 weeks | NA | 0.90 (0.70-1.14) |
| 10 | *pcv13 vs pcv7* | Asia | 6-10-14 weeks | NA | 0.88 (0.74-1.03) |
| 11 | *pcv13 vs pcv7* | Asia | 2-4-6 months | NA | 0.99 (0.90-1.09) |
| 12 | *pcv13 vs pcv7* | Europe | 3–5 months | DTPa-HBV-Hib-TT/IPV | 0.67 (0.58-0.77) |
| 13 | *pcv13 vs pcv7* | Europe | 2-3-4 months | DTPa-Hib-TT/IPV | 0.81 (0.68-0.95) |
| 14 | *pcv13 vs pcv7* | Asia | 2-4-6 months | NA | 0.76 (0.58-1.00) |
| 15 | *pcv13 vs pcv7* | Europe | 2-3-4 months | DTPa-HBV-Hib-TT/IPV | 0.83 (0.72-0.96) |
| 16 | *pcv13 vs pcv7* | Asia | 2-4-6 months | NA | 0.94 (0.74-1.20) |
| 17 | *pcv13 vs pcv7* | North America | 2-4-6 months | MMR | 0.94 (0.82-1.07) |
| 19 | *pcv13 vs pcv7* | Europe | 2-4 months | Hib-MenC-TT | 0.71 (0.52-0.98) |
| 20 | *pcv13 vs pcv7* | Asia | enr 3-6 m, 4-8 w int | DTPa | 0.81 (0.67-0.99) |
| 21 | *pcv13 vs pcv7* | South America | 2-4-6 months | DTwP-Hib, OPV | 0.87 (0.71-1.07) |
| 22 | *pcv13 vs pcv7* | North America | 2-4-6 months | MMRV, Hib, HAV | 0.68 (0.58-0.80) |
| 23 | *pcv13 vs pcv7* | Asia | 3-4-5 months | NA | 0.99 (0.88-1.11) |

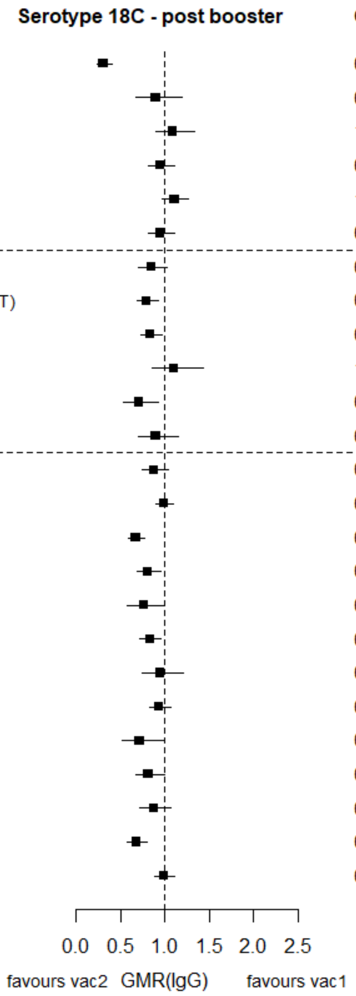

GMR: Geometric mean ratio; pcv: Pneumococcal conjugate vaccine; DTaP – diphtheria and tetanus toxoids, and acellular pertussis vaccine; DTwP – diphtheria and tetanus toxoids, and whole-cell pertussis vaccine; Hib-TT – Haemophilus influenzae type b vaccine (tetanus toxoid conjugate); HB – Hepatitis B vaccine; IPV – Inactivated polio vaccine; OPV – Oral polio vaccine; MenC – Meningococcal C vaccine; TT – tetanus toxoid conjugate; NA: not applicable; enr 3-6 m, 4-8 w int: enrolment at 3-6 months of age, and at 4-8 weeks interval of a 3 doses primary vaccines in total.

Each solid line in the figure shows the GMR from each trial. Black boxes and lines show the point estimates and confidence intervals for geometric mean ratios comparing vac1 vs vac2. Concomitant vaccines are vaccines co-administered with PCV primary vaccine series. Information on co-administered vaccine is not always available. Concomitant vaccines in the bracket are those administered in some but not all of the study sites.

**Supplementary Figure 35. Trial level geometric mean ratios for serotypes 19F at post-booster.**

| ID | Comparison vac1 vs vac2 | Continent | Schedule-prime | Concomitant vaccines | GMR (95%CI) |
|---|---|---|---|---|---|
| 1 | *pcv7 vs pcv10* | Asia | 6-10-24 weeks | DTPw-HBV-Hib-TT + OPV | 0.31 (0.23-0.43) |
| 1 | *pcv7 vs pcv10* | Europe | 2-4-6 months | DTPw-HBV-Hib-TT + IPV | 0.46 (0.34-0.62) |
| 2 | *pcv7 vs pcv10* | Asia | 2-4-6 months | Hib-TT | 0.29 (0.23-0.36) |
| 7 | *pcv7 vs pcv10* | Europe | 2-3-4 months | DTPa-(HBV)-Hib-TT/IPV | 0.44 (0.37-0.51) |
| 8 | *pcv7 vs pcv10* | Europe | 2-3-4 months | DTPa-(HBV)-Hib-TT/IPV | 0.84 (0.72-0.98) |
| 9 | *pcv7 vs pcv10* | Europe | 2-4-6 months | DTPa-(HBV)-Hib-IPV + MenC | 0.51 (0.44-0.60) |
| 4 | *pcv13 vs pcv10* | Europe | 2-3-4 months | DTPa-HBV-Hib-TT/IPV | 0.97 (0.79-1.19) |
| 5 | *pcv13 vs pcv10* | Europe | 2–3–4 months | DTPa-HBV-Hib-TT/IPV + (MenC-TT) | 0.77 (0.67-0.89) |
| 6 | *pcv13 vs pcv10* | Asia | 2-4 months | DTPa-HBV-Hib-TT/IPV | 0.72 (0.62-0.84) |
| 27 | *pcv13 vs pcv10* | Africa | 6 weeks | NA | 1.36 (0.90-2.06) |
| 27 | *pcv13 vs pcv10* | Africa | 14 weeks | NA | 2.02 (1.32-3.11) |
| 27 | *pcv13 vs pcv10* | Africa | 6-14 weeks | NA | 1.16 (0.88-1.54) |
| 10 | *pcv13 vs pcv7* | Asia | 6-10-14 weeks | NA | 1.49 (1.20-1.86) |
| 11 | *pcv13 vs pcv7* | Asia | 2-4-6 months | NA | 2.07 (1.87-2.29) |
| 12 | *pcv13 vs pcv7* | Europe | 3–5 months | DTPa-HBV-Hib-TT/IPV | 1.10 (0.91-1.32) |
| 13 | *pcv13 vs pcv7* | Europe | 2-3-4 months | DTPa-Hib-TT/IPV | 1.23 (0.98-1.54) |
| 14 | *pcv13 vs pcv7* | Asia | 2-4-6 months | NA | 1.09 (0.83-1.42) |
| 15 | *pcv13 vs pcv7* | Europe | 2-3-4 months | DTPa-HBV-Hib-TT/IPV | 1.26 (1.06-1.50) |
| 16 | *pcv13 vs pcv7* | Asia | 2-4-6 months | NA | 1.42 (1.10-1.84) |
| 17 | *pcv13 vs pcv7* | North America | 2-4-6 months | MMR | 1.60 (1.37-1.86) |
| 19 | *pcv13 vs pcv7* | Europe | 2-4 months | Hib-MenC-TT | 1.56 (0.91-2.65) |
| 20 | *pcv13 vs pcv7* | Asia | enr 3-6 m, 4-8 w int | DTPa | 1.78 (1.42-2.23) |
| 21 | *pcv13 vs pcv7* | South America | 2-4-6 months | DTwP-Hib, OPV | 1.48 (1.19-1.84) |
| 22 | *pcv13 vs pcv7* | North America | 2-4-6 months | MMRV, Hib, HAV | 1.20 (1.01-1.42) |
| 23 | *pcv13 vs pcv7* | Asia | 3-4-5 months | NA | 1.56 (1.38-1.77) |

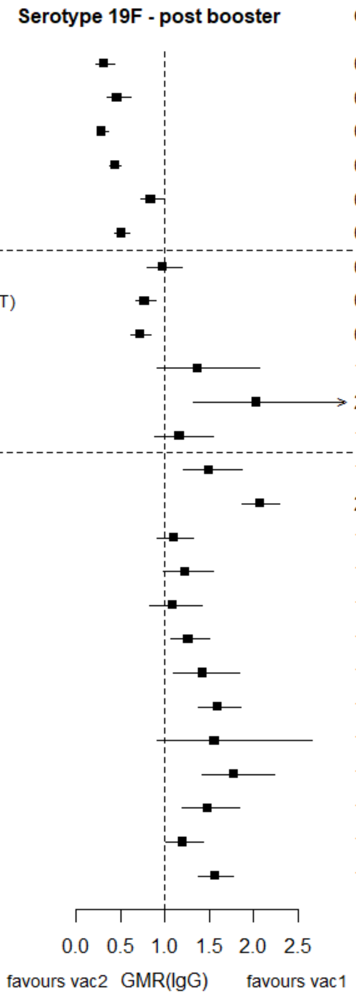

GMR: Geometric mean ratio; pcv: Pneumococcal conjugate vaccine; DTaP – diphtheria and tetanus toxoids, and acellular pertussis vaccine; DTwP – diphtheria and tetanus toxoids, and whole-cell pertussis vaccine; Hib-TT – Haemophilus influenzae type b vaccine (tetanus toxoid conjugate); HB – Hepatitis B vaccine; IPV – Inactivated polio vaccine; OPV – Oral polio vaccine; MenC – Meningococcal C vaccine; TT – tetanus toxoid conjugate; NA: not applicable; enr 3-6 m, 4-8 w int: enrolment at 3-6 months of age, and at 4-8 weeks interval of a 3 doses primary vaccines in total.

Each solid line in the figure shows the GMR from each trial. Black boxes and lines show the point estimates and confidence intervals for geometric mean ratios comparing vac1 vs vac2. Concomitant vaccines are vaccines co-administered with PCV primary vaccine series. Information on co-administered vaccine is not always available. Concomitant vaccines in the bracket are those administered in some but not all of the study sites.

**Supplementary Figure 36. Trial level geometric mean ratios for serotypes 23F at post-booster.**

| ID | Comparison vac1 vs vac2 | Continent | Schedule-prime | Concomitant vaccines | GMR (95%CI) |
|---|---|---|---|---|---|
| 1 | *pcv7 vs pcv10* | Asia | 6-10-24 weeks | DTPw-HBV-Hib-TT + OPV | 1.09 (0.69-1.71) |
| 1 | *pcv7 vs pcv10* | Europe | 2-4-6 months | DTPw-HBV-Hib-TT + IPV | 3.26 (2.18-4.89) |
| 2 | *pcv7 vs pcv10* | Asia | 2-4-6 months | Hib-TT | 3.97 (3.08-5.11) |
| 7 | *pcv7 vs pcv10* | Europe | 2-3-4 months | DTPa-(HBV)-Hib-TT/IPV | 2.14 (1.76-2.61) |
| 8 | *pcv7 vs pcv10* | Europe | 2-3-4 months | DTPa-(HBV)-Hib-TT/IPV | 1.72 (1.44-2.06) |
| 9 | *pcv7 vs pcv10* | Europe | 2-4-6 months | DTPa-(HBV)-Hib-IPV + MenC | 3.19 (2.61-3.90) |
| 4 | *pcv13 vs pcv10* | Europe | 2-3-4 months | DTPa-HBV-Hib-TT/IPV | 1.87 (1.51-2.31) |
| 5 | *pcv13 vs pcv10* | Europe | 2–3–4 months | DTPa-HBV-Hib-TT/IPV + (MenC-TT) | 2.20 (1.86-2.59) |
| 6 | *pcv13 vs pcv10* | Asia | 2-4 months | DTPa-HBV-Hib-TT/IPV | 1.73 (1.45-2.05) |
| 27 | *pcv13 vs pcv10* | Africa | 6 weeks | NA | 1.15 (0.81-1.63) |
| 27 | *pcv13 vs pcv10* | Africa | 14 weeks | NA | 1.09 (0.80-1.48) |
| 27 | *pcv13 vs pcv10* | Africa | 6-14 weeks | NA | 1.75 (1.33-2.32) |
| 10 | *pcv13 vs pcv7* | Asia | 6-10-14 weeks | NA | 0.81 (0.67-0.99) |
| 11 | *pcv13 vs pcv7* | Asia | 2-4-6 months | NA | 0.74 (0.67-0.83) |
| 12 | *pcv13 vs pcv7* | Europe | 3–5 months | DTPa-HBV-Hib-TT/IPV | 0.70 (0.59-0.83) |
| 13 | *pcv13 vs pcv7* | Europe | 2-3-4 months | DTPa-Hib-TT/IPV | 0.77 (0.64-0.94) |
| 14 | *pcv13 vs pcv7* | Asia | 2-4-6 months | NA | 0.69 (0.52-0.92) |
| 15 | *pcv13 vs pcv7* | Europe | 2-3-4 months | DTPa-HBV-Hib-TT/IPV | 0.80 (0.66-0.96) |
| 16 | *pcv13 vs pcv7* | Asia | 2-4-6 months | NA | 1.09 (0.83-1.45) |
| 17 | *pcv13 vs pcv7* | North America | 2-4-6 months | MMR | 0.75 (0.64-0.87) |
| 19 | *pcv13 vs pcv7* | Europe | 2-4 months | Hib-MenC-TT | 0.64 (0.46-0.90) |
| 20 | *pcv13 vs pcv7* | Asia | enr 3-6 m, 4-8 w int | DTPa | 0.83 (0.68-1.03) |
| 21 | *pcv13 vs pcv7* | South America | 2-4-6 months | DTwP-Hib, OPV | 0.86 (0.69-1.07) |
| 22 | *pcv13 vs pcv7* | North America | 2-4-6 months | MMRV, Hib, HAV | 0.65 (0.54-0.77) |
| 23 | *pcv13 vs pcv7* | Asia | 3-4-5 months | NA | 0.94 (0.80-1.09) |

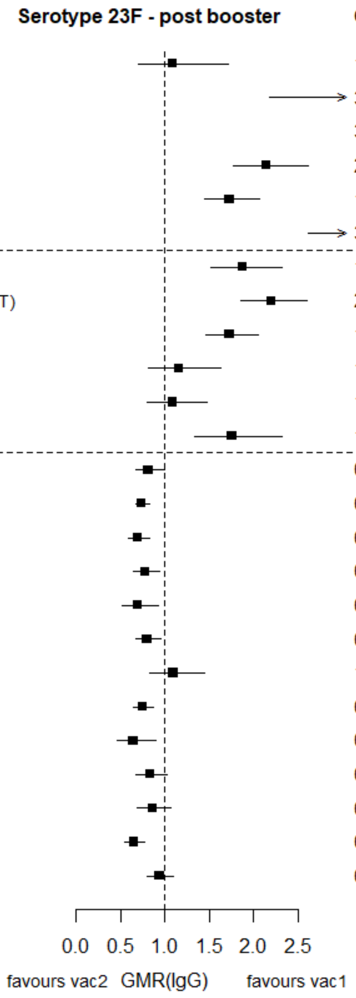

GMR: Geometric mean ratio; pcv: Pneumococcal conjugate vaccine; DTaP – diphtheria and tetanus toxoids, and acellular pertussis vaccine; DTwP – diphtheria and tetanus toxoids, and whole-cell pertussis vaccine; Hib-TT – Haemophilus influenzae type b vaccine (tetanus toxoid conjugate); HB – Hepatitis B vaccine; IPV – Inactivated polio vaccine; OPV – Oral polio vaccine; MenC – Meningococcal C vaccine; TT – tetanus toxoid conjugate; NA: not applicable; enr 3-6 m, 4-8 w int: enrolment at 3-6 months of age, and at 4-8 weeks interval of a 3 doses primary vaccines in total.

Each solid line in the figure shows the GMR from each trial. Black boxes and lines show the point estimates and confidence intervals for geometric mean ratios comparing vac1 vs vac2. Concomitant vaccines are vaccines co-administered with PCV primary vaccine series. Information on co-administered vaccine is not always available. Concomitant vaccines in the bracket are those administered in some but not all of the study sites.

**Supplementary Figure 37. Trial level geometric mean ratios for serotypes 1 at post-booster.**

| ID | Comparison vac1 vs vac2 | Continent | Schedule-prime | Concomitant vaccines | Serotype 1 - post booster | GMR (95%CI) |
|---|---|---|---|---|---|---|
| **4** | *pcv13 vs pcv10* | Europe | 2-3-4 months | DTPa-HBV-Hib-TT/IPV | | 1.55 (1.28-1.89) |
| **5** | *pcv13 vs pcv10* | Europe | 2–3–4 months | DTPa-HBV-Hib-TT/IPV + (MenC-TT) | | 1.74 (1.51-2.00) |
| **6** | *pcv13 vs pcv10* | Asia | 2-4 months | DTPa-HBV-Hib-TT/IPV | | 1.73 (1.48-2.03) |
| **27** | *pcv13 vs pcv10* | Africa | 6 weeks | NA | | 1.86 (1.35-2.56) |
| **27** | *pcv13 vs pcv10* | Africa | 14 weeks | NA | | 2.32 (1.74-3.09) |
| **27** | *pcv13 vs pcv10* | Africa | 6-14 weeks | NA | | 1.25 (0.93-1.68) |

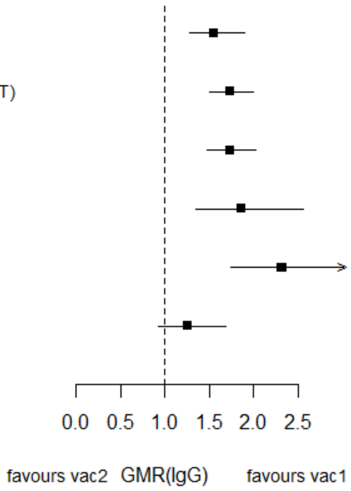

GMR: Geometric mean ratio; pcv: Pneumococcal conjugate vaccine; DTaP – diphtheria and tetanus toxoids, and acellular pertussis vaccine; DTwP – diphtheria and tetanus toxoids, and whole-cell pertussis vaccine; Hib-TT – Haemophilus influenzae type b vaccine (tetanus toxoid conjugate); HB – Hepatitis B vaccine; IPV – Inactivated polio vaccine; OPV – Oral polio vaccine; MenC – Meningococcal C vaccine; TT – tetanus toxoid conjugate; NA: not applicable; enr 3-6 m, 4-8 w int: enrolment at 3-6 months of age, and at 4-8 weeks interval of a 3 doses primary vaccines in total.

Each solid line in the figure shows the GMR from each trial. Black boxes and lines show the point estimates and confidence intervals for geometric mean ratios comparing vac1 vs vac2. Concomitant vaccines are vaccines co-administered with PCV primary vaccine series. Information on co-administered vaccine is not always available. Concomitant vaccines in the bracket are those administered in some but not all of the study sites.

**Supplementary Figure 38. Trial level geometric mean ratios for serotypes 5 at post-booster.**

| ID | Comparison vac1 vs vac2 | Continent | Schedule-prime | Concomitant vaccines | Serotype 5 - post booster | GMR (95%CI) |
|---|---|---|---|---|---|---|
| 4 | *pcv13 vs pcv10* | Europe | 2-3-4 months | DTPa-HBV-Hib-TT/IPV | | 2.19 (1.80-2.68) |
| 5 | *pcv13 vs pcv10* | Europe | 2–3–4 months | DTPa-HBV-Hib-TT/IPV + (MenC-TT) | | 2.30 (1.99-2.65) |
| 6 | *pcv13 vs pcv10* | Asia | 2-4 months | DTPa-HBV-Hib-TT/IPV | | 2.52 (2.21-2.88) |
| 27 | *pcv13 vs pcv10* | Africa | 6 weeks | NA | | 1.19 (0.84-1.68) |
| 27 | *pcv13 vs pcv10* | Africa | 14 weeks | NA | | 1.38 (1.05-1.83) |
| 27 | *pcv13 vs pcv10* | Africa | 6-14 weeks | NA | | 1.63 (1.24-2.15) |

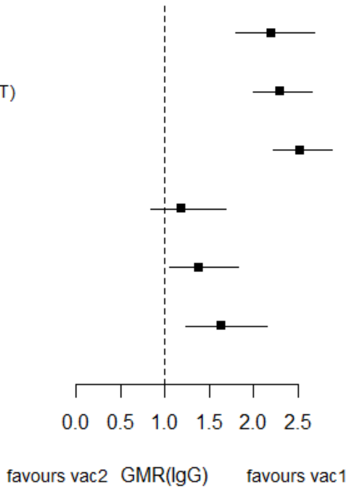

GMR: Geometric mean ratio; pcv: Pneumococcal conjugate vaccine; DTaP – diphtheria and tetanus toxoids, and acellular pertussis vaccine; DTwP – diphtheria and tetanus toxoids, and whole-cell pertussis vaccine; Hib-TT – Haemophilus influenzae type b vaccine (tetanus toxoid conjugate); HB – Hepatitis B vaccine; IPV – Inactivated polio vaccine; OPV – Oral polio vaccine; MenC – Meningococcal C vaccine; TT – tetanus toxoid conjugate; NA: not applicable; enr 3-6 m, 4-8 w int: enrolment at 3-6 months of age, and at 4-8 weeks interval of a 3 doses primary vaccines in total.

Each solid line in the figure shows the GMR from each trial. Black boxes and lines show the point estimates and confidence intervals for geometric mean ratios comparing vac1 vs vac2. Concomitant vaccines are vaccines co-administered with PCV primary vaccine series. Information on co-administered vaccine is not always available. Concomitant vaccines in the bracket are those administered in some but not all of the study sites.

**Supplementary Figure 39. Trial level geometric mean ratios for serotypes 7F at post-booster.**

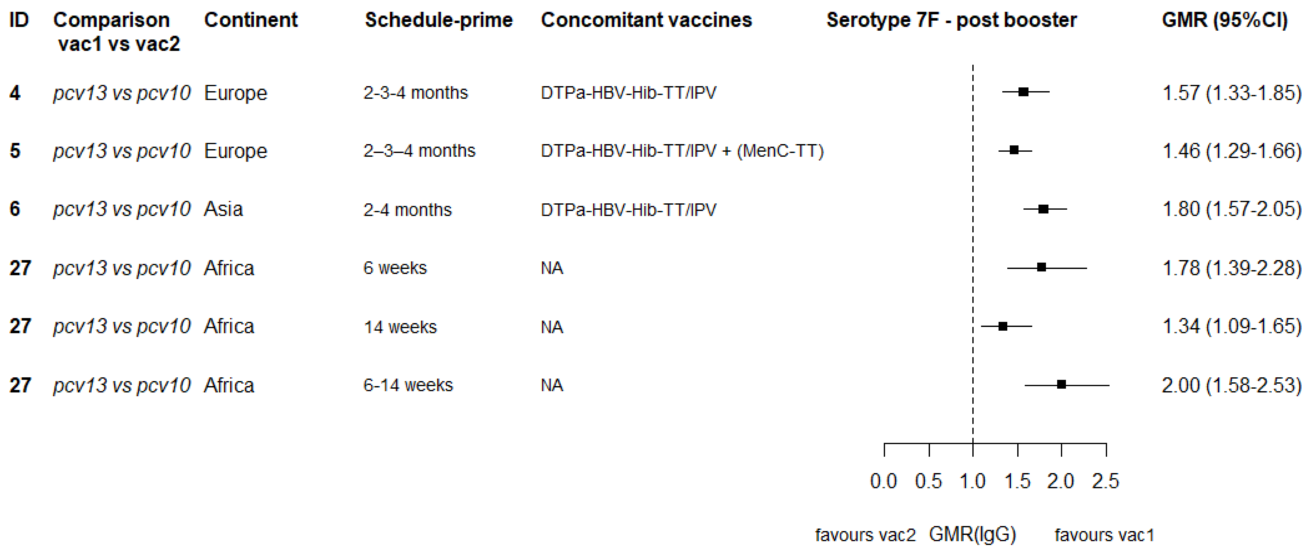

GMR: Geometric mean ratio; pcv: Pneumococcal conjugate vaccine; DTaP – diphtheria and tetanus toxoids, and acellular pertussis vaccine; DTwP – diphtheria and tetanus toxoids, and whole-cell pertussis vaccine; Hib-TT – Haemophilus influenzae type b vaccine (tetanus toxoid conjugate); HB – Hepatitis B vaccine; IPV – Inactivated polio vaccine; OPV – Oral polio vaccine; MenC – Meningococcal C vaccine; TT – tetanus toxoid conjugate; NA: not applicable; enr 3-6 m, 4-8 w int: enrolment at 3-6 months of age, and at 4-8 weeks interval of a 3 doses primary vaccines in total.

Each solid line in the figure shows the GMR from each trial. Black boxes and lines show the point estimates and confidence intervals for geometric mean ratios comparing vac1 vs vac2. Concomitant vaccines are vaccines co-administered with PCV primary vaccine series. Information on co-administered vaccine is not always available. Concomitant vaccines in the bracket are those administered in some but not all of the study sites.

**Supplementary Figure 40. Trial level geometric mean ratios for serotypes 3 at post-booster.**

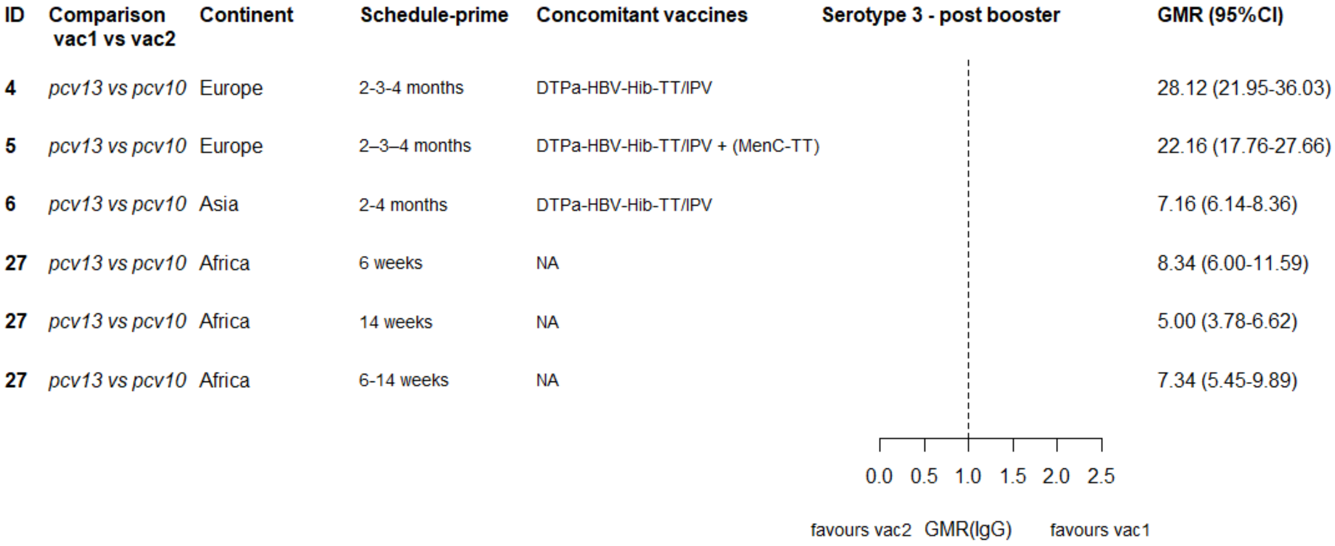

GMR: Geometric mean ratio; pcv: Pneumococcal conjugate vaccine; DTaP – diphtheria and tetanus toxoids, and acellular pertussis vaccine; DTwP – diphtheria and tetanus toxoids, and whole-cell pertussis vaccine; Hib-TT – Haemophilus influenzae type b vaccine (tetanus toxoid conjugate); HB – Hepatitis B vaccine; IPV – Inactivated polio vaccine; OPV – Oral polio vaccine; MenC – Meningococcal C vaccine; TT – tetanus toxoid conjugate; NA: not applicable; enr 3-6 m, 4-8 w int: enrolment at 3-6 months of age, and at 4-8 weeks interval of a 3 doses primary vaccines in total.

Each solid line in the figure shows the GMR from each trial. Black boxes and lines show the point estimates and confidence intervals for geometric mean ratios comparing vac1 vs vac2. Concomitant vaccines are vaccines co-administered with PCV primary vaccine series. Information on co-administered vaccine is not always available. Concomitant vaccines in the bracket are those administered in some but not all of the study sites.

**Supplementary Figure 41. Trial level geometric mean ratios for serotypes 6A at post-booster.**

| ID | Comparison vac1 vs vac2 | Continent | Schedule-prime | Concomitant vaccines | Serotype 6A - post booster | GMR (95%CI) |
|---|---|---|---|---|---|---|
| **4** | *pcv13 vs pcv10* | Europe | 2-3-4 months | DTPa-HBV-Hib-TT/IPV | | 8.07 (6.13-10.62) |
| **5** | *pcv13 vs pcv10* | Europe | 2–3–4 months | DTPa-HBV-Hib-TT/IPV + (MenC-TT) | | 10.30 (8.41-12.61) |
| **6** | *pcv13 vs pcv10* | Asia | 2-4 months | DTPa-HBV-Hib-TT/IPV | | 6.33 (5.21-7.70) |
| **27** | *pcv13 vs pcv10* | Africa | 6 weeks | NA | | 23.61 (16.53-33.72) |
| **27** | *pcv13 vs pcv10* | Africa | 14 weeks | NA | | 21.04 (14.86-29.78) |
| **27** | *pcv13 vs pcv10* | Africa | 6-14 weeks | NA | | 24.32 (16.95-34.90) |

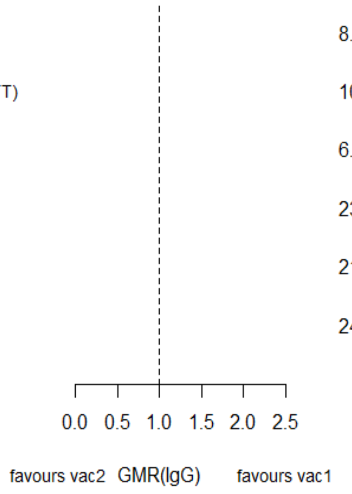

GMR: Geometric mean ratio; pcv: Pneumococcal conjugate vaccine; DTaP – diphtheria and tetanus toxoids, and acellular pertussis vaccine; DTwP – diphtheria and tetanus toxoids, and whole-cell pertussis vaccine; Hib-TT – Haemophilus influenzae type b vaccine (tetanus toxoid conjugate); HB – Hepatitis B vaccine; IPV – Inactivated polio vaccine; OPV – Oral polio vaccine; MenC – Meningococcal C vaccine; TT – tetanus toxoid conjugate; NA: not applicable; enr 3-6 m, 4-8 w int: enrolment at 3-6 months of age, and at 4-8 weeks interval of a 3 doses primary vaccines in total.

Each solid line in the figure shows the GMR from each trial. Black boxes and lines show the point estimates and confidence intervals for geometric mean ratios comparing vac1 vs vac2. Concomitant vaccines are vaccines co-administered with PCV primary vaccine series. Information on co-administered vaccine is not always available. Concomitant vaccines in the bracket are those administered in some but not all of the study sites.

**Supplementary Figure 42. Trial level geometric mean ratios for serotypes 19A at post-booster.**

| ID | Comparison vac1 vs vac2 | Continent | Schedule-prime | Concomitant vaccines | Serotype 19A - post booster | GMR (95%CI) |
|---|---|---|---|---|---|---|
| 4 | *pcv13 vs pcv10* | Europe | 2-3-4 months | DTPa-HBV-Hib-TT/IPV | | 6.65 (5.07-8.73) |
| 5 | *pcv13 vs pcv10* | Europe | 2–3–4 months | DTPa-HBV-Hib-TT/IPV + (MenC-TT) | | 6.07 (4.87-7.57) |
| 6 | *pcv13 vs pcv10* | Asia | 2-4 months | DTPa-HBV-Hib-TT/IPV | | 5.22 (4.39-6.20) |
| 27 | *pcv13 vs pcv10* | Africa | 6 weeks | NA | | 14.16 (9.52-21.06) |
| 27 | *pcv13 vs pcv10* | Africa | 14 weeks | NA | | 12.19 (8.23-18.05) |
| 27 | *pcv13 vs pcv10* | Africa | 6-14 weeks | NA | | 11.40 (7.80-16.66) |

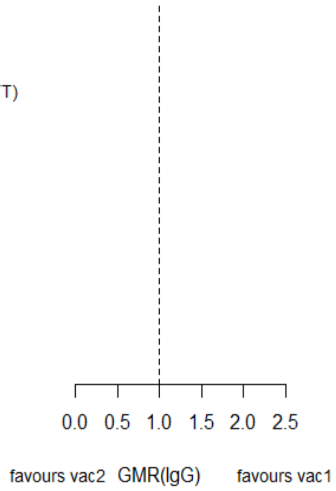

GMR: Geometric mean ratio; pcv: Pneumococcal conjugate vaccine; DTaP – diphtheria and tetanus toxoids, and acellular pertussis vaccine; DTwP – diphtheria and tetanus toxoids, and whole-cell pertussis vaccine; Hib-TT – Haemophilus influenzae type b vaccine (tetanus toxoid conjugate); HB – Hepatitis B vaccine; IPV – Inactivated polio vaccine; OPV – Oral polio vaccine; MenC – Meningococcal C vaccine; TT – tetanus toxoid conjugate; NA: not applicable; enr 3-6 m, 4-8 w int: enrolment at 3-6 months of age, and at 4-8 weeks interval of a 3 doses primary vaccines in total.

Each solid line in the figure shows the GMR from each trial. Black boxes and lines show the point estimates and confidence intervals for geometric mean ratios comparing vac1 vs vac2. Concomitant vaccines are vaccines co-administered with PCV primary vaccine series. Information on co-administered vaccine is not always available. Concomitant vaccines in the bracket are those administered in some but not all of the study sites.

**Supplementary Figure 43. Sensitivity analysis on geometric mean ratios comparing PCV13 vs PCV10, PCV7 vs PCV10, and PCV13 vs PCV7 at a) post-primary vaccination series, b) pre-boost, and c) post-boost by including studies providing data for all three time points.**

| | Serotype | No. cohorts | Direct Evidence (%) | GMR (95% CI) PCV13 vs PCV10 | GMR (95% CI) PCV7 vs PCV10 | GMR (95% CI) PCV13 vs PCV7 |
|---|---|---|---|---|---|---|
| PCV7 | 4 | 14 | 62 | 1.01 (0.83, 1.23) | 1.49 (1.23, 1.81) | 0.68 (0.54, 0.85) |
| | 6B | 14 | 76 | 0.80 (0.66, 0.97) | 1.36 (1.12, 1.66) | 0.59 (0.47, 0.73) |
| | 9V | 14 | 63 | 1.15 (0.95, 1.41) | 1.63 (1.34, 1.99) | 0.71 (0.56, 0.89) |
| | 14 | 14 | 64 | 1.01 (0.85, 1.21) | 1.32 (1.11, 1.57) | 0.77 (0.63, 0.94) |
| | 18C | 14 | 62 | 1.18 (0.87, 1.61) | 1.03 (0.75, 1.42) | 1.15 (0.80, 1.65) |
| | 19F | 14 | 65 | 0.93 (0.63, 1.37) | 0.94 (0.63, 1.42) | 0.99 (0.62, 1.56) |
| | 23F | 14 | 68 | 1.38 (1.09, 1.74) | 1.86 (1.46, 2.37) | 0.74 (0.56, 0.97) |
| PCV10 | 1 | 6 | 100 | 1.11 (0.72, 1.70) | 0.02 (0.02, 0.03) | 58.61 (42.73, 80.39) |
| | 5 | 6 | 100 | 0.92 (0.60, 1.40) | 0.03 (0.01, 0.05) | 19.07 (9.66, 37.67) |
| | 7F | 6 | 100 | 1.32 (1.13, 1.55) | 0.02 (0.01, 0.02) | 66.25 (52.26, 84.00) |
| PCV13 | 3 | 6 | 100 | 15.28 (7.54, 30.98) | 0.92 (0.29, 2.85) | 16.81 (9.03, 31.31) |
| | 6A | 6 | 100 | 5.41 (1.94, 15.06) | 1.01 (0.52, 1.96) | 6.21 (4.39, 8.80) |
| | 19A | 6 | 100 | 6.23 (2.94, 13.18) | 0.94 (0.52, 1.67) | 2.75 (1.46, 5.21) |

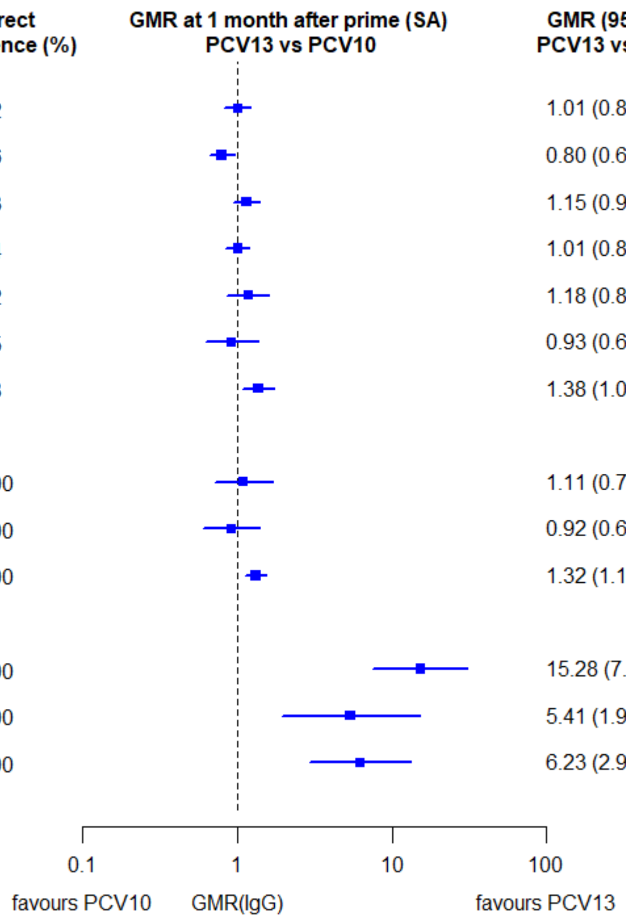

a)

| | Serotype | No. cohorts | Direct Evidence (%) | GMR (95% CI) PCV13 vs PCV10 | GMR (95% CI) PCV7 vs PCV10 | GMR (95% CI) PCV13 vs PCV7 |
|---|---|---|---|---|---|---|
| **PCV7** | **4** | 17 | 69 | 0.66 (0.56, 0.78) | 0.95 (0.80, 1.12) | 0.70 (0.58, 0.84) |
| | **6B** | 17 | 74 | 0.41 (0.32, 0.51) | 0.53 (0.42, 0.68) | 0.76 (0.58, 0.98) |
| | **9V** | 17 | 68 | 0.74 (0.62, 0.89) | 0.96 (0.80, 1.16) | 0.77 (0.63, 0.95) |
| | **14** | 17 | 65 | 1.26 (1.13, 1.40) | 1.53 (1.37, 1.71) | 0.82 (0.73, 0.92) |
| | **18C** | 17 | 66 | 0.78 (0.67, 0.91) | 0.77 (0.66, 0.90) | 1.01 (0.85, 1.19) |
| | **19F** | 17 | 79 | 0.57 (0.46, 0.70) | 0.38 (0.30, 0.48) | 1.48 (1.16, 1.88) |
| | **23F** | 17 | 73 | 0.64 (0.54, 0.77) | 0.81 (0.67, 0.97) | 0.80 (0.65, 0.97) |
| **PCV10** | **1** | 7 | 100 | 1.36 (1.09, 1.71) | 0.13 (0.10, 0.16) | 12.48 (9.83, 15.84) |
| | **5** | 7 | 100 | 1.30 (1.09, 1.56) | 0.18 (0.11, 0.29) | 4.43 (2.56, 7.68) |
| | **7F** | 7 | 100 | 1.18 (1.01, 1.38) | 0.05 (0.04, 0.06) | 20.43 (17.11, 24.39) |
| **PCV13** | **3** | 7 | 100 | 2.70 (1.78, 4.09) | 0.95 (0.51, 1.76) | 2.90 (2.07, 4.06) |
| | **6A** | 7 | 100 | 2.11 (1.50, 2.97) | 0.75 (0.58, 0.96) | 2.77 (2.16, 3.55) |
| | **19A** | 7 | 100 | 2.23 (1.76, 2.82) | 0.63 (0.46, 0.87) | 1.50 (1.28, 1.76) |

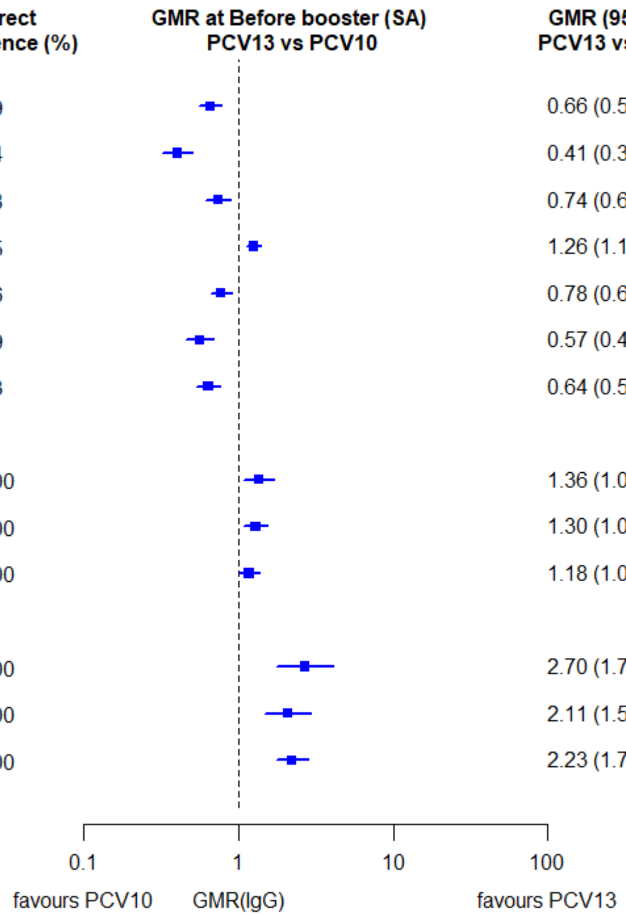

b)

| | Serotype | No. cohorts | Direct Evidence (%) | GMR (95% CI) PCV13 vs PCV10 | GMR (95% CI) PCV7 vs PCV10 | GMR (95% CI) PCV13 vs PCV7 |
|---|---|---|---|---|---|---|
| PCV7 | 4 | 14 | 73 | 1.04 (0.93, 1.15) | 1.38 (1.23, 1.54) | 0.75 (0.67, 0.85) |
| | 6B | 14 | 73 | 1.25 (1.01, 1.55) | 1.63 (1.30, 2.04) | 0.77 (0.60, 0.99) |
| | 9V | 14 | 72 | 1.40 (1.18, 1.67) | 1.68 (1.40, 2.02) | 0.83 (0.68, 1.02) |
| | 14 | 14 | 69 | 1.43 (1.22, 1.66) | 1.72 (1.47, 2.00) | 0.83 (0.70, 0.99) |
| | 18C | 14 | 73 | 0.78 (0.66, 0.93) | 0.89 (0.74, 1.07) | 0.88 (0.72, 1.08) |
| | 19F | 14 | 74 | 0.93 (0.74, 1.15) | 0.56 (0.45, 0.70) | 1.64 (1.27, 2.12) |
| | 23F | 14 | 76 | 1.60 (1.33, 1.92) | 2.22 (1.81, 2.71) | 0.72 (0.58, 0.89) |
| PCV10 | 1 | 6 | 100 | 1.71 (1.51, 1.93) | 0.02 (0.01, 0.03) | 125.80 (71.00, 222.91) |
| | 5 | 6 | 100 | 1.87 (1.52, 2.30) | 0.04 (0.02, 0.10) | 25.12 (9.55, 66.10) |
| | 7F | 6 | 100 | 1.63 (1.46, 1.82) | 0.01 (0.01, 0.02) | 115.07 (73.61, 179.87) |
| PCV13 | 3 | 6 | 100 | 10.56 (6.03, 18.49) | 0.76 (0.33, 1.78) | 13.92 (12.19, 15.90) |
| | 6A | 6 | 100 | 13.40 (8.48, 21.16) | 1.98 (1.38, 2.83) | 4.48 (3.24, 6.21) |
| | 19A | 6 | 100 | 8.36 (6.08, 11.49) | 0.65 (0.36, 1.16) | 2.46 (2.23, 2.73) |

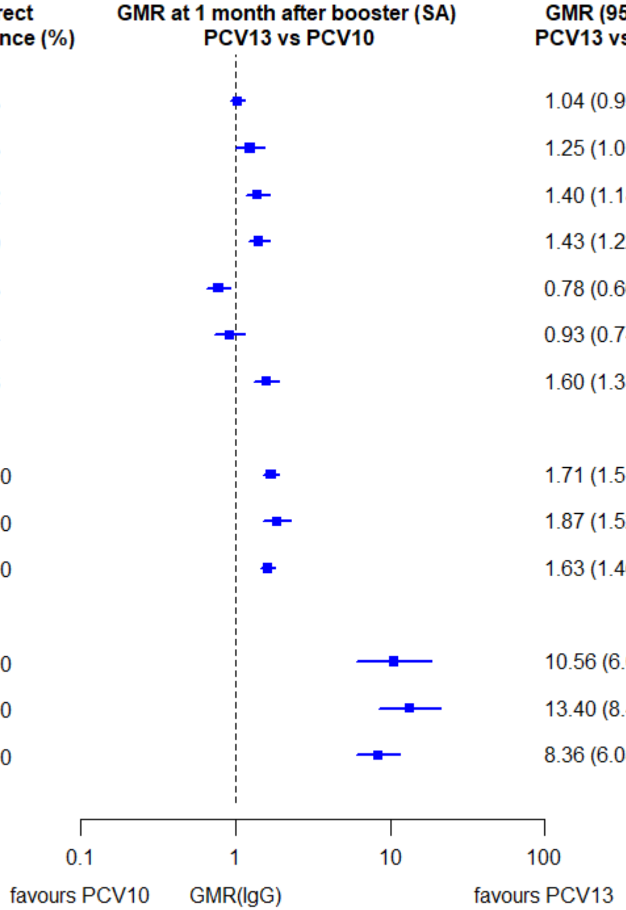

c)

GMR: Geometric mean ratio; PCV: Pneumococcal conjugate vaccine. Each line in the figure shows the output from a network meta-analyses (PCV7 serotypes) or direct meta-analyses (PCV13 but non-PCV7 serotypes). Blue boxes and blue lines show the point estimates and confidence intervals for geometric mean ratios comparing PCV13 vs PCV10. Points to the right of the vertical line are those with higher antibody responses in the PCV13 arm of the study, and points to the left are those with higher antibody responses in the PCV10 arm. The direct evidence column shows the percentage of evidence from studies directly comparing PCV13 vs PCV10 that contributes to the estimates presented in the figure in blue (PCV13 vs PCV10). GMR of PCV13 vs PCV10 for PCV10 and PCV13 serotypes are from a meta-analysis of only head-to-head studies of PCV13 vs PCV10.

**Supplementary Figure 44. Direct and indirect evidence on relative risk comparing PCV13 vs PCV10 for serotypes in PCV7 (4, 6B, 9V, 14, 18C, 19F and 23F)**

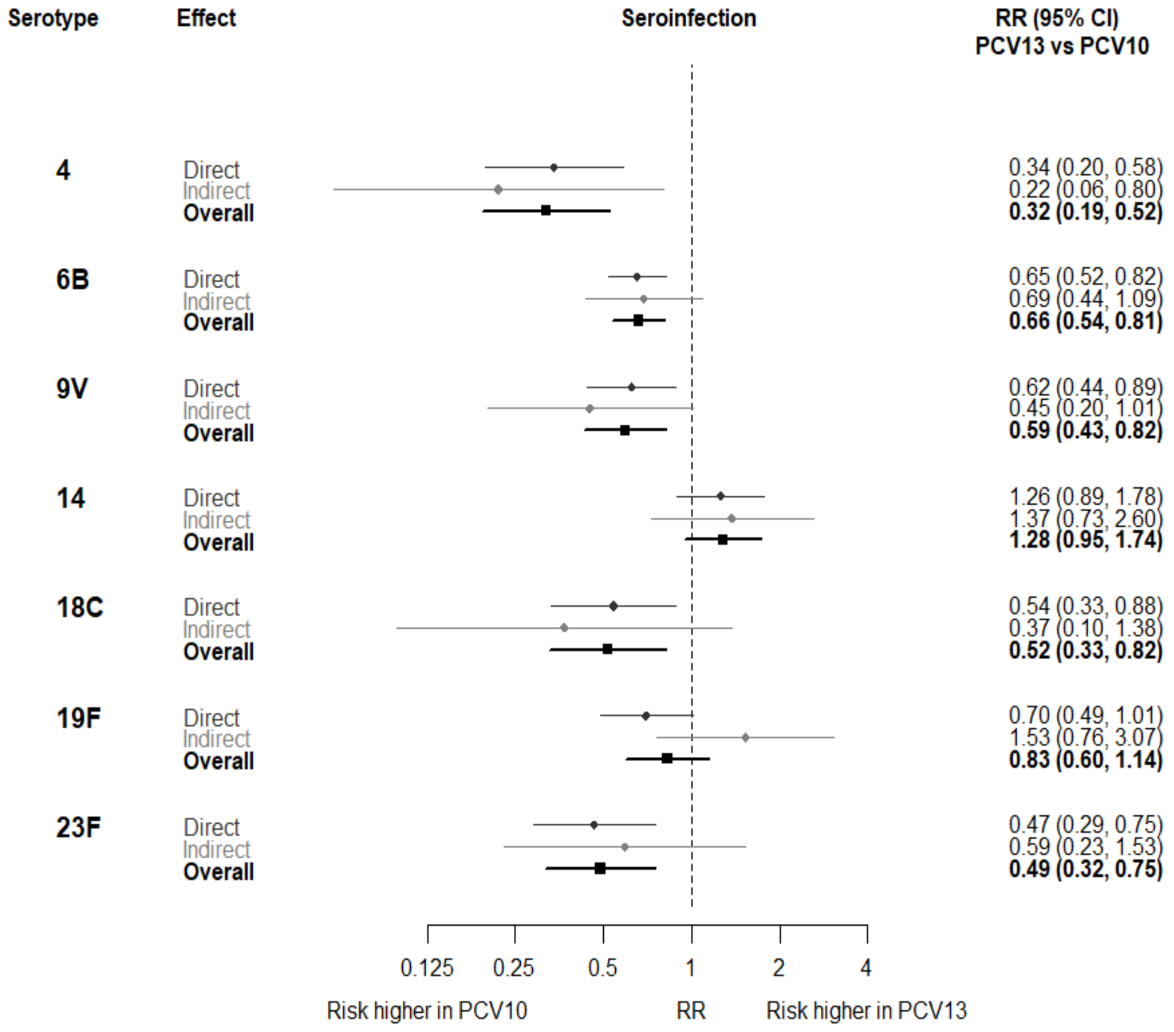

RR: Relative risk; PCV: Pneumococcal conjugate vaccine. Each line in the figure shows the output from a network meta-analyses. Dark grey diamonds and lines show the point estimates and confidence intervals relative risks from studies directly comparing PCV13 vs PCV10. Light grey diamonds and lines show the point estimates and confidence intervals for relative risks from studies comparing PCV13 vs PCV10 through PCV7. Black boxes and lines show the point estimates and confidence intervals incorporating both direct and indirect evidence.

**Supplementary Figure 45. Trial level relative risk for serotype 4.**

| ID | Comparison vac1 vs vac2 | Continent | Schedule-prime | co-adm vaccines | RR (95%CI) |
|---|---|---|---|---|---|
| 1 | *pcv7 vs pcv10* | Asia | 6-10-24 weeks | DTPw-HBV-Hib-TT + OPV | 0.20 (0.01-3.39) |
| 1 | *pcv7 vs pcv10* | Europe | 2-4-6 months | DTPw-HBV-Hib-TT + IPV | 0.24 (0.01-4.25) |
| 3 | *pcv7 vs pcv10* | Europe | 2-3-4 months | DTPa-HBV-Hib-TT/IPV | 0.15 (0.01-2.91) |
| 7 | *pcv7 vs pcv10* | Europe | 2-3-4 months | DTPa-(HBV)-Hib-TT/IPV | 0.06 (0.00-1.00) |
| 8 | *pcv7 vs pcv10* | Europe | 2-3-4 months | DTPa-(HBV)-Hib-TT/IPV | 0.44 (0.16-1.24) |
| 9 | *pcv7 vs pcv10* | Europe | 2-4-6 months | DTPa-(HBV)-Hib-TT/IPV + MenC | 0.16 (0.02-1.21) |
| 4 | *pcv13 vs pcv10* | Europe | 2-3-4 months | DTPa-HBV-Hib-TT/IPV | 0.30 (0.10-0.89) |
| 27 | *pcv13 vs pcv10* | Africa | 14 weeks | DTPa-HBV-Hib-TT/IPV/Rotavirus | 0.70 (0.40-1.20) |
| 27 | *pcv13 vs pcv10* | Africa | 6-14 weeks | DTPa-HBV-Hib-TT/IPV/Rotavirus | 0.97 (0.20-4.67) |
| 27 | *pcv13 vs pcv10* | Africa | 6 weeks | DTPa-HBV-Hib-TT/IPV/Rotavirus | 0.22 (0.09-0.54) |
| 5 | *pcv13 vs pcv10* | Europe | 2–3–4 months | DTPa-HBV-Hib-TT/IPV + (MenC-TT) | 0.08 (0.02-0.35) |
| 6 | *pcv13 vs pcv10* | Asia | 2-4 months | DTPa-HBV-Hib-TT/IPV | 0.10 (0.01-0.80) |
| 15 | *pcv13 vs pcv7* | Europe | 2-3-4 months | DTPa-HBV-Hib-TT/IPV | 0.67 (0.19-2.34) |
| 19 | *pcv13 vs pcv7* | Europe | 2-4 months | DTaP-IPV-Hib/MenC | 0.77 (0.02-37.81) |
| 22 | *pcv13 vs pcv7* | North America | 2-4-6 months | DTPa-HBV-Hib-TT/IPV | 1.11 (0.38-3.24) |

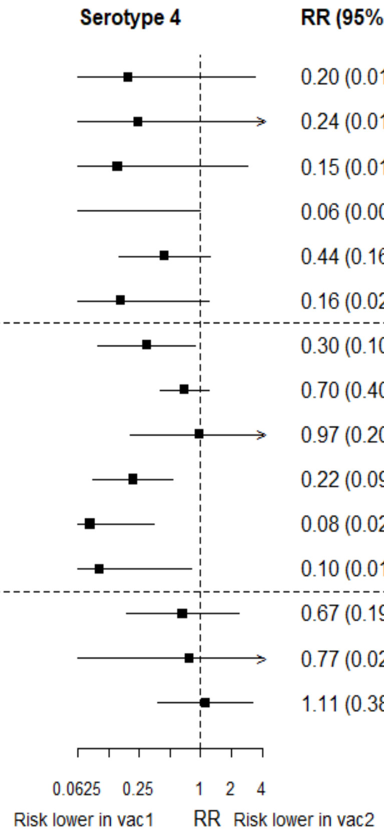

RR: relative risk; pcv: Pneumococcal conjugate vaccine; DTaP – diphtheria and tetanus toxoids, and acellular pertussis vaccine; DTwP – diphtheria and tetanus toxoids, and whole-cell pertussis vaccine; Hib-TT – Haemophilus influenzae type b vaccine (tetanus toxoid conjugate); HB – Hepatitis B vaccine; IPV – Inactivated polio vaccine; OPV – Oral polio vaccine; MenC – Meningococcal C vaccine; TT – tetanus toxoid conjugate; NA: not applicable.

Each solid line in the figure shows the RR from each trial. Black boxes and lines show the point estimates and confidence intervals for relative risks comparing vac1 vs vac2. co-adm vaccines are vaccines co-administered with PCV primary vaccine series. Information on co-administered vaccine is not always available. Concomitant vaccines in the bracket are those administered in some but not all of the study sites.

**Supplementary Figure 46. Trial level relative risk for serotype 6B.**

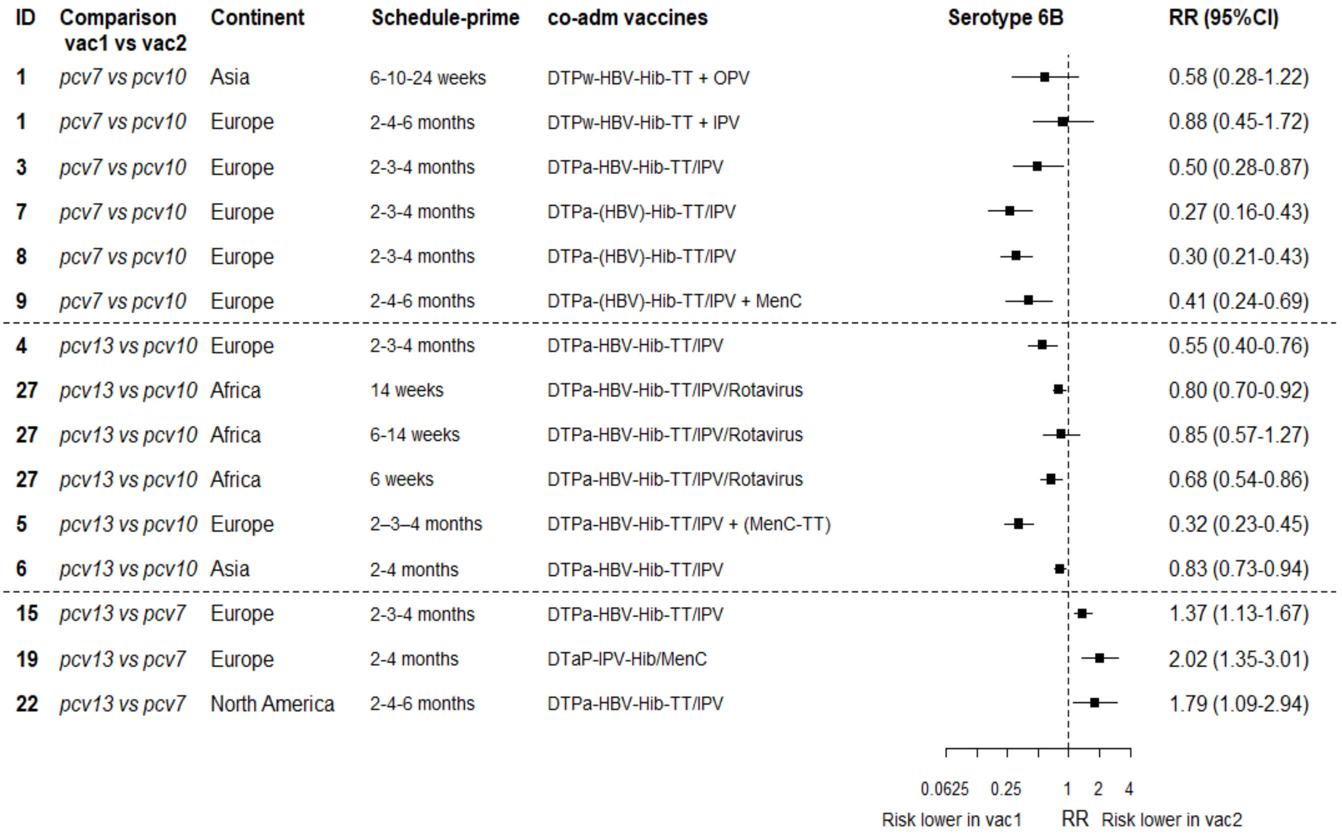

RR: relative risk; pcv: Pneumococcal conjugate vaccine; DTaP – diphtheria and tetanus toxoids, and acellular pertussis vaccine; DTwP – diphtheria and tetanus toxoids, and whole-cell pertussis vaccine; Hib-TT – Haemophilus influenzae type b vaccine (tetanus toxoid conjugate); HB – Hepatitis B vaccine; IPV – Inactivated polio vaccine; OPV – Oral polio vaccine; MenC – Meningococcal C vaccine; TT – tetanus toxoid conjugate; NA: not applicable.

Each solid line in the figure shows the RR from each trial. Black boxes and lines show the point estimates and confidence intervals for relative risks comparing vac1 vs vac2. co-adm vaccines are vaccines co-administered with PCV primary vaccine series. Information on co-administered vaccine is not always available. Concomitant vaccines in the bracket are those administered in some but not all of the study sites.

**Supplementary Figure 47. Trial level relative risk for serotype 9V.**

| ID | Comparison vac1 vs vac2 | Continent | Schedule-prime | co-adm vaccines | RR (95%CI) |
|---|---|---|---|---|---|
| 1 | *pcv7 vs pcv10* | Asia | 6-10-24 weeks | DTPw-HBV-Hib-TT + OPV | 0.24 (0.03-1.75) |
| 1 | *pcv7 vs pcv10* | Europe | 2-4-6 months | DTPw-HBV-Hib-TT + IPV | 0.28 (0.04-2.10) |
| 3 | *pcv7 vs pcv10* | Europe | 2-3-4 months | DTPa-HBV-Hib-TT/IPV | 0.47 (0.13-1.72) |
| 7 | *pcv7 vs pcv10* | Europe | 2-3-4 months | DTPa-(HBV)-Hib-TT/IPV | 0.25 (0.11-0.58) |
| 8 | *pcv7 vs pcv10* | Europe | 2-3-4 months | DTPa-(HBV)-Hib-TT/IPV | 0.56 (0.33-0.95) |
| 9 | *pcv7 vs pcv10* | Europe | 2-4-6 months | DTPa-(HBV)-Hib-TT/IPV + MenC | 0.48 (0.24-0.95) |
| 4 | *pcv13 vs pcv10* | Europe | 2-3-4 months | DTPa-HBV-Hib-TT/IPV | 0.62 (0.26-1.44) |
| 27 | *pcv13 vs pcv10* | Africa | 14 weeks | DTPa-HBV-Hib-TT/IPV/Rotavirus | 1.00 (0.76-1.32) |
| 27 | *pcv13 vs pcv10* | Africa | 6-14 weeks | DTPa-HBV-Hib-TT/IPV/Rotavirus | 1.06 (0.50-2.26) |
| 27 | *pcv13 vs pcv10* | Africa | 6 weeks | DTPa-HBV-Hib-TT/IPV/Rotavirus | 0.51 (0.32-0.80) |
| 5 | *pcv13 vs pcv10* | Europe | 2–3–4 months | DTPa-HBV-Hib-TT/IPV + (MenC-TT) | 0.29 (0.14-0.61) |
| 6 | *pcv13 vs pcv10* | Asia | 2-4 months | DTPa-HBV-Hib-TT/IPV | 0.42 (0.20-0.90) |
| 15 | *pcv13 vs pcv7* | Europe | 2-3-4 months | DTPa-HBV-Hib-TT/IPV | 0.77 (0.34-1.73) |
| 19 | *pcv13 vs pcv7* | Europe | 2-4 months | DTaP-IPV-Hib/MenC | 2.30 (0.25-21.18) |
| 22 | *pcv13 vs pcv7* | North America | 2-4-6 months | DTPa-HBV-Hib-TT/IPV | 1.28 (0.62-2.63) |

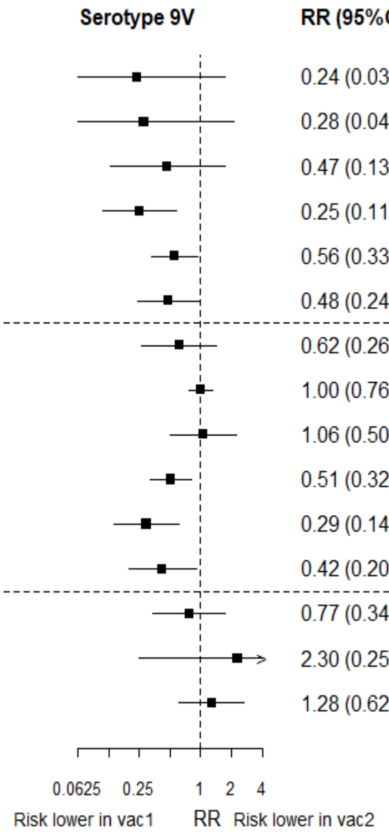

RR: relative risk; pcv: Pneumococcal conjugate vaccine; DTaP – diphtheria and tetanus toxoids, and acellular pertussis vaccine; DTwP – diphtheria and tetanus toxoids, and whole-cell pertussis vaccine; Hib-TT – Haemophilus influenzae type b vaccine (tetanus toxoid conjugate); HB – Hepatitis B vaccine; IPV – Inactivated polio vaccine; OPV – Oral polio vaccine; MenC – Meningococcal C vaccine; TT – tetanus toxoid conjugate; NA: not applicable.

Each solid line in the figure shows the RR from each trial. Black boxes and lines show the point estimates and confidence intervals for relative risks comparing vac1 vs vac2. co-adm vaccines are vaccines co-administered with PCV primary vaccine series. Information on co-administered vaccine is not always available. Concomitant vaccines in the bracket are those administered in some but not all of the study sites.

**Supplementary Figure 48. Trial level relative risk for serotype 14.**

| ID | Comparison vac1 vs vac2 | Continent | Schedule-prime | co-adm vaccines | RR (95%CI) |
|---|---|---|---|---|---|
| 1 | *pcv7 vs pcv10* | Asia | 6-10-24 weeks | DTPw-HBV-Hib-TT + OPV | 1.71 (0.88-3.32) |
| 1 | *pcv7 vs pcv10* | Europe | 2-4-6 months | DTPw-HBV-Hib-TT + IPV | 0.22 (0.03-1.65) |
| 3 | *pcv7 vs pcv10* | Europe | 2-3-4 months | DTPa-HBV-Hib-TT/IPV | 1.25 (0.49-3.21) |
| 7 | *pcv7 vs pcv10* | Europe | 2-3-4 months | DTPa-(HBV)-Hib-TT/IPV | 0.86 (0.52-1.40) |
| 8 | *pcv7 vs pcv10* | Europe | 2-3-4 months | DTPa-(HBV)-Hib-TT/IPV | 1.11 (0.73-1.68) |
| 9 | *pcv7 vs pcv10* | Europe | 2-4-6 months | DTPa-(HBV)-Hib-TT/IPV + MenC | 1.82 (1.07-3.11) |
| 4 | *pcv13 vs pcv10* | Europe | 2-3-4 months | DTPa-HBV-Hib-TT/IPV | 1.16 (0.69-1.94) |
| 27 | *pcv13 vs pcv10* | Africa | 14 weeks | DTPa-HBV-Hib-TT/IPV/Rotavirus | 0.73 (0.57-0.94) |
| 27 | *pcv13 vs pcv10* | Africa | 6-14 weeks | DTPa-HBV-Hib-TT/IPV/Rotavirus | 4.73 (2.24-9.99) |
| 27 | *pcv13 vs pcv10* | Africa | 6 weeks | DTPa-HBV-Hib-TT/IPV/Rotavirus | 1.17 (0.76-1.79) |
| 5 | *pcv13 vs pcv10* | Europe | 2–3–4 months | DTPa-HBV-Hib-TT/IPV + (MenC-TT) | 1.96 (1.15-3.33) |
| 6 | *pcv13 vs pcv10* | Asia | 2-4 months | DTPa-HBV-Hib-TT/IPV | 0.86 (0.62-1.19) |
| 15 | *pcv13 vs pcv7* | Europe | 2-3-4 months | DTPa-HBV-Hib-TT/IPV | 1.11 (0.84-1.46) |
| 19 | *pcv13 vs pcv7* | Europe | 2-4 months | DTaP-IPV-Hib/MenC | 1.72 (0.85-3.51) |
| 22 | *pcv13 vs pcv7* | North America | 2-4-6 months | DTPa-HBV-Hib-TT/IPV | 0.89 (0.56-1.43) |

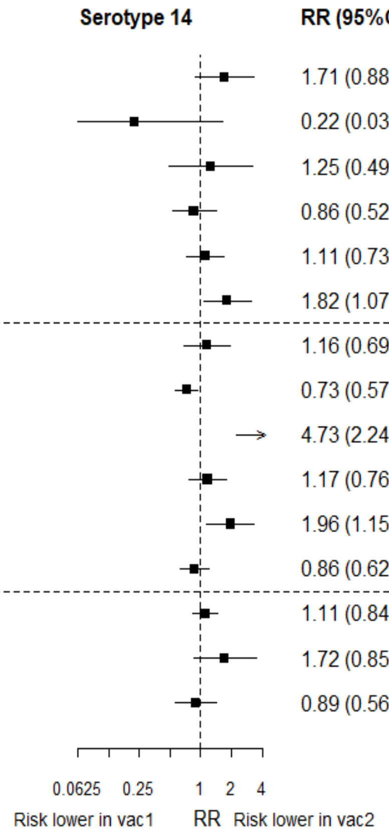

RR: relative risk; pcv: Pneumococcal conjugate vaccine; DTaP – diphtheria and tetanus toxoids, and acellular pertussis vaccine; DTwP – diphtheria and tetanus toxoids, and whole-cell pertussis vaccine; Hib-TT – Haemophilus influenzae type b vaccine (tetanus toxoid conjugate); HB – Hepatitis B vaccine; IPV – Inactivated polio vaccine; OPV – Oral polio vaccine; MenC – Meningococcal C vaccine; TT – tetanus toxoid conjugate; NA: not applicable.

Each solid line in the figure shows the RR from each trial. Black boxes and lines show the point estimates and confidence intervals for relative risks comparing vac1 vs vac2. co-adm vaccines are vaccines co-administered with PCV primary vaccine series. Information on co-administered vaccine is not always available. Concomitant vaccines in the bracket are those administered in some but not all of the study sites.

**Supplementary Figure 49. Trial level relative risk for serotype 18C.**

| ID | Comparison vac1 vs vac2 | Continent | Schedule-prime | co-adm vaccines | RR (95%CI) |
|---|---|---|---|---|---|
| 1 | *pcv7 vs pcv10* | Asia | 6-10-24 weeks | DTPw-HBV-Hib-TT + OPV | 0.50 (0.06-4.04) |
| 1 | *pcv7 vs pcv10* | Europe | 2-4-6 months | DTPw-HBV-Hib-TT + IPV | 2.30 (0.53-9.90) |
| 3 | *pcv7 vs pcv10* | Europe | 2-3-4 months | DTPa-HBV-Hib-TT/IPV | 0.56 (0.05-5.98) |
| 7 | *pcv7 vs pcv10* | Europe | 2-3-4 months | DTPa-(HBV)-Hib-TT/IPV | 0.20 (0.07-0.55) |
| 8 | *pcv7 vs pcv10* | Europe | 2-3-4 months | DTPa-(HBV)-Hib-TT/IPV | 0.26 (0.08-0.85) |
| 9 | *pcv7 vs pcv10* | Europe | 2-4-6 months | DTPa-(HBV)-Hib-TT/IPV + MenC | 0.28 (0.07-1.18) |
| 4 | *pcv13 vs pcv10* | Europe | 2-3-4 months | DTPa-HBV-Hib-TT/IPV | 0.67 (0.26-1.71) |
| 27 | *pcv13 vs pcv10* | Africa | 14 weeks | DTPa-HBV-Hib-TT/IPV/Rotavirus | 0.94 (0.65-1.36) |
| 27 | *pcv13 vs pcv10* | Africa | 6-14 weeks | DTPa-HBV-Hib-TT/IPV/Rotavirus | 0.42 (0.17-1.03) |
| 27 | *pcv13 vs pcv10* | Africa | 6 weeks | DTPa-HBV-Hib-TT/IPV/Rotavirus | 0.86 (0.52-1.44) |
| 5 | *pcv13 vs pcv10* | Europe | 2–3–4 months | DTPa-HBV-Hib-TT/IPV + (MenC-TT) | 0.28 (0.12-0.64) |
| 6 | *pcv13 vs pcv10* | Asia | 2-4 months | DTPa-HBV-Hib-TT/IPV | 0.20 (0.07-0.56) |
| 15 | *pcv13 vs pcv7* | Europe | 2-3-4 months | DTPa-HBV-Hib-TT/IPV | 0.60 (0.15-2.50) |
| 19 | *pcv13 vs pcv7* | Europe | 2-4 months | DTaP-IPV-Hib/MenC | 0.25 (0.01-5.86) |
| 22 | *pcv13 vs pcv7* | North America | 2-4-6 months | DTPa-HBV-Hib-TT/IPV | 1.91 (0.48-7.53) |

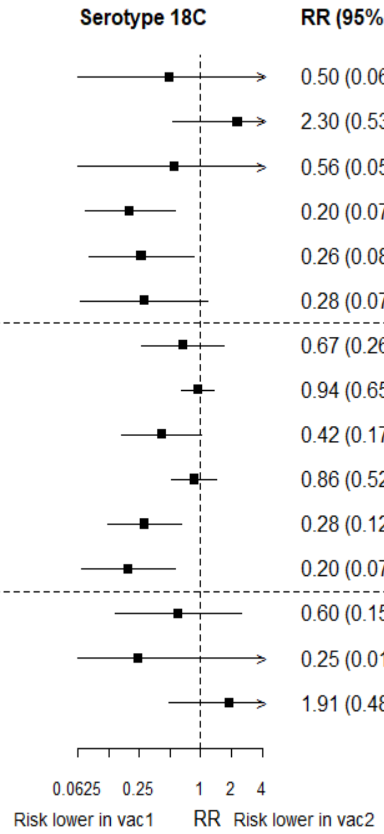

RR: relative risk; pcv: Pneumococcal conjugate vaccine; DTaP – diphtheria and tetanus toxoids, and acellular pertussis vaccine; DTwP – diphtheria and tetanus toxoids, and whole-cell pertussis vaccine; Hib-TT – Haemophilus influenzae type b vaccine (tetanus toxoid conjugate); HB – Hepatitis B vaccine; IPV – Inactivated polio vaccine; OPV – Oral polio vaccine; MenC – Meningococcal C vaccine; TT – tetanus toxoid conjugate; NA: not applicable.

Each solid line in the figure shows the RR from each trial. Black boxes and lines show the point estimates and confidence intervals for relative risks comparing vac1 vs vac2. co-adm vaccines are vaccines co-administered with PCV primary vaccine series. Information on co-administered vaccine is not always available. Concomitant vaccines in the bracket are those administered in some but not all of the study sites.

**Supplementary Figure 50. Trial level relative risk for serotype 4.**

| ID | Comparison vac1 vs vac2 | Continent | Schedule-prime | co-adm vaccines | RR (95%CI) |
|---|---|---|---|---|---|
| 1 | *pcv7 vs pcv10* | Asia | 6-10-24 weeks | DTPw-HBV-Hib-TT + OPV | 1.50 (0.39-5.76) |
| 1 | *pcv7 vs pcv10* | Europe | 2-4-6 months | DTPw-HBV-Hib-TT + IPV | 1.95 (0.80-4.74) |
| 3 | *pcv7 vs pcv10* | Europe | 2-3-4 months | DTPa-HBV-Hib-TT/IPV | 0.55 (0.18-1.71) |
| 7 | *pcv7 vs pcv10* | Europe | 2-3-4 months | DTPa-(HBV)-Hib-TT/IPV | 0.56 (0.32-0.96) |
| 8 | *pcv7 vs pcv10* | Europe | 2-3-4 months | DTPa-(HBV)-Hib-TT/IPV | 0.78 (0.51-1.21) |
| 9 | *pcv7 vs pcv10* | Europe | 2-4-6 months | DTPa-(HBV)-Hib-TT/IPV + MenC | 1.62 (0.85-3.10) |
| 4 | *pcv13 vs pcv10* | Europe | 2-3-4 months | DTPa-HBV-Hib-TT/IPV | 1.27 (0.58-2.80) |
| 27 | *pcv13 vs pcv10* | Africa | 14 weeks | DTPa-HBV-Hib-TT/IPV/Rotavirus | 0.47 (0.31-0.70) |
| 27 | *pcv13 vs pcv10* | Africa | 6-14 weeks | DTPa-HBV-Hib-TT/IPV/Rotavirus | 0.78 (0.39-1.55) |
| 27 | *pcv13 vs pcv10* | Africa | 6 weeks | DTPa-HBV-Hib-TT/IPV/Rotavirus | 0.89 (0.51-1.55) |
| 5 | *pcv13 vs pcv10* | Europe | 2–3–4 months | DTPa-HBV-Hib-TT/IPV + (MenC-TT) | 0.56 (0.28-1.12) |
| 6 | *pcv13 vs pcv10* | Asia | 2-4 months | DTPa-HBV-Hib-TT/IPV | 0.66 (0.35-1.25) |
| 15 | *pcv13 vs pcv7* | Europe | 2-3-4 months | DTPa-HBV-Hib-TT/IPV | 1.35 (0.78-2.35) |
| 19 | *pcv13 vs pcv7* | Europe | 2-4 months | DTaP-IPV-Hib/MenC | 2.50 (0.74-8.43) |
| 22 | *pcv13 vs pcv7* | North America | 2-4-6 months | DTPa-HBV-Hib-TT/IPV | 1.59 (0.82-3.08) |

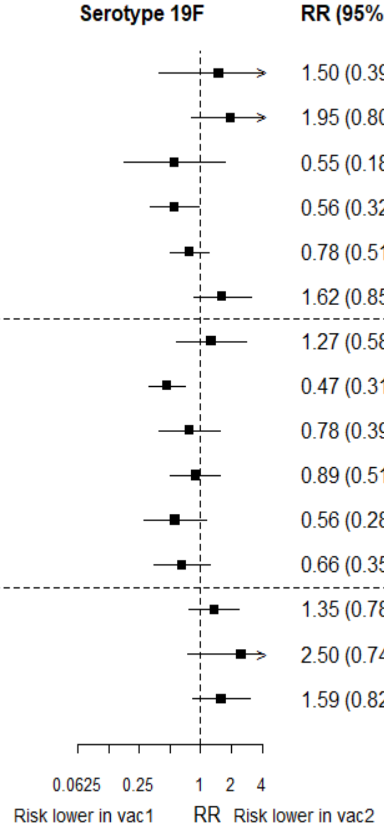

RR: relative risk; pcv: Pneumococcal conjugate vaccine; DTaP – diphtheria and tetanus toxoids, and acellular pertussis vaccine; DTwP – diphtheria and tetanus toxoids, and whole-cell pertussis vaccine; Hib-TT – Haemophilus influenzae type b vaccine (tetanus toxoid conjugate); HB – Hepatitis B vaccine; IPV – Inactivated polio vaccine; OPV – Oral polio vaccine; MenC – Meningococcal C vaccine; TT – tetanus toxoid conjugate; NA: not applicable.

Each solid line in the figure shows the RR from each trial. Black boxes and lines show the point estimates and confidence intervals for relative risks comparing vac1 vs vac2. co-adm vaccines are vaccines co-administered with PCV primary vaccine series. Information on co-administered vaccine is not always available. Concomitant vaccines in the bracket are those administered in some but not all of the study sites.

#### Supplementary Figure 51. Trial level relative risk for serotype 23F.

| ID | Comparison vac1 vs vac2 | Continent | Schedule-prime | co-adm vaccines | Serotype 23F | RR (95%CI) |
|---|---|---|---|---|---|---|
| 1 | *pcv7 vs pcv10* | Asia | 6-10-24 weeks | DTPw-HBV-Hib-TT + OPV | | 0.48 (0.20-1.15) |
| 1 | *pcv7 vs pcv10* | Europe | 2-4-6 months | DTPw-HBV-Hib-TT + IPV | | 2.30 (1.03-5.11) |
| 3 | *pcv7 vs pcv10* | Europe | 2-3-4 months | DTPa-HBV-Hib-TT/IPV | | 0.18 (0.06-0.58) |
| 7 | *pcv7 vs pcv10* | Europe | 2-3-4 months | DTPa-(HBV)-Hib-TT/IPV | | 0.24 (0.14-0.43) |
| 8 | *pcv7 vs pcv10* | Europe | 2-3-4 months | DTPa-(HBV)-Hib-TT/IPV | | 0.35 (0.23-0.54) |
| 9 | *pcv7 vs pcv10* | Europe | 2-4-6 months | DTPa-(HBV)-Hib-TT/IPV + MenC | | 0.47 (0.23-0.97) |
| 4 | *pcv13 vs pcv10* | Europe | 2-3-4 months | DTPa-HBV-Hib-TT/IPV | | 0.21 (0.11-0.38) |
| 27 | *pcv13 vs pcv10* | Africa | 14 weeks | DTPa-HBV-Hib-TT/IPV/Rotavirus | | 0.85 (0.70-1.04) |
| 27 | *pcv13 vs pcv10* | Africa | 6-14 weeks | DTPa-HBV-Hib-TT/IPV/Rotavirus | | 0.42 (0.20-0.85) |
| 27 | *pcv13 vs pcv10* | Africa | 6 weeks | DTPa-HBV-Hib-TT/IPV/Rotavirus | | 0.95 (0.72-1.25) |
| 5 | *pcv13 vs pcv10* | Europe | 2–3–4 months | DTPa-HBV-Hib-TT/IPV + (MenC-TT) | | 0.29 (0.18-0.45) |
| 6 | *pcv13 vs pcv10* | Asia | 2-4 months | DTPa-HBV-Hib-TT/IPV | | 0.39 (0.28-0.56) |
| 15 | *pcv13 vs pcv7* | Europe | 2-3-4 months | DTPa-HBV-Hib-TT/IPV | | 0.69 (0.38-1.24) |
| 19 | *pcv13 vs pcv7* | Europe | 2-4 months | DTaP-IPV-Hib/MenC | | 2.25 (0.90-5.62) |
| 22 | *pcv13 vs pcv7* | North America | 2-4-6 months | DTPa-HBV-Hib-TT/IPV | | 1.78 (0.72-4.37) |

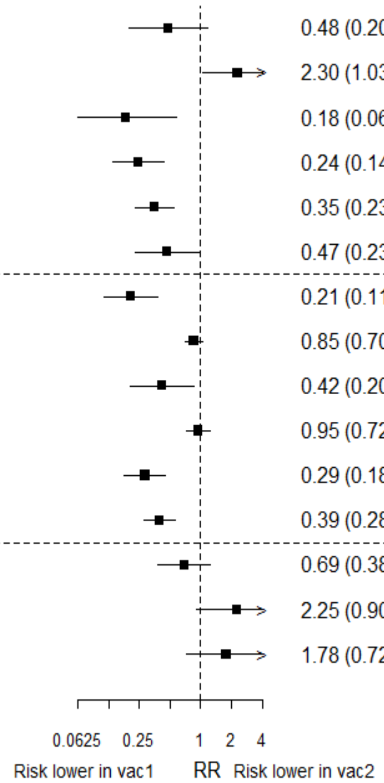

RR: relative risk; pcv: Pneumococcal conjugate vaccine; DTaP – diphtheria and tetanus toxoids, and acellular pertussis vaccine; DTwP – diphtheria and tetanus toxoids, and whole-cell pertussis vaccine; Hib-TT – Haemophilus influenzae type b vaccine (tetanus toxoid conjugate); HB – Hepatitis B vaccine; IPV – Inactivated polio vaccine; OPV – Oral polio vaccine; MenC – Meningococcal C vaccine; TT – tetanus toxoid conjugate; NA: not applicable.

Each solid line in the figure shows the RR from each trial. Black boxes and lines show the point estimates and confidence intervals for relative risks comparing vac1 vs vac2. co-adm vaccines are vaccines co-administered with PCV primary vaccine series. Information on co-administered vaccine is not always available. Concomitant vaccines in the bracket are those administered in some but not all of the study sites.

**Supplementary Figure 52. Trial level relative risk for serotype 1.**

| ID | Comparison vac1 vs vac2 | Continent | Schedule-prime | co-adm vaccines | RR (95%CI) |
|---|---|---|---|---|---|
| **4** | *pcv13 vs pcv10* | Europe | 2-3-4 months | DTPa-HBV-Hib-TT/IPV | 2.46 (0.49-12.48) |
| **27** | *pcv13 vs pcv10* | Africa | 14 weeks | DTPa-HBV-Hib-TT/IPV/Rotavirus | 1.42 (0.73-2.74) |
| **27** | *pcv13 vs pcv10* | Africa | 6-14 weeks | DTPa-HBV-Hib-TT/IPV/Rotavirus | 1.53 (0.63-3.73) |
| **27** | *pcv13 vs pcv10* | Africa | 6 weeks | DTPa-HBV-Hib-TT/IPV/Rotavirus | 1.56 (0.87-2.83) |
| **5** | *pcv13 vs pcv10* | Europe | 2–3–4 months | DTPa-HBV-Hib-TT/IPV + (MenC-TT) | 0.48 (0.09-2.57) |
| **6** | *pcv13 vs pcv10* | Asia | 2-4 months | DTPa-HBV-Hib-TT/IPV | 0.32 (0.11-0.96) |

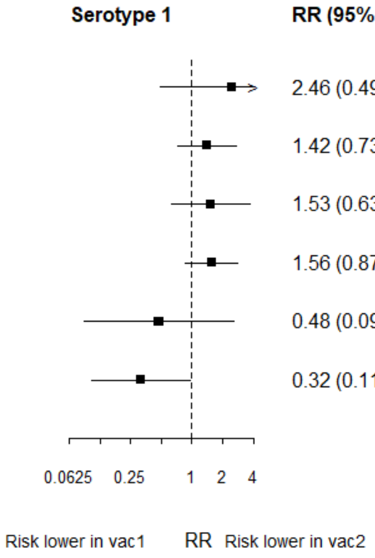

RR: relative risk; pcv: Pneumococcal conjugate vaccine; DTaP – diphtheria and tetanus toxoids, and acellular pertussis vaccine; DTwP – diphtheria and tetanus toxoids, and whole-cell pertussis vaccine; Hib-TT – Haemophilus influenzae type b vaccine (tetanus toxoid conjugate); HB – Hepatitis B vaccine; IPV – Inactivated polio vaccine; OPV – Oral polio vaccine; MenC – Meningococcal C vaccine; TT – tetanus toxoid conjugate; NA: not applicable.

Each solid line in the figure shows the RR from each trial. Black boxes and lines show the point estimates and confidence intervals for relative risks comparing vac1 vs vac2. co-adm vaccines are vaccines co-administered with PCV primary vaccine series. Information on co-administered vaccine is not always available. Concomitant vaccines in the bracket are those administered in some but not all of the study sites.

**Supplementary Figure 53. Trial level relative risk for serotype 5.**

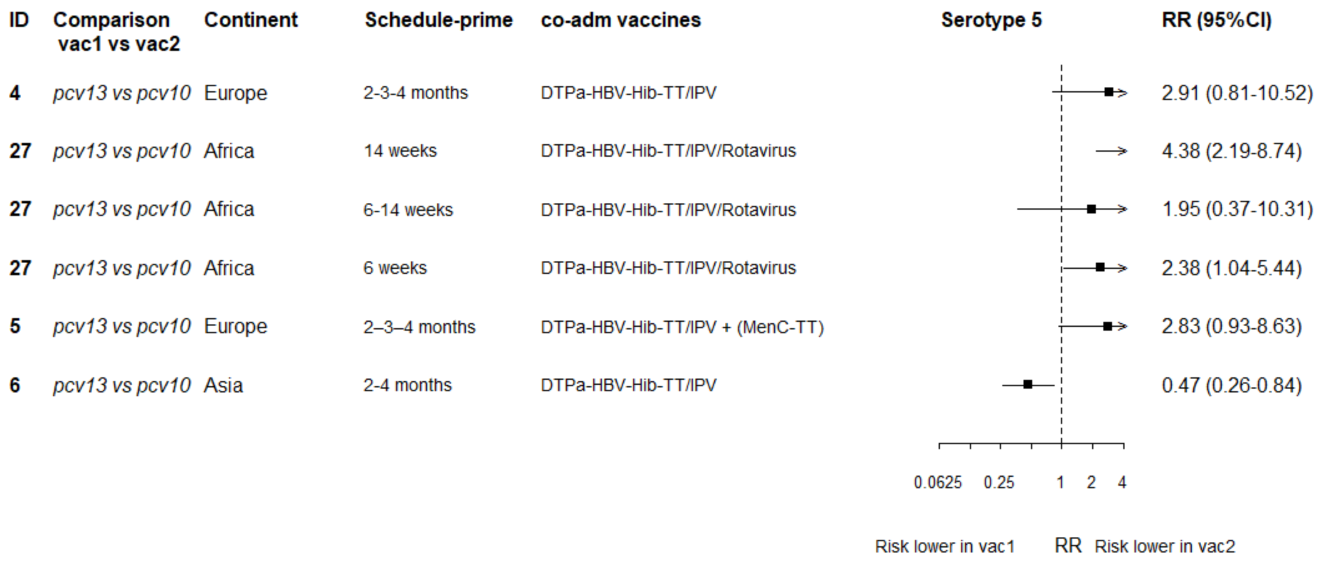

RR: relative risk; pcv: Pneumococcal conjugate vaccine; DTaP – diphtheria and tetanus toxoids, and acellular pertussis vaccine; DTwP – diphtheria and tetanus toxoids, and whole-cell pertussis vaccine; Hib-TT – Haemophilus influenzae type b vaccine (tetanus toxoid conjugate); HB – Hepatitis B vaccine; IPV – Inactivated polio vaccine; OPV – Oral polio vaccine; MenC – Meningococcal C vaccine; TT – tetanus toxoid conjugate; NA: not applicable.

Each solid line in the figure shows the RR from each trial. Black boxes and lines show the point estimates and confidence intervals for relative risks comparing vac1 vs vac2. co-adm vaccines are vaccines co-administered with PCV primary vaccine series. Information on co-administered vaccine is not always available. Concomitant vaccines in the bracket are those administered in some but not all of the study sites.

**Supplementary Figure 54. Trial level relative risk for serotype 7F.**

| ID | Comparison vac1 vs vac2 | Continent | Schedule-prime | co-adm vaccines | Serotype 7F | RR (95%CI) |
|---|---|---|---|---|---|---|
| 4 | *pcv13 vs pcv10* | Europe | 2-3-4 months | DTPa-HBV-Hib-TT/IPV | | 1.29 (0.56-2.97) |
| 27 | *pcv13 vs pcv10* | Africa | 14 weeks | DTPa-HBV-Hib-TT/IPV/Rotavirus | | 1.00 (0.73-1.36) |
| 27 | *pcv13 vs pcv10* | Africa | 6-14 weeks | DTPa-HBV-Hib-TT/IPV/Rotavirus | | 0.49 (0.13-1.88) |
| 27 | *pcv13 vs pcv10* | Africa | 6 weeks | DTPa-HBV-Hib-TT/IPV/Rotavirus | | 0.83 (0.59-1.16) |
| 5 | *pcv13 vs pcv10* | Europe | 2–3–4 months | DTPa-HBV-Hib-TT/IPV + (MenC-TT) | | 0.58 (0.31-1.09) |
| 6 | *pcv13 vs pcv10* | Asia | 2-4 months | DTPa-HBV-Hib-TT/IPV | | 0.77 (0.33-1.80) |

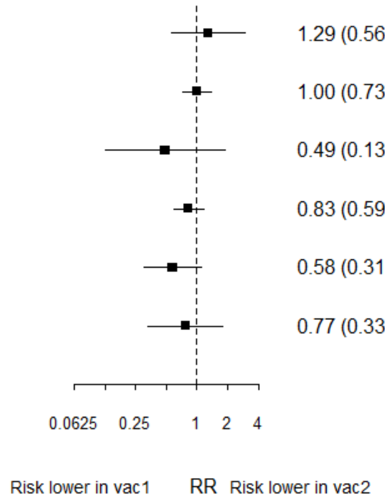

RR: relative risk; pcv: Pneumococcal conjugate vaccine; DTaP – diphtheria and tetanus toxoids, and acellular pertussis vaccine; DTwP – diphtheria and tetanus toxoids, and whole-cell pertussis vaccine; Hib-TT – Haemophilus influenzae type b vaccine (tetanus toxoid conjugate); HB – Hepatitis B vaccine; IPV – Inactivated polio vaccine; OPV – Oral polio vaccine; MenC – Meningococcal C vaccine; TT – tetanus toxoid conjugate; NA: not applicable.

Each solid line in the figure shows the RR from each trial. Black boxes and lines show the point estimates and confidence intervals for relative risks comparing vac1 vs vac2. co-adm vaccines are vaccines co-administered with PCV primary vaccine series. Information on co-administered vaccine is not always available. Concomitant vaccines in the bracket are those administered in some but not all of the study sites.

**Supplementary Figure 55. Study level association between geometric mean ratio and relative risk for each serotype in PCV7**

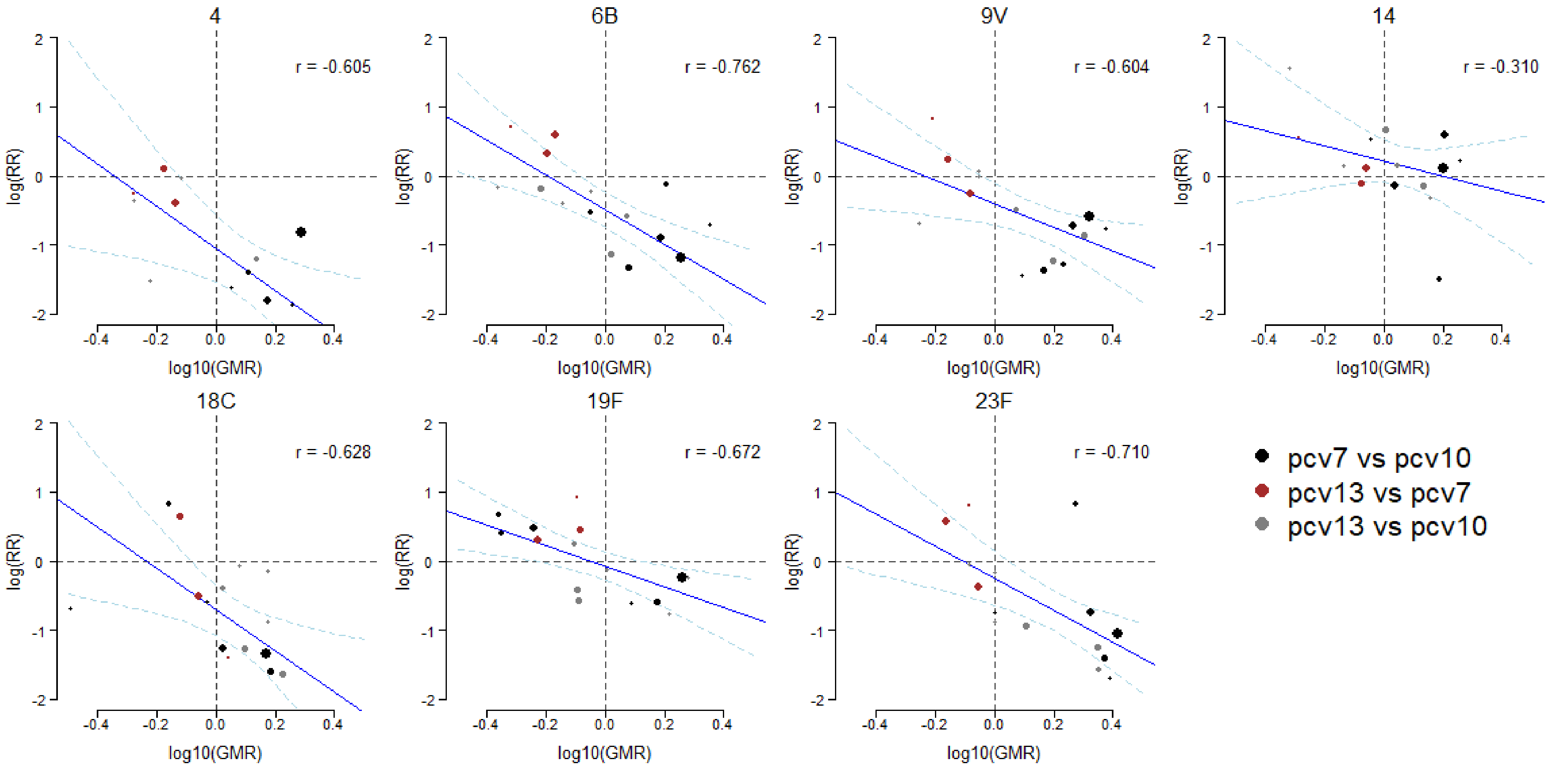

RR: relative risk; GMR: Geometric mean ratio; PCV: Pneumococcal conjugate vaccine. Each point shows results of a serotype specific head-to-head comparison between two vaccines from one study. Solid line shows the relationship between relative risk predicted from the crude model and geometric mean ratio. Dashed line shows the confidence intervals of predicted relative risk. Reference lines show geometric mean ratio equivalent to one (vertical) and relative risk equivalent to one (horizontal) which represent values associated with no difference between vaccines. Points sizes represent sample size of the trial. Each panel shows one PCV7 serotype (4, 6B, 9V, 14, 18C, 19F and 23F).
